# Transferability of European-derived Alzheimer’s Disease Polygenic Risk Scores across Multi-Ancestry Populations

**DOI:** 10.1101/2023.10.17.23297061

**Authors:** Aude Nicolas, Richard Sherva, Benjamin Grenier-Boley, Yoontae Kim, Masataka Kikuchi, Jigyasha Timsina, Itziar de Rojas, María Carolina Dalmasso, Xiaopu Zhou, Yann Le Guen, Carlos E Arboleda-Bustos, Maria Aparecida Camargos Bicalho, Maëlenn Guerchet, Sven van der Lee, Monica Goss, Atahualpa Castillo, Céline Bellenguez, Fahri Küçükali, Claudia Satizabal Barrera, Bernard Fongang, Qiong yang, Oliver Peters, Anja Schneider, Martin Dichgans, Dan Rujescu, Norbert Scherbaum, Jürgen Deckert, Steffi Riedel-Heller, Lucrezia Hausner, Laura Molina Porcel, Emrah Düzel, Timo Grimmer, Jens Wiltfang, Stefanie Heilmann-Heimbach, Susanne Moebus, Thomas Tegos, Nikolaos Scarmeas, Oriol Dols-Icardo, Fermin Moreno, Jordi Pérez-Tur, María J. Bullido, Pau Pastor, Raquel Sánchez-Valle, Victoria Álvarez, Han Cao, Nancy Y. Ip, Amy K. Y. Fu, Fanny C. F. Ip, Natividad Olivar, Carolina Muchnik, Carolina Cuesta, Lorenzo Campanelli, Patricia Solis, Daniel Gustavo Politis, Silvia Kochen, Luis Ignacio Brusco, Mercè Boada, Pablo García-González, Raquel Puerta, Pablo Mir, Luis M Real, Gerard Piñol-Ripoll, Jose María García-Alberca, Jose Luís Royo, Eloy Rodriguez-Rodriguez, Hilkka Soininen, Sami Heikkinen, Alexandre de Mendonça, Shima Mehrabian, Latchezar Traykov, Jakub Hort, Martin Vyhnalek, Katrine Laura Rasmussen, Jesper Qvist Thomassen, Yolande A.L. Pijnenburg, Henne Holstege, John van Swieten, Harro Seelaar, Jurgen A.H.R. Claassen, Willemijn J. Jansen, Inez Ramakers, Frans Verhey, Aad van der Lugt, Philip Scheltens, Jenny Ortega-Rojas, Ana Gabriela Concha Mera, Maria F. Mahecha, Rodrogo Pardo, Gonzalo Arboleda, Shahram Bahrami, Vera Fominykh, Geir Selbæk, Caroline Graff, Goran Papenberg, Vilmantas Giedraitis, Anne Boland, Jean-François Deleuze, Luiz Armando de Marco, Edgar Nunes de Moraes, Bernardo de Mattos Viana, Marco Túlio Gualberto Cintra, Teresa Juárez-Cedillo, Anthony Grsiwold, Tatiana Forund, Jonathan Haines, Lindsay Farrer, Anita DeStefano, Ellen Wijsman, Richard Mayeux, Margaret Pericak-Vance, Brian Kunkle, Alison Goate, Gerard D. Schellenberg, Badri Vardarajan, Li-San Wang, Yuk Yee Leung, Clifton Dalgard, Gael Nicolas, David Wallon, Carole Dufouil, Florence Pasquier, Olivier Hanon, Stéphanie Debette, Edna Grünblatt, Julius Popp, Bárbara Angel, Sergio Golger, Maria Victoria Chacon, Rafael Aranguiz, Paulina Orellana, Andrea Slachevsky, Christian Gonzalez-Billault, Cecilia Albala, Patricio Fuentes, Perminder Sachdev, Karen Mather, Richard L. Hauger, Victoria Merritt, Matthew Panizzon, Rui Zhang, Michael Gaziano, Roberta Ghidoni, Daniela Galimberti, Beatrice Arosio, Patrizia Mecocci, Vincenzo Solfrizzi, Lucilla Parnetti, Alessio Squassina, Lucio Tremolizzo, Barbara Borroni, Benedetta Nacmias, Paolo Caffarra, Davide Seripa, Innocenzo Rainero, Antonio Daniele, Fabrizio Piras, EADB, Hampton L Leonard, Jennifer S. Yokoyama, Mike A Nalls, Akinori Miyashita, Norikazu Hara, Kouichi Ozaki, Shumpei Niida, Julie Williams, Carlo Masullo, Philippe Amouyel, Pierre-Marie Preux, Pascal Mbelesso, Bébène Bandzouzi, Andy Saykin, Frank Jessen, Patrick Kehoe, Cornelia Van Duijn, Nesrine Ben Salem, Ruth Frikke-Schmidt, Lofti Cherni, Michael D. Greicius, Magda Tsolaki, Pascual Sánchez-Juan, Marco Aurélio Romano Silva, Tenielle Porter, Simon M Laws, Kristel Sleegers, Martin Ingelsson, Jean-François Dartigues, Sudha Seshadri, Giacomina Rossi, Laura Morelli, Mikko Hiltunen, Rebecca Sims, Wiesje van der Flier, Ole Andreassen, Humberto Arboleda, Carlos Cruchaga, Valentina Escott-Price, Agustín Ruiz, Kun Ho Lee, Takeshi Ikeuchi, Alfredo Ramirez, Jungsoo Gim, Mark Logue, Jean-Charles Lambert

## Abstract

A polygenic score (PGS) for Alzheimer’s disease (AD) was recently derived from data on genome-wide significant loci in European ancestry populations. We applied this PGS to populations in 17 European countries and observed a consistent association with AD risk, age at onset, and cerebrospinal fluid levels of AD biomarkers, independently of apolipoprotein E *(APOE)*. This PGS was also associated with the AD risk in many other populations of diverse ancestries. A cross-ancestry polygenic risk score (PRS) improved the association with AD risk in most of the multi-ancestry populations tested when the *APOE* region was included. Lastly, we found that the PGS/PRS, captured AD-specific information because the association weakened as the diagnosis was broadened. In conclusion, a simple PGS captures the AD-specific genetic information that is common to populations of different ancestries, but studies of more diverse populations are still needed for a better characterization of the AD genetics.

---

Over the last 15 years, genome-wide association studies (GWASs) have fostered the development of powerful approaches for characterizing disease processes and proposed diagnostic/prognostic tools such as polygenic scores (PGS)^1,2^. Given the high estimated heritability (60-80%, in twin studies) of Alzheimer’s disease (AD)^3^, a number of PGSs have been developed and their associations with AD risk or related phenotypes have been almost systematically reported^4,5,6,7,8,9,10^. However, comparisons across studies are complicated by marked differences in the populations analyzed, PGS-calculation methods, the summary statistics used, and the variants included^11^. Furthermore, most PGSs have been developed from studies of European-ancestry populations, and only a few studies have investigated PGS performance in populations of different ancestries^12,13,14,15^.

In this manuscript, we first generated a PGS (PGS^ALZ)^ including the genome-wide significant, independent sentinel single nucleotide polymorphisms (SNPs) at the loci reported by Bellenguez et al.^16^ excluding the *apolipoprotein E* (*APOE)* locus (n=83; see Supplementary Table 1 for the list of variants). We studied the associations between PGS^ALZ^ and AD risk or relevant endophenotypes in populations from 17 European countries. We next extended PGS^ALZ^ study to populations of diverse ancestries from Asia, Africa, Latin and North America. Finally, as already developed in other complex human diseases^17,18,19,20^, we generated a cross-ancestry polygenic risk score (PRS) by integrating GWAS summary statistics from multiple populations to potentially improve PGS^ALZ^ predictive performance^21,22^.

We first evaluated the association between PGS^ALZ^ and the risk of developing AD in case-control studies of European countries (see Supplementary Table 2 for population description and adjustments used per population; Supplementary Fig. 1-3 for PGS^ALZ^ distributions). PGS^ALZ^ was significantly associated with AD risk irrespective of *APOE* adjustment (Extended Fig. 1A and Supplementary Fig. 4). PGS^ALZ^ was similarly associated with AD risk in men and in women (Extended Fig. 1B and Supplementary Fig. 6) and with a younger age at onset (Extended Fig. 2). It is noteworthy that when the PGSs were adjusted for difference in PGS^ALZ^ distribution between the European populations, the association with AD remained similar (Supplementary Fig. 5).

As we did not identify any potential bias/heterogeneity when comparing PGS^ALZ^ in the European populations, we performed a combined analysis (mega-analysis) of our European datasets to assess the risk of developing AD within various PGS^ALZ^ strata: 0-2%, 2-5%, 10-20%, 20-40%, 60-80%, 80-90%, 90-95%, 95-98%, and 98-100% with the 40-60% PGS^ALZ^ stratum as the reference. We also generated a PGS that included both the sentinel AD GWAS loci and the two SNPs defining the ε2/ε3/ε4 *APOE* alleles. As expected, the risk of developing AD in the most extreme strata was particularly high when *APOE* was included (Fig. 1A). The association with PGS^ALZ^ was also significant in all strata analyzed, irrespective of *APOE* adjustment. In the 0-2% and 98-100% strata, PGS^ALZ^ was respectively associated with a more than 2-fold decrease in the AD risk and a more than 3-fold increase in the AD risk compared with the 40-60% stratum (Fig. 1A and Supplementary Table 3).

**Figure 1:**
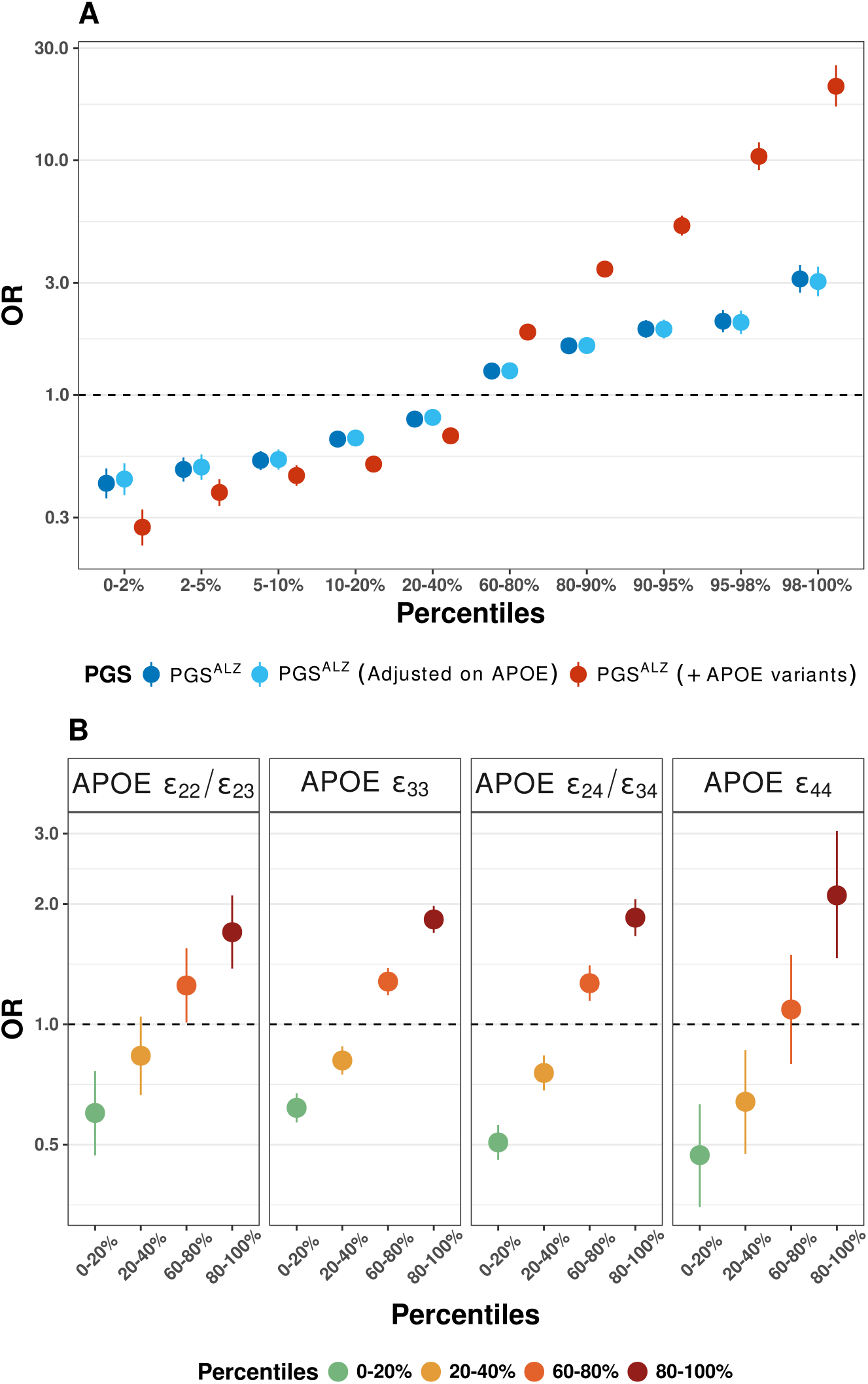
Associations between the various PGSs and the risk of developing AD, as a function of *APOE* status. **(A)** The risk of developing AD, by PGS^ALZ^ stratum (0-2%, 2-5%, 10-20%, 20-40%, 60-80%, 80-90%, 90-95%, 95-98%, and 98-100%). The 40-60% PGS^ALZ^ stratum was used as the reference. **(B)** Risk of developing AD, by PGS^ALZ^ stratum (0-20%, 20-40%, 60-80%, and 80-100%) and the *APOE* genotype (by grouping together the ε2ε2/ε2ε3, ε3ε3, ε2ε4/ε3ε4, ε4ε4 carriers). The 40-60% PGS^ALZ^ stratum was used as the reference.

Since these results suggested an association for PGS^ALZ^ independently of *APOE*, we took advantage of our mega-analysis to determine how PGS^ALZ^ interacts with the *APOE* genotypes. We found a limited interaction between PGS^ALZ^ and the number *APOE* ε4 alleles on the risk of developing AD (p=3×10^-4^). We next stratified the mega-analysis into four *APOE* genotype groups (ε2ε2/ε2ε3, ε3ε3, ε2ε4/ε3ε4, and ε4ε4) and assessed the association between PGS^ALZ^ and the AD risk per quintile (0-20%, 20-40%, 60-80%, and 80-100%) for each subpopulation (reference: 40-60% stratum). PGS^ALZ^ was similarly associated with the AD risk in all the strata, even if a stronger association might be present among ε4ε4 carriers (Fig. 1B and Supplementary Table 4).

To determine whether PGS^ALZ^ is associated with AD pathophysiological processes, we analyzed GWAS data of CSF levels of Aβ42, tau and p-tau (n=13,051 individuals) as previously described^23^. PGS^ALZ^ was associated with a decrement in Aβ_42_ levels and an increment in tau and p-tau levels whatever the adjustment on *APOE* (Fig. 2A,B and Supplementary Fig. 7). We also checked the available samples for a possible association between PGS^ALZ^ and Aβ_42_ levels, tau and p-tau levels in quintiles (0-20%, 20-40%, 60-80%, and 80-100%); again, the 40-60% stratum served as a reference. As expected, PGS^ALZ^ was associated with the lowest and highest levels of p-tau and Aβ_42_ in the 0-20% strata and, conversely, the highest and lowest levels of p-tau and Aβ_42_ in the 80-100% stratum. (Fig. 2C and Supplementary Table 5).

**Figure 2:**
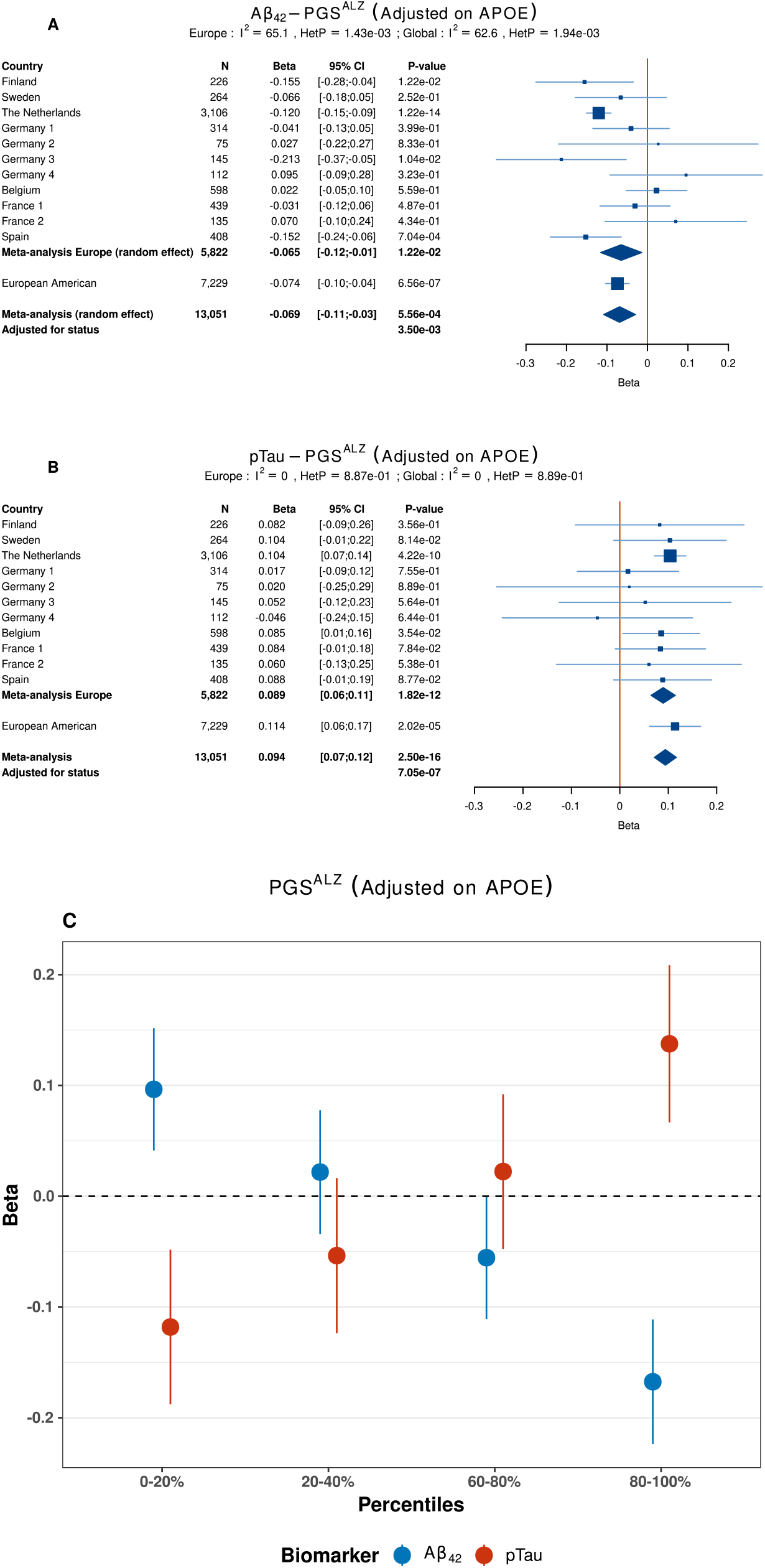
Association of PGS^ALZ^ with the level of Aβ_42_ and p-tau in the cerebrospinal fluid **(A)** across European-ancestry populations and **(B)** according to PGS^ALZ^ strata (0-20%, 20-40%, 60-80% and 80-100%). The 40-60% PGS^ALZ^ stratum was used as the reference. Ncases, number of cases; Ncontrols, number of controls, OR, Odds ratio per standard deviation. The lines in the Forest plots indicate the 95% confidence interval for the ORs. If HetP <0.05, random-effect is shown for the meta-analysis results.

We then extended the PGS^ALZ^ analyses to other European-ancestry populations (USA, Australia), populations from India, East Asia (China, Japan and Korea), North Africa (Tunisia), sub-Saharan Africa (Central African Republic/the Congo Republic), South America (Argentina, Brazil, Chile, and Colombia), and African-, Native- and Latino American ancestry populations from US studies (i.e. more than 75% African or Native American ancestry or self-reporting for Latino American populations; Extended Fig. 3A; supplementary Table 2 for population description). With the exception of the analyses for Korea and Japan (where respectively 72 and 74 SNPs were available), most PGSs were constructed from 78 to 85 SNPs (including APOE variants; Supplementary Table 1; Supplementary Fig. 8-10 for PGS^ALZ^ distributions). The strength of the *APOE* ε4-AD association differed by population, as previously described in the literature^24,25^. ORs ranged from 1.36 in sub-Saharan Africa to 5.46 in Maghreb (Extended Fig 3B).

As expected, the PGS^ALZ^ association with AD risk was strongest in European-ancestry populations in the USA or Australia. PGS^ALZ^ was also significantly associated with the AD risk in Maghreb, East Asia, Latino American and African American populations (Fig. 3A and Supplementary Fig. 11). Lastly, PGS^ALZ^ was not associated with AD risk in the sub-Saharan African and Indian populations; this might be related to the small sample size and corresponding lack of statistical power. PGS^ALZ^ was associated with a younger age at onset in most of the populations studied, with the notable exception of the Chinese and Korean populations (Extended Fig. 4). It is noteworthy that the APOE ε2/ε3/ε4 alleles influenced age at onset in the two latter populations (Supplementary Fig. 12).

To refine our analysis of these populations of diverse ancestries, we calculated the association between AD and PGS^ALZ^ quintile (0-20%, 20-40%, 60-80%, and 80-100%; reference: 40-60% stratum) and meta-analyzed them per ancestry (Fig. 3B and C, Supplementary Table 6 and Supplementary Table 7). The Indian, Maghreb and sub-Saharan African populations were excluded because of the small sample size. The strength of the association with PGS^ALZ^ decreased from the European American, East Asian, and Latino American populations to the African American population, in that order (Fig. 3B and supplementary Table 6). PGS^ALZ^ generated from European-ancestry population GWAS performed poorly in African-ancestry populations.

**Figure 3:**
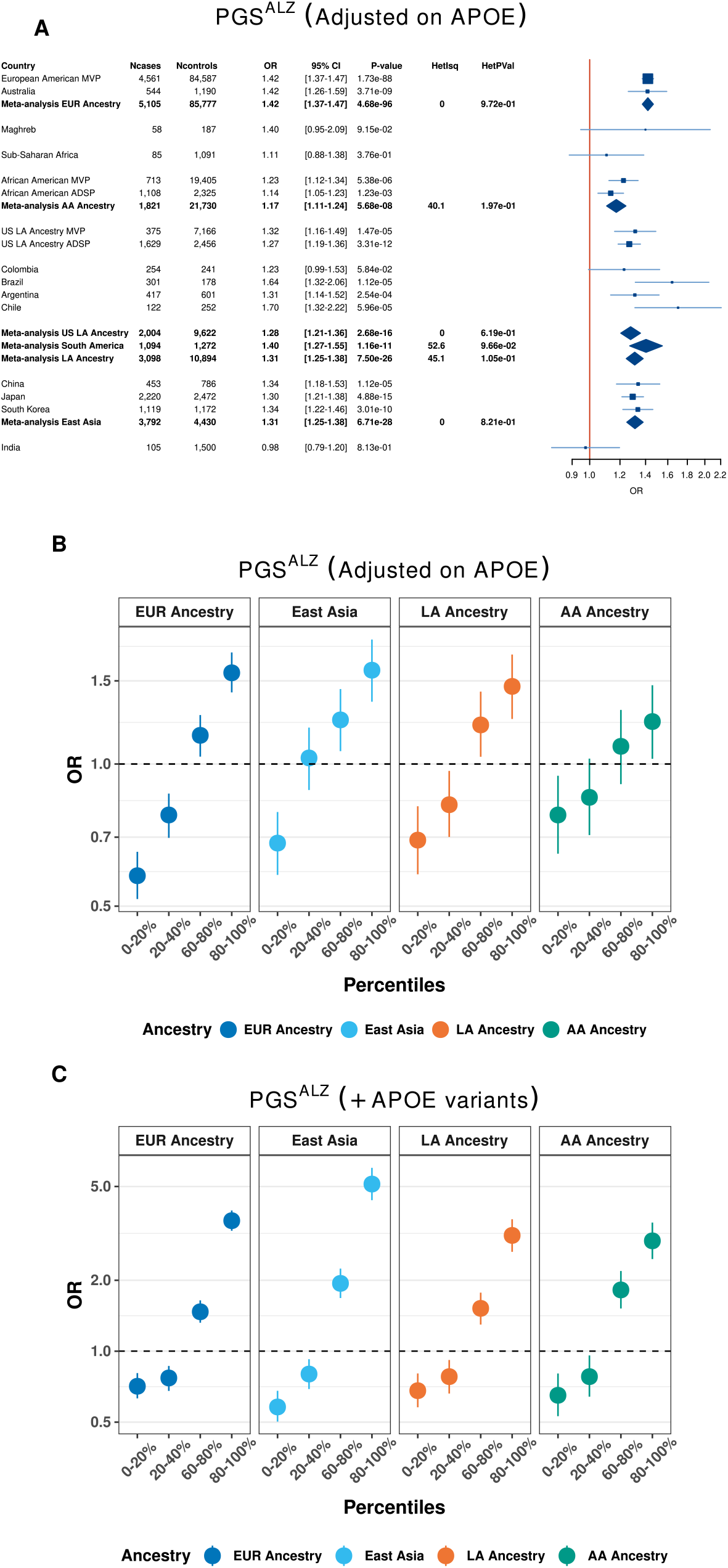
Association of PGS^ALZ^ across multi-ancestry populations. **(A)** Association of PGS^ALZ^ with the risk of developing AD in multi-ancestry populations. European-ancestry meta-analysis includes MVP and Australia. African-American-ancestry (more than 75% AA ancestry) meta-analysis includes MVP and ADSP. East-Asia meta-analysis includes China, Korea and Japan. Latino-American ancestry (self-reporting) meta-analysis includes MVP and ADSP. South-America meta-analysis includes Argentina, Brazil, Chile and Colombia. **(B)** Risk of developing AD according to PGS^ALZ^ (adjusted or not for *APOE* or included *APOE* variants) strata (0-20%, 20-40%, 60-80% and 80-100%) in multi-ancestry populations. The 40-60% PGS^ALZ^ stratum was used as the reference in each population and results were meta-analyzed. European-ancestry meta-analysis includes MVP and Australia. African-American ancestry meta-analysis includes MVP and ADSP. East-Asia meta-analysis includes China, Korea and Japan. Latin-American ancestry meta-analysis includes MVP and ADSP. South-America meta-analysis includes Argentina, Brazil, Chile and Colombia. Ncases, number of cases; Ncontrols, number of controls; OR, Odds ratio per standard deviation. The lines in the Forest plots indicate the 95% confidence interval for the ORs. If HetP <0.05, random effect is shown for the meta-analysis results. EUR, European; LA, Latino-American; AA, African American.

This latter observation was strengthened by analyzing the association between PGS^ALZ^ and AD risk as a function of the African American admixture. The strength of the association decreased as the percentage of African-ancestry increased and ultimately reached a level similar to that observed in our sub-Saharan African population: the association between PGS^ALZ^ and the AD risk in populations in whom more than 90% of the members were of African ancestry had an OR of 1.09 (95%CI 0.98-1.21, P=1.4×10^-1^, adjusted on *APOE*). It is noteworthy that a similar pattern was observed in the Alzheimer Disease Sequencing Project (ADSP) Native American population: the strength of the association decreased as the Native American-ancestry percentage increased from OR=1.21 (95%CI 1.12-1.32), P=5.3×10^-6^ and OR=1.14 (95%CI 1.05-1.25), P=2.6×10^-3^ to OR=1.12 (95%CI 1.02-1.24, P=1.4×10^-2^ in the populations with more than 50%, 75% and 90% of individuals of Native American-ancestry, respectively; adjusted for *APOE*. A similar finding was seen in Chilean and Argentinian populations: the PGS^ALZ^ association weakened as the proportion of Native American ancestry rose^14^.

We next checked whether we had fully captured the genetic information within the GWAS-defined loci in the non-European admixed populations. To this end, we developed a PGS (PGS^ALZ+^) that included other SNPs associated with AD risk in non-European ancestry populations (p<10^-3^) at the European-GWAS-defined loci (for details, see Online Methods). We used the summary statistics generated by Kunkle et al.^26^, Lake et al.^27^ and Shigemizu et al.^28^, and added 30, 13 and 47 variants to the initial 83 PGS^ALZ^ variants for Latino American, East Asian and African American ancestries, respectively (Supplementary Table 8). We did not detect any increment in the strength of the PGS^ALZ+^ association with the AD risk or in PGS^ALZ+^ predictive performance, relative to PGS^ALZ^ (Supplementary Table 9).

By initially restricting our analyses to the genome-wide significant loci from European ancestry AD GWAS, we likely excluded genetic information associated with the risk of developing AD in these European populations and (especially) non-European multi-ancestry populations (for which ancestry-specific loci may exist). Furthermore, the effect sizes used to construct PGS^ALZ^ were extracted from European ancestry populations without taking account population differences. To deal with these various questions, we used the Bayesian polygenic modeling method PRS-CSx to build a cross-ancestry PRS (PRS)^20^ which re-estimates variant effect sizes, by coupling diverse summary statistics with external ancestry-matched allele frequencies and local Linkage Disequilibrium structure, according to a sparseness parameter of the genetic architecture of AD. We used GWAS summary statistics generated from European (36,569 AD cases and 63,137 controls), African American (2,784 AD cases and 5,222 controls), Latino American (1,088 AD cases and 1,152 controls) and East Asian (3,962 AD cases and 4,074 controls) populations^26,27,28^. Polygenic Risk Scores (PRS), all adjusted for population structure, were generated in multi-ancestry populations from the Million Veteran Program (MVP; European, Latino American and African American ancestries), EPIDEMCA (Sub-Saharan Africa ancestry) and GARD studies (East Asian ancestry; Supplementary Fig. 13).

We assessed potential increment of PRS association with AD risk and predictive performance when the summary statistics of the European, African American, Latino American or East Asian populations were used independently (respectively, PRS^EUR^, PRS^AA^, PRS^LA^, PRS^EA^), or when they were combined (PRS^COMB^) at multiple sparseness parameter (10^-8^, 10^-7^, 10^-6^, 10^-5^, 10^-4^, 10^-2^ and 1). We initially excluded the APOE *region* to enable a comparison with PGS^ALZ^. We did not observe any increases in the association with AD risk or predictive performance in the different multi-ancestry populations (Fig. 4, supplementary Figure 14, supplementary Table 10) with the exception of the Latin American MVP population. However, we cannot exclude the possibility that this improvement is due to overfitting. Next, we included the *APOE* region when generating the different PRSs. While no impact was observed in European-ancestry populations when comparing PRS^EUR^ and PRS^COMB^), we detected a potential increment in both the strength of association with the AD risk and predictive performance when comparing PRS^EUR^ and PRS^COMB^ for all other populations, indicating that a cross-ancestry PRS is more effective than a PRS constructed solely from European summary statistics when the *APOE* region is included (whatever the global shrinkage parameter used; Fig. 5, supplementary Figure 14, supplementary Table 10).

**Figure 4:**
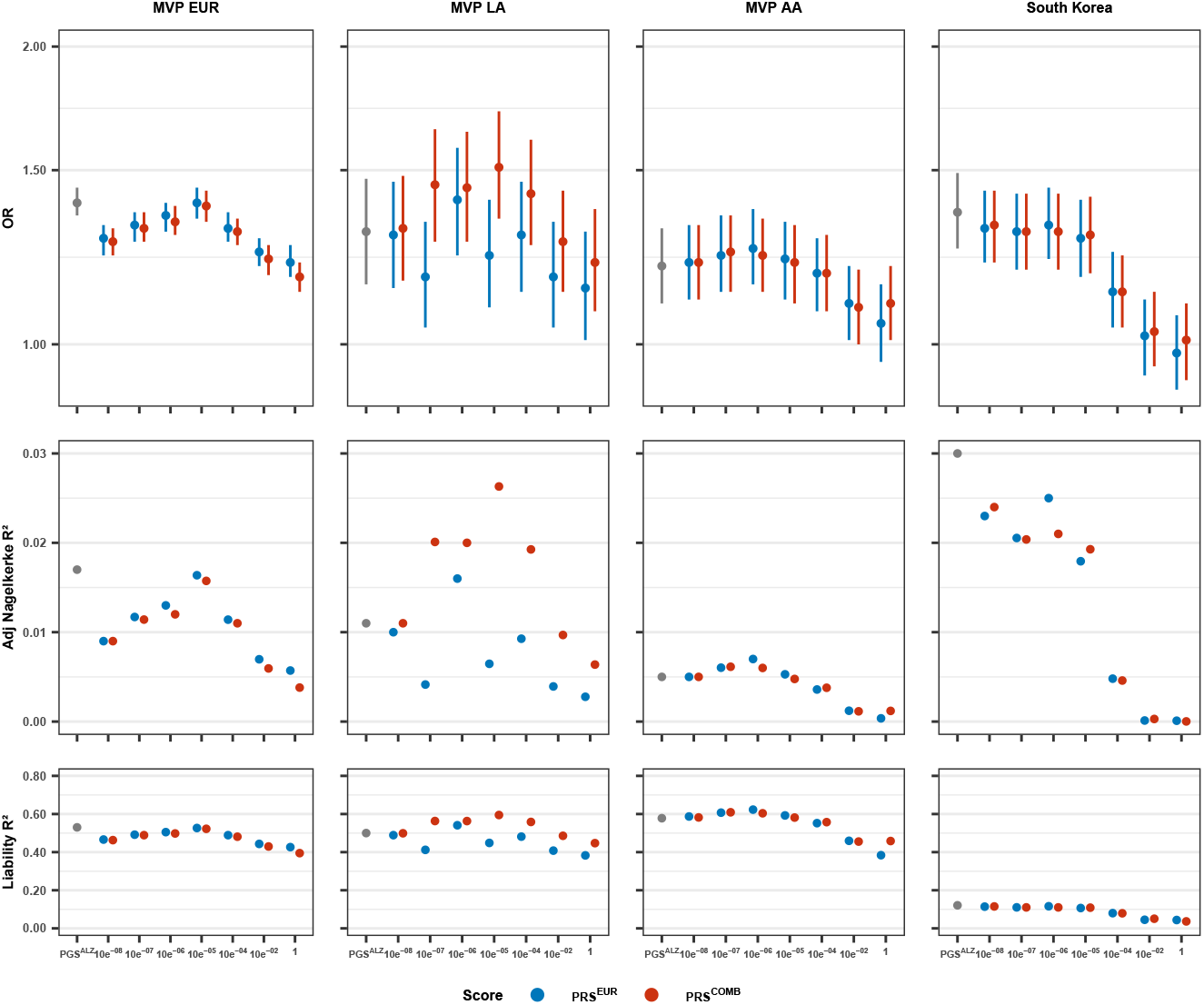
Comparison of the association of PGS^ALZ^ or PRS (excluding *APOE* region) with AD risk and the corresponding predictive values (adjusted Nagelkerke R^2^ and Liability R^2^). All PGS^ALZ^ and PRS were adjusted for difference in distribution between populations; OR, Odds ratio per standard deviation; PRS^EUR^ were generated by using only European ancestry summary statistics; PRS^COMB^ were generated by combining European, African American (AA), Latin-American (LA) and East Asian ancestry summary statistics. Sparseness parameter at 10^-8^, 10^-7^, 10^-6^, 10^-5^, 10^-4^, 10^-3^, 10^-2^ or 1.

**Figure 5:**
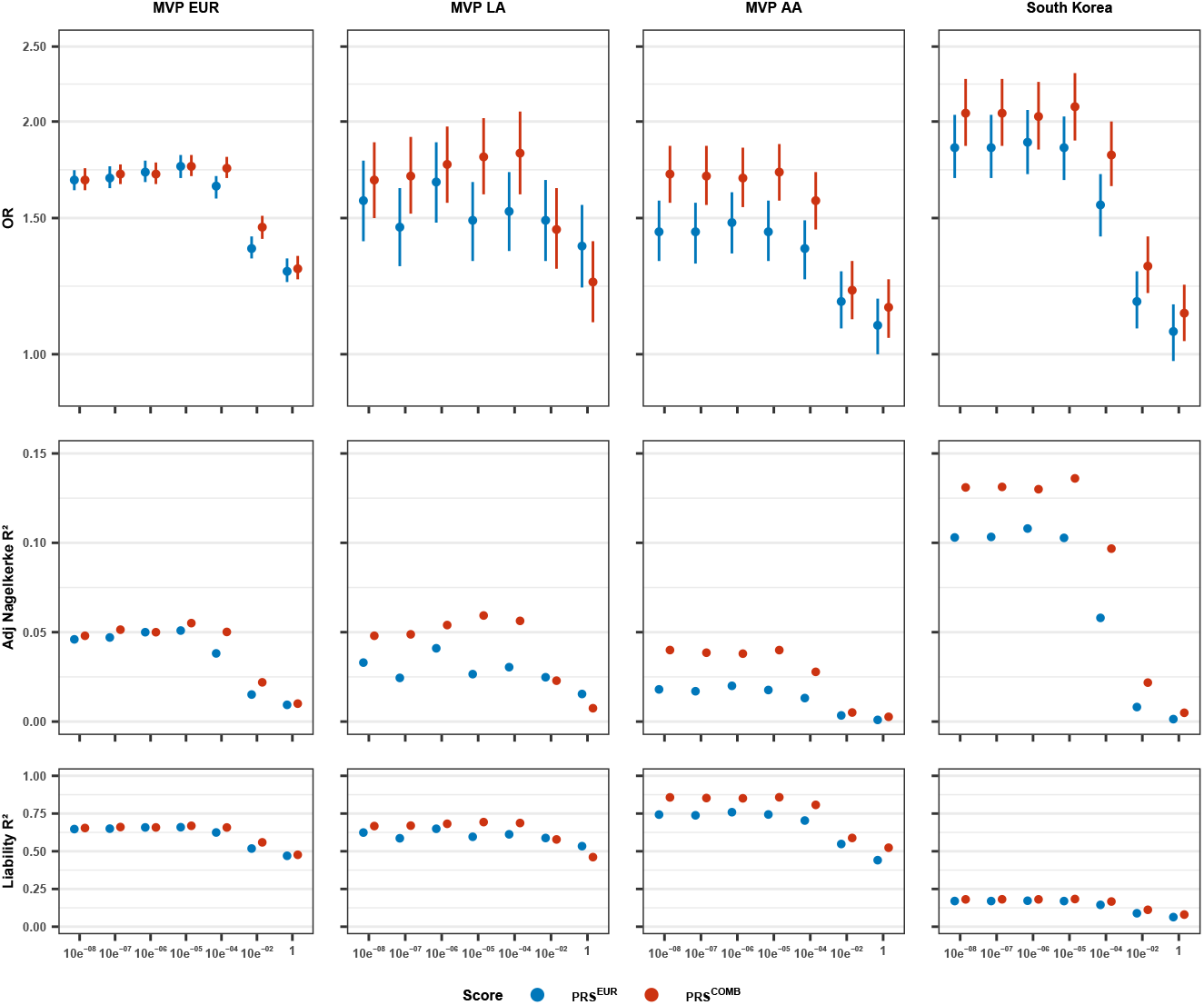
Association of PRS (including *APOE* region) with AD risk and the corresponding predictive values (adjusted Nagelkerke R^2^ and Liability R^2^). All PGS^ALZ^ and PRS were adjusted for difference in distribution between populations; OR, Odds ratio per standard deviation; PRS^EUR^ were generated by using only European ancestry summary statistics; PRS^COMB^ were generated by combining European, African American (AA), Latin-American (LA) and East Asian ancestry summary statistics. Sparseness parameter at 10^-8^, 10^-7^, 10^-6^, 10^-5^, 10^-4^, 10^-3^, 10^-2^ or 1.

Lastly, we leveraged the MVP data to determine how the association between PGS^ALZ^ or PRS^COMB^ (APOE region excluded) and the risk of AD changed in multi-ancestry populations as a function of diagnostic specificity. That is, we looked at how a PGS^ALZ^/ PRS^COMB^ derived from AD case/control performed when the applied to cohorts when the diagnosis was broadened to dementia. In all the multi-ancestry population studied, the association between PGS^ALZ^/ PRS^COMB^ and the AD risk weakened as the diagnosis became less specific (Fig. 6 and Supplementary Table 11).

**Figure 6:**
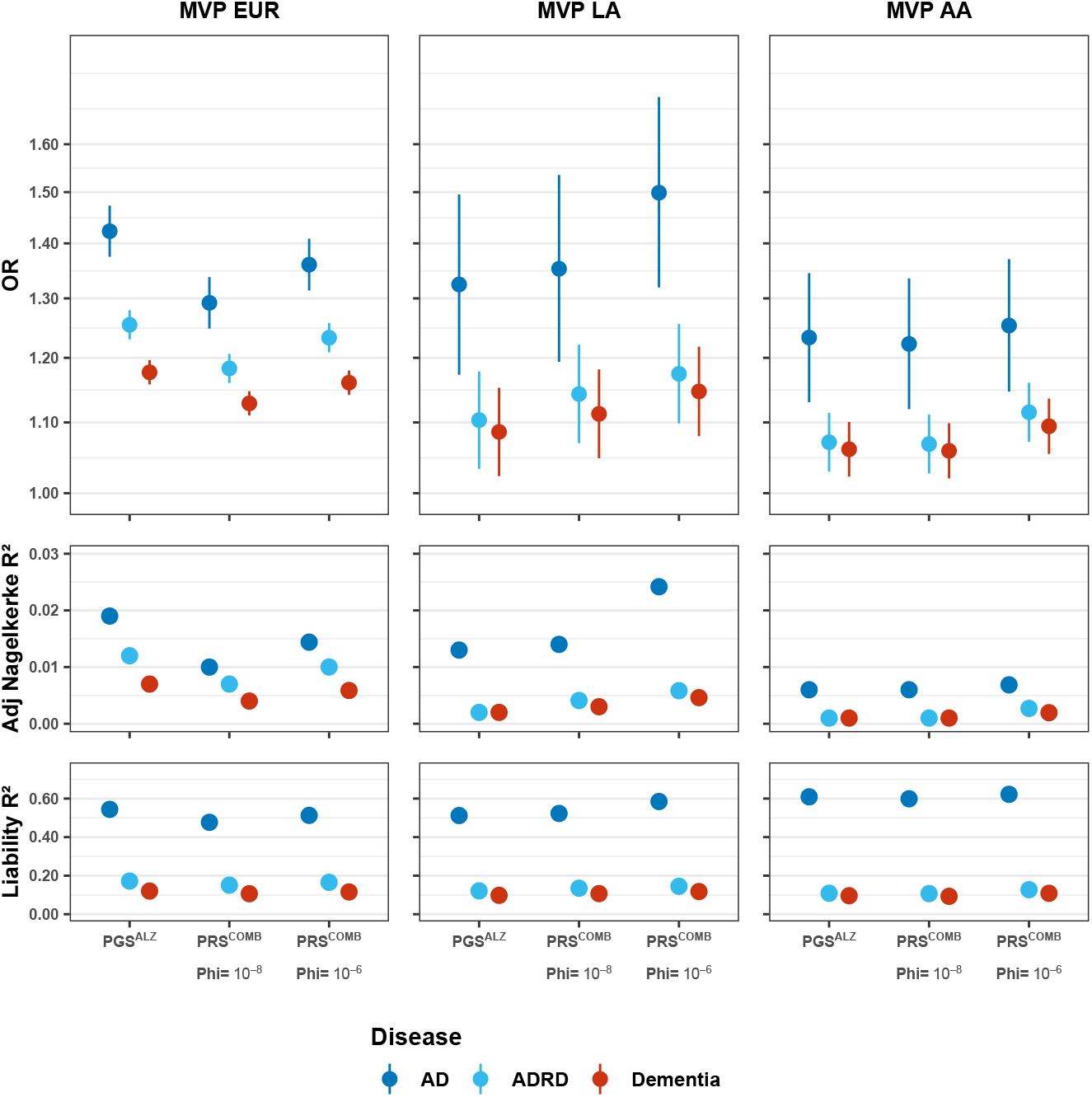
Association of PGS^ALZ^ or PRS^COMB^ (excluding the *APOE* region) with AD, AD and related dementia (ADRD) and dementia in MVP and the corresponding predictive values (adjusted Nagelkerke R^2^ and Liability R^2^). PGS^ALZ^ and PRS were adjusted for *APOE* and for difference in distribution between populations; OR, Odds ratio per standard deviation; PRS-CSx were generated by combining European (EUR), African American (AA) and Latin-American (LA) and East Asian ancestry summary statistics. Sparseness parameter at 10^-8^ and 10^-6^.

Several major points can thus be highlighted from our work: (i) In European populations, the associations of PGS^ALZ^ with AD risk are potentially slightly impacted by the *APOE* genotype (suggesting two independent genetic entities for sporadic AD; *APOE* ε4-associated sporadic AD, and *APOE* ε4-unassociated sporadic AD as previously proposed^29^); (ii) this simple PGS^ALZ^ based on the largest GWAS from European populations and the resulting European GWAS-defined loci appears to be enough to detect AD genetic risk in most of the different ancestry populations. Our results thus suggest that most of the different ancestry populations are likely to be affected by shared pathophysiological processes driven in part by genetic risk factors; (iii) Conversely, it is observed in the genetics of complex traits^30^ and other multifactorial diseases^17,31,32^, a cross-ancestry PRS built with a Bayesian polygenic modelling method did not systematically outperform a simple PGS^ALZ^ (when the APOE locus is excluded). The small population size of GWAS for the different ancestry populations can significantly limit the power of the PRS-CSx approach, potentially explaining this observation. However, this may also indicate that a high proportion of AD genetic risk is already accounted for by the European ancestry GWAS-defined loci; (iv) When PRS includes the *APOE* region, this region appears to likely contain additional multi-ancestry genetic variability as already proposed^33,34,35,36^; (v) Finally, the PGS/PRS associations mainly captures genetic information related to AD as their association weakened as the diagnosis was broadened. This observation suggests that the quality of the clinical diagnosis can interfere with the measurement of the association between the PGS/PRS and the AD risk in a given population.

In conclusion, our study of diverse ancestry populations and AD highlights the importance of cross-ancestry analyses for characterizing the genetic complexities of this devastating disease and to evaluate AD risk assessments in various populations. However, the field of AD genetics is still limited by the size of GWASs in these diverse ancestry populations. In addition, it is likely that the different ancestry populations will differ strongly regarding rare/very rare variants associated with the AD risk. this could clearly impact PRSs association with AD risk and their predictive performances^37^. Both GWAS and sequencing studies of more diverse populations are thus needed for a better characterization of AD genetics.

## Supporting information

Supplementary Informations

## Acknowledgments

We thank all the study participants, researchers, and staff for contributing to or collecting the data. We also thank the staff at the University of Lille’s high-performance computing service. This work was funded by a grant (European Alzheimer&Dementia DNA BioBank, EADB) from the EU Joint Programme – Neurodegenerative Disease Research (JPND) and La Fondation Recherche Alzheimer. A.N. was funded by Fondation pour la Recherche Médicale (EQU202003010147) and La Fondation Recherche Alzheimer. UMR1167 is also funded by the Inserm, Institut Pasteur de Lille, Lille Métropole Communauté Urbaine, and the French government’s LABEX DISTALZ program (development of innovative strategies for a transdisciplinary approach to Alzheimer’s disease). Full consortium acknowledgements and funding are given in the Supplementary Note. We thank David Fraser for his help.

## Author contributions

**Project coordination**: A.N., J-C.L; **data collection coordination**: A.N., Y.LG., J.G., M.D.G., S.V.D.L, E.N.D.M., J-F.D., H.A., V.E-P., A.Rui., K.H.L., T.I., A.Ram., M.L., J-C.L; **Data analyses**: A.N, R.She., B.G-B., Y.K., M.K., J.T., I.D.R., C.D., X.Z., Y.L.G. C.E.A-B., M.A.C.B., M.Gue., S.v.d.L., M.Gos., A.C., C.B., F.K; **EADB sample contribution**: I.d.R., A.C., S.V.D.L,., C.B., F.K., O.P., A.Sch., M.D., D.R., N.Sch., D.J., S.R-H., L.H., L.M.P., E.D., O.G., J.Wilt., S.H-H., S.Moe., T.T., N.Sca., J.C., F.M., J.P-T., M.J.B., P.P., R.S-V., V.Á. M.B., P.G-G., R.Pue., P.Mir., L.M.R., G.P-R., J.M.G-A., J.L.R., E.R-R., H.Soi., T.K., A.d.M., S.Meh., J.Hor., M.V., K.L.R., J.Q.T., Y.A.P., H.H. J.v.S., H.See., J.A.H.R.C., W.J.S., I.Ram., F.V., A.v.d.L. P.Sch., C.G., G.P., V.G., G.N., C.Duf., F.P., O.H., S.D., A.B., J-F.Del., E.G., J.P., D.G., B.Aro., P.Mec., V.S., L.P., A.Squ., L.T., B.Bor., B.N., P.C., D.S., I.Rai., A.Dan., J.Will., C.Mas., P.A., F.J., P.K., C.V.D., R.F-S., T.M., P.S-J., K.S., M.I., G.R., M.H., R.Sim., W.v.d.F., O.A., A.Rui., A.Ram., J-C.L., **Australian sample:** T.P., S.M.L.; **MVP sample contribution:** R.She., R.H., V.M., M.P., R.Z., M.Gaz., M.L.**; Salsa sample contribution:** M.Gos., C.S.B., B.F., Q.Y. S.S.; **ADSP sample contribution:** A.Gri., T.F., C.Cru., J.Hai., L.F., A.Des., E.W., R.M., M.P-V., B.K., A.Goa., G.D.S., B.V., L-S.W., Y.Y.L., C.Dalg., A.Say., H.L.L., J.S.Y., M.A.N., S.S.**; Africa sample contribution:** M.Gue., P-M.P., P.Mbe, B.Ban, N.B.S., L.Che., J-F.Dar.**; East Asia sample contribution:** Y.K., M.K., X.Z., H.C., N.Y.I., A.K.Y.F., F.C.F.I., A.M., N.H., K.O., S.N., J.G., V.E-P., K.H.L., T.I.**; South America sample contribution:** C.Dalm., C.E.A-B., M.A.C.B., N.O., T.J-C., C.Muc., C.Cue., L.Cam., P.Sol., D.G.P., S.K., L.I.B., J.O-R., A.G.C.M.,M.F.M., R.Par., G.A., L.A.d.M., M.A.R.S., B.d.M.V., M.T.G.C., B.Ang., S.G., M.V.C., R.A., P.O., A.Sla., C.G-B., C.A., P.F., E.N.d.M., L.M., H.A., A.Rui., A.Ram**; Core writing group**: A.N, B.G-B., J-C.L.

## Competing Interests Statement

S.V.D.L is recipient of ABOARD funding, which is a public-private partnership funded by from ZonMW (#73305095007) and Health∼Holland, Topsector Life Sciences & Health (PPP-allowance; #LSHM20106). C.C. has received research support from GSK and EISAI. The study’s funders had no role in the collection, analysis, or interpretation of data; in the writing of the report; or in the decision to submit the paper for publication. C.C. is an advisory board member for Vivid Genomics and Circular Genomics and owns stocks. L.M.P. received personal fees from Biogen for consulting activities unrelated to the submitted work T.G. received consulting fees from AbbVie, Alector, Anavex, Biogen, Cogthera, Eli Lilly, Functional Neuromodulation, Grifols, Iqvia, Janssen, Noselab, Novo Nordisk, NuiCare, Orphanzyme, Roche Diagnostics, Roche Pharma, UCB, and Vivoryon; lecture fees from Biogen, Eisai, Grifols, Medical Tribune, Novo Nordisk, Roche Pharma, Schwabe, and Synlab; and has received grants to his institution from Biogen, Eisai, and Roche Diagnostics.

O.A.A. is a consultant to Cortechs and Precision Health AS, and has received speaker’s honorarium from Lundbeck, Sunovion, Otsuka and Janssen.

## METHODS Online

### Sample and variant quality controls

To ensure that the βs were completely independent of the summary statistics, all samples from ADGC, CHARGE and FinnGen GWASs were filtered out. In addition, Sample overlap was systematically assessed and there was no sample overlap between any of the non-US studies analyzed. Overlap between ADSP and MVP is likely to be negligible (no more than a few cases). For biomarker analysis, there is overlap of 460 samples between the American samples used in the biomarker analyses and the ADGC (which is included in the summary statistics we used to generate the β for the PGS^ALZ^). However, this overlap is limited (less than 2.5%) and in addition, we only analyzed in these samples the association of PGS^ALZ^ with quantitative traits (p-tau, tau and Aβ42 CSF concentrations), limiting the risk of inflated results. After it met the conventional GWAS gold standard of sample quality control, each sample was included in the analyses^16^. If a discordance in the variant dose or covariate or a discordance between imputed and genotyped (if available) APOE status was observed, the sample was discarded. After the quality control, a demographic description of each study is shown in Supplementary Table 1^38^. Genotyped variants had to meet the gold standard GWAS variant quality control^16^. All the studies including genotyping data were imputed with the TOPmed reference panel^39,38^. If the variants were imputed, those with a Rsq below 0.3 were excluded. For whole-genome sequencing data, only variants passing the corresponding quality control were selected (see the supplementary information for the ADSP and China samples) (Supplementary Table 2). The global ancestry of each individual in the ADSP samples was determined with SNPweights v.2.1^40^ using a set of ancestry-weighted variants computed on reference populations from the 1000 Genomes Project as in^41^. By applying a global ancestry percentage cutoff of >75%, the samples were assigned to the different ancestry populations. The ancestry of MVP participants was determined using the harmonized ancestry and race/ethnicity (HARE) method^42^, which is similar to other genotype-based ancestry calling methods, except that concordance is checked between self-report and genetically inferred ancestry, and those with discrepant ancestry groups are not assigned a HARE category. Within-group PCs for ancestry were computed using FlashPCA2^43^.

### European mega-analysis methods

For the mega-analysis of European countries, we merged samples from five datasets: EADB-core, GERAD, EADI, Demgene, and Bonn. To adjust for the population structure, we computed principal components using the following procedure. From the list of 146,705 variants used in the principal component analysis of EADB-core^43^, we extracted TOPMed imputed variants having an imputation quality ≥0.9 in each dataset; this resulted in 91,353 variants. Next, we set a genotype to “missing” if none of the genotype probabilities was higher than 0.8. Lastly, we merged all the datasets and removed variants for which the proportion of missing genotypes was above 0.02. Ultimately, 90,471 variants were fed into the principal component analysis (performed with FlashPCA2). The analyses were adjusted for the first 14 principal components, the genotyping chip, and the center.

### PGS and PRS Computations

The equation used to calculate the PGSs and the coPRSs is as follow:

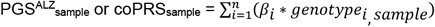

where the PGS^ALZ^/PRS is the sum per sample, of the product of the variant *i* effect size *βi*, extracted from GWAS summary statistics, and the number of risk alleles of this variant *i*, either as a dosage or as a genotype.

The PGS^ALZ^ includes the 83 independent signals associated with AD^13^, listed in Supplementary Table 1. We also calculated another PGS^ALZ^ combining the same 83 independent signals and the two SNPs encoding the APOE ε2 (rs7412) and APOE ε4 alleles (rs429358). The PGS^APOE^ includes only these two last SNPs. The stage I meta-analysis of EADB studies^13^ without the UK Biobank samples, contained 39,106 clinically diagnosed AD cases versus 25,392 in the stage II meta-analysis (including the ADGC, CHARGE and FinnGen data)^13^. To respect the independence between the samples and the GWAS summary statistics, in the PGS analyses, the European summary statistics used are from stage II or (for APOE variants only). In the PGS^ALZ^/PRS analyses adjusted for difference in distribution between populations, the European more powerful summary statistics, *i*.*e*. stage I meta-analysis of EADB were preferred.

The PGS^ALZ+^ score was developed to include additional SNPs within the GWAS-defined loci to capture more genetic information in non-European-ancestry populations. Firstly, each locus’s “start and end positions” (as specified in the GRCh38 assembly) were defined manually by looking at the regional plots and extracting (i) recombination rate peak positions, (ii) starts/ends of chromosomes, (iii) specific variant positions, or (iv) start/end positions of regions containing no variants. Next, insertions and deletions were excluded. Variants that were non ambiguous (i.e. A/T or C/G) and present in the 1000 Genomes Phase 3 data (1000GP3) and had an imputation quality above 0.3 in the EADB-core TOPMed imputations, were selected. To extract information of these variants in non-European-ancestry populations, we used the summary statistics generated by Lake et al., Shigemizu et al. and Kunkle et al. to represent Latino American, East Asian and African-American ancestries, respectively^27,28,26^. Since these summary statistics were based on the GRCh37 assembly, we lifted their positions and alleles in the GRCh38 assembly by using the Picard LiftoverVcf tool (v2.27.5) and restricting the process to variants with a minor allele frequency above 0.01. In order to remove variants in LD with the sentinel variant of each locus, we computed the LD between each sentinel variant and all other variants within the locus by using the 1000GP3 data restricted to samples representing European ancestries (EUR super-population), Latino American ancestries (AMR super-population plus IBS population), Japanese ancestries (JPT population) and African-American ancestries (AFR super-population). Since one of the sentinel variant (chr9:104903697:C:G) was not present in the 1000GP3 data, we replaced it with a proxy variant (chr9:104903754:G:GC, R^2^=1 in the EUR super-population). In each set of summary statistics, we removed variants with R^2^>0.1 in either the European summary statistics or the summary statistics for the corresponding ancestry. Lastly, we performed a clumping procedure on the remaining variants in each of the three ancestries by using plink v1.9, a p-value threshold of 1×10^-3^, a R^2^ of 0.05 (as estimated in the corresponding 1000GP3 data samples, as described above), and a distance of 1Mb. For the PGS^ALZ+^, this led us to select 30, 13 and 47 variants (in addition to the initial 85 PGS variants) for the Latino American, East Asian and African-American ancestries, respectively.

PRS-CSx^20,44^ was, by the time of analysis, one of the most performing cross-ancestry PRS model method^45,46^, without a validation dataset and using GWAS summary statistics. With a Bayesian high-dimensional regression framework model based on continuous shrinkage priors, the variant effect sizes were adaptively re-estimated, by coupling cross-ancestry GWAS summary statistics^13,26,27,28^, and external ancestry-matched allele frequencies and local Linkage Disequilibrium structure, according to a global shrinkage parameter. This global shrinkage parameter provides the sparseness of the genetic architecture of AD, by avoiding over-shrinkage of true signals and by shrinking noisy signals. This sparseness was modeled for the values of 1, 10^-2^, 10^-4^, 10^-5^,10^-6^, 10^-7^ and 10^-8^, with the --meta option and the Strawderman–Berger prior default parameters (*a* = 1 and *b* = 0.5). The initial 1,297 432 variants present in the 1000 Genomes reference panel, were lifted over in GRCh38. Then, new ancestry-specific or joint-ancestry effect size estimates were obtained with PRS-CSx, leading to a maximal number of 1,292 532 variants in the joint-ancestry summary statistics, potentially included in the PRS computations. The coPRSs were computed per chromosome, with 1-joint-ancestry and 2-European ancestry and 3-ancestry-specific PRS-CSx-effect size estimates, using PLINK (v.2.0.a) software^47^ and its --score option, then summed across all chromosomes.

### Adjustment for difference in PGS^ALZ^/PRS distribution across populations

To account for population structure, PRS_raw_ and PGS^ALZ^_raw_ were adjusted for difference in distributions across populations^48^. The adjustment was performed with a selection of 84,035 variants, common to all studies, independent and well-imputed variants (R > 0.8). With this list of variants, FlashPCA2 projected the samples into the 1000G Phase 3 PC-space and calculated the projected PCs. For each study, the raw score was fit in a linear model, in controls, according to the 5 first projected PCs. This model was used to compute a predicted score in all the samples. The resulting adjusted score was the difference between the raw score and the predicted score.

### Statistical analyses

The different scores (PGS or coPRS) were standardized in a standard normal distribution, using the mean and the standard deviation, calculated over all the samples. The associations between AD status and the different scores were tested in logistic regression, named according to the score and the covariates. The name “ALZinclAPOE” was attributed, if the score included variants in the APOE region (from 43Mb to 47Mb). The other covariates included age, sex, in addition to covariates specific to each study (Supplementary Table 2).

- Model PGS^ALZ^: AD ∼ PGS^ALZ^ + COV
- Model PGS^ALZ^: AD ∼ PGS^ALZ^ + COV + the count of APOE ε2 alleles + the count of APOE ε4 alleles (when adjusted on *APOE*)
- Model PRS: AD ∼ PRS + COV
- Model PRS: AD ∼ PRS + COV + the count of APOE ε2 alleles + the count of APOE ε4 alleles (when adjusted on *APOE*)
- Model PRS^ALZinclAPOE^: AD ∼ PRS^ALZinclAPOE^ + COV

To estimate the proportion of phenotypic variance explained by the variance of the score, we computed the Nagelkerke’s *Pseudo*-R^2^_Full_ using the function Nagelkerke implemented in the package *rcompanion* in R^49,50^. A *Pseudo*-R^2^_Null_ was computed including only the covariates. The adjusted *Pseudo*-R^2^ is the difference between *Pseudo*-R^2^_Full_ and the tied *Pseudo*-R^2^_Null_. This adjusted Pseudo-R^2^ corresponds to the phenotypic variance explained by the genetic score only. The adjusted *Pseudo*-R^2^ was also transformed into a liability scale, for ascertained case-control studies^51^, using a prevalence value of 0.15.

We chose a prevalence of 15%, which we consider to be consistent for populations with an average age over 75. However, this prevalence is different in multi-ethnic populations of the same average age. Furthermore, the AD prevalence increases with age, so the genetic liability is not homogeneous across all age groups. AD heritability cannot be expressed as a single number because it depends on the ages of the cases and controls^52^.

### Quantile or percentile analyses

Depending on the value of the corresponding PGS^ALZ^, the samples were classified into the reference group or into one of the test groups. In the mega-analysis, the reference group corresponded to the 40-60% percentile and was tested across different percentiles (0-2%, 2-5%, 5-10%, 10-20%, 20-40%, 60-80%, 80-90%, 95-98%, and 98-100%). In the *APOE*-stratified analysis and in the multi-ancestry analyses, the reference group was defined as the 40-60% percentile and was tested across different quintiles (0-20%, 20-40%, 60-80%, 80-100%). The multi ancestry analyses were performed for each population and then meta-analyzed per genetic ancestry using the inverse variance method, as implemented in METAL^53^. It should be noted that the Indian, North African and sub-Saharan African populations were excluded because of their small sample size.

- Model PGS^ALZ^: AD ∼ Group_0/1_(PGS^ALZ^) + COV
- Model PGS^ALZ^: AD ∼ Group_0/1_(PGS^ALZ^) + COV + number of APOE ε2 alleles + number of APOE ε4 alleles (when adjusted on *APOE*)
- Model PGS^ALZinclAPOE^: AD ∼ Group_0/1_(PGS^ALZinclAPOE^) + COV

## Data Availability

1000GP3:

http://ftp.1000genomes.ebi.ac.uk/vol1/ftp/data_collections/1000_genomes_project/release/20190312_biallelic_SNV_and_INDEL/)

ADSP: https://dss.niagads.org/datasets/ng00067/

## Code availability

The sets of scripts are available upon request.

---

**Extended Figure 1:**
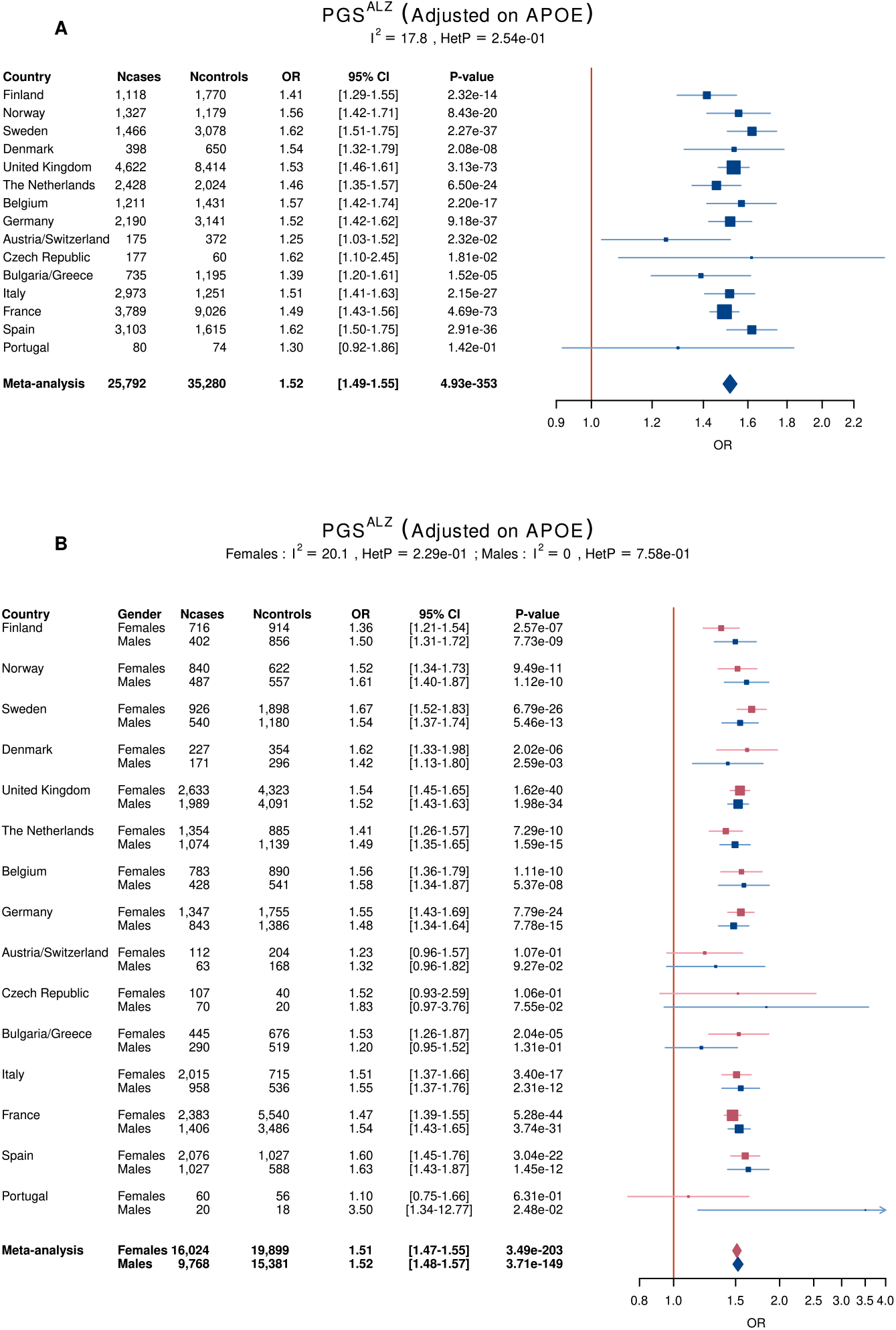
Association of PGS^ALZ^ with the risk of developing AD **(A)** in 17 European countries and **(B)** in Men and Women. Ncases, number of cases; Ncontrols, number of controls; OR, Odds ratio per Standard deviation. The lines in the Forest plots indicate the 95% confidence interval for the ORs.

**Extended Figure 2:**
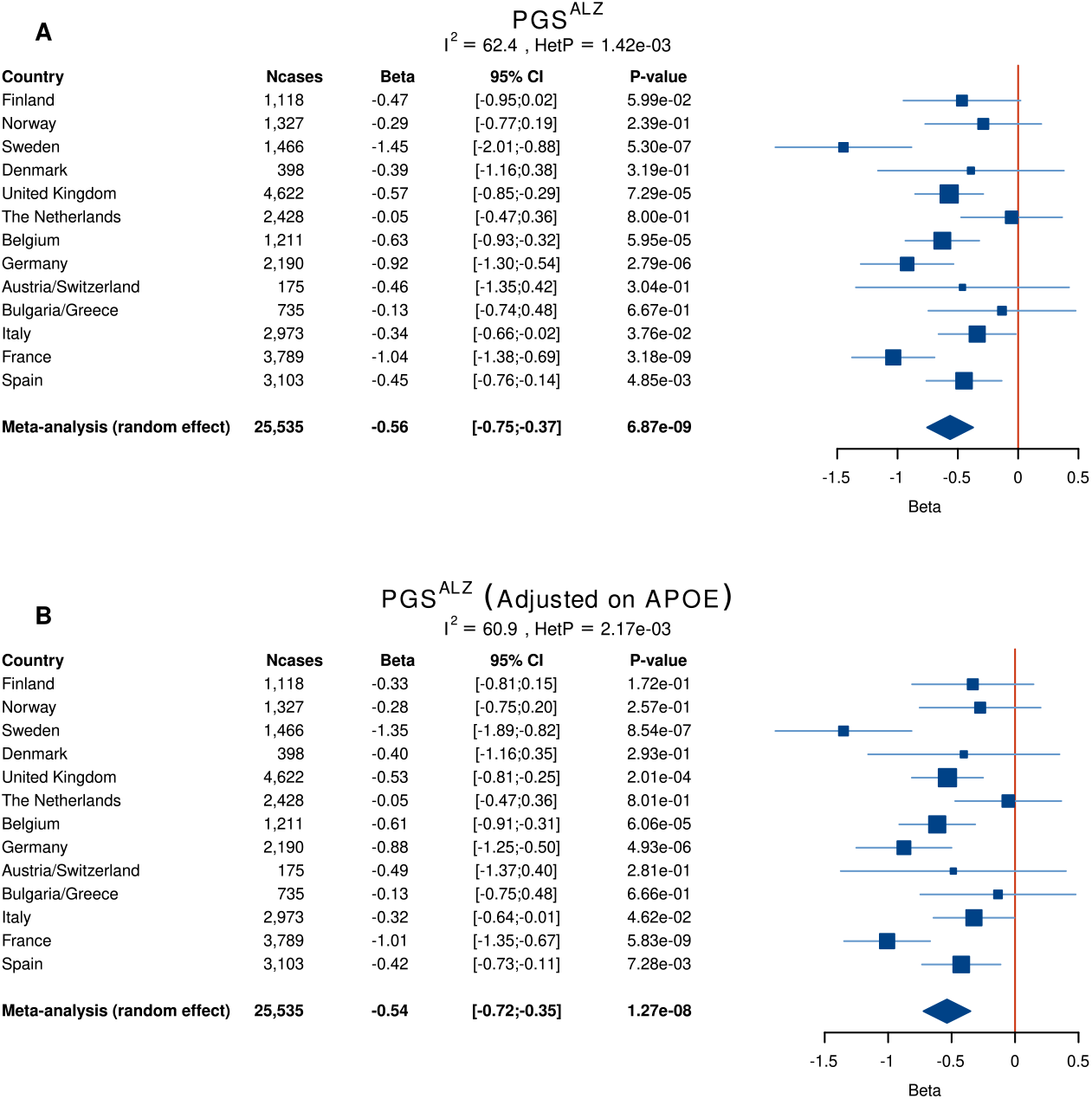
Associations between (A) PGS^ALZ^ or (B) PGS^ALZ^ adjusted for *APOE* and age at onset of AD in European countries. N_cases_, the number of cases. Since HetP <0.05, the random effect is shown for the meta-analysis results.

**Extended Figure 3:**
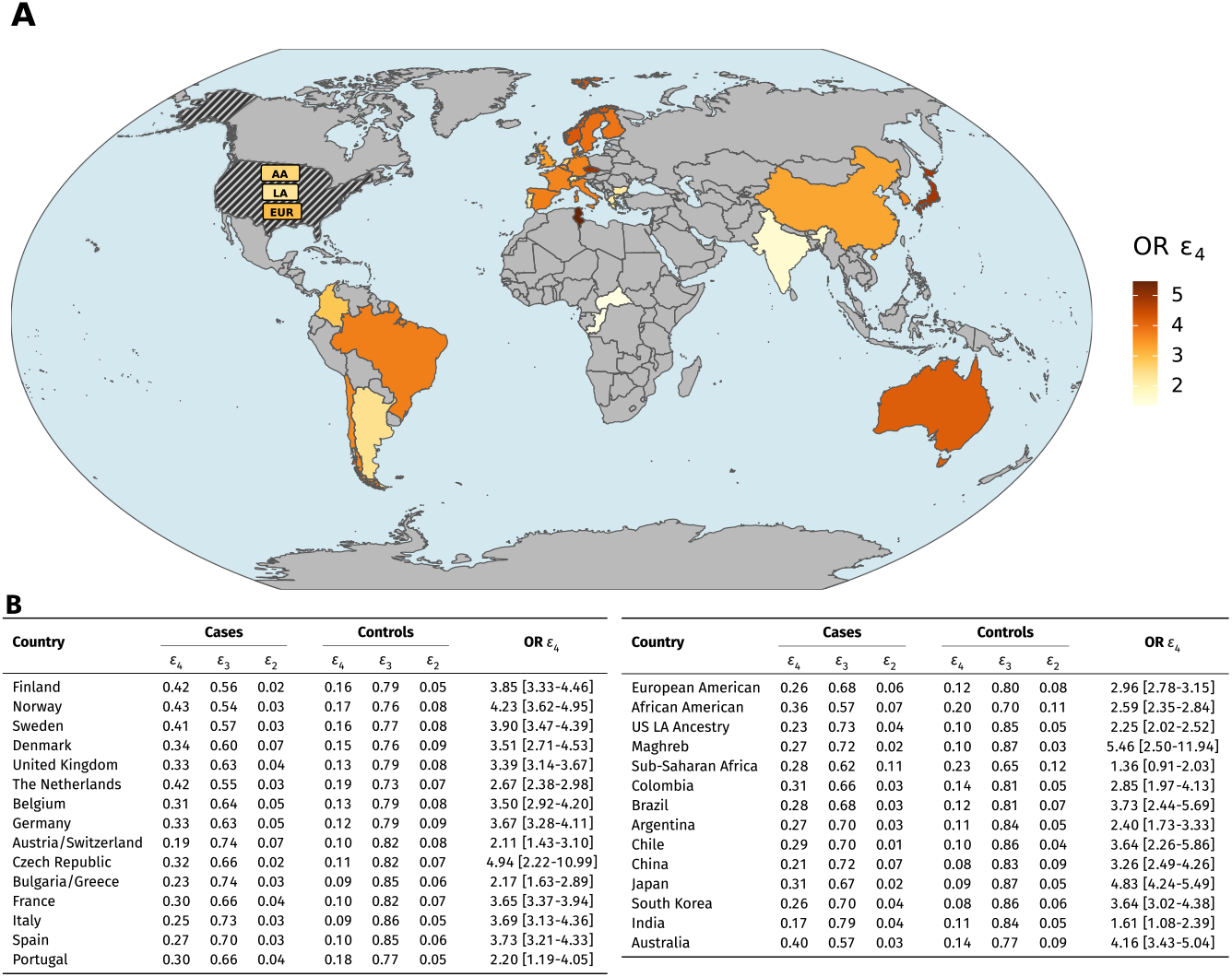
Distribution and association of *APOE* ε2/ε3/ε4 alleles with AD risk worldwide. **(A)** World map showing the populations analyzed. A color gradient indicates the strength of the association between *APOE* ε2/ε3/ε4 alleles and the risk of developing AD in different countries **(B)** frequencies of *APOE* ε2/ε3/ε4 alleles in case and controls as well association of *APOE* ε4 alleles with the risk of developing AD in different countries.

**Extended Figure 4:**
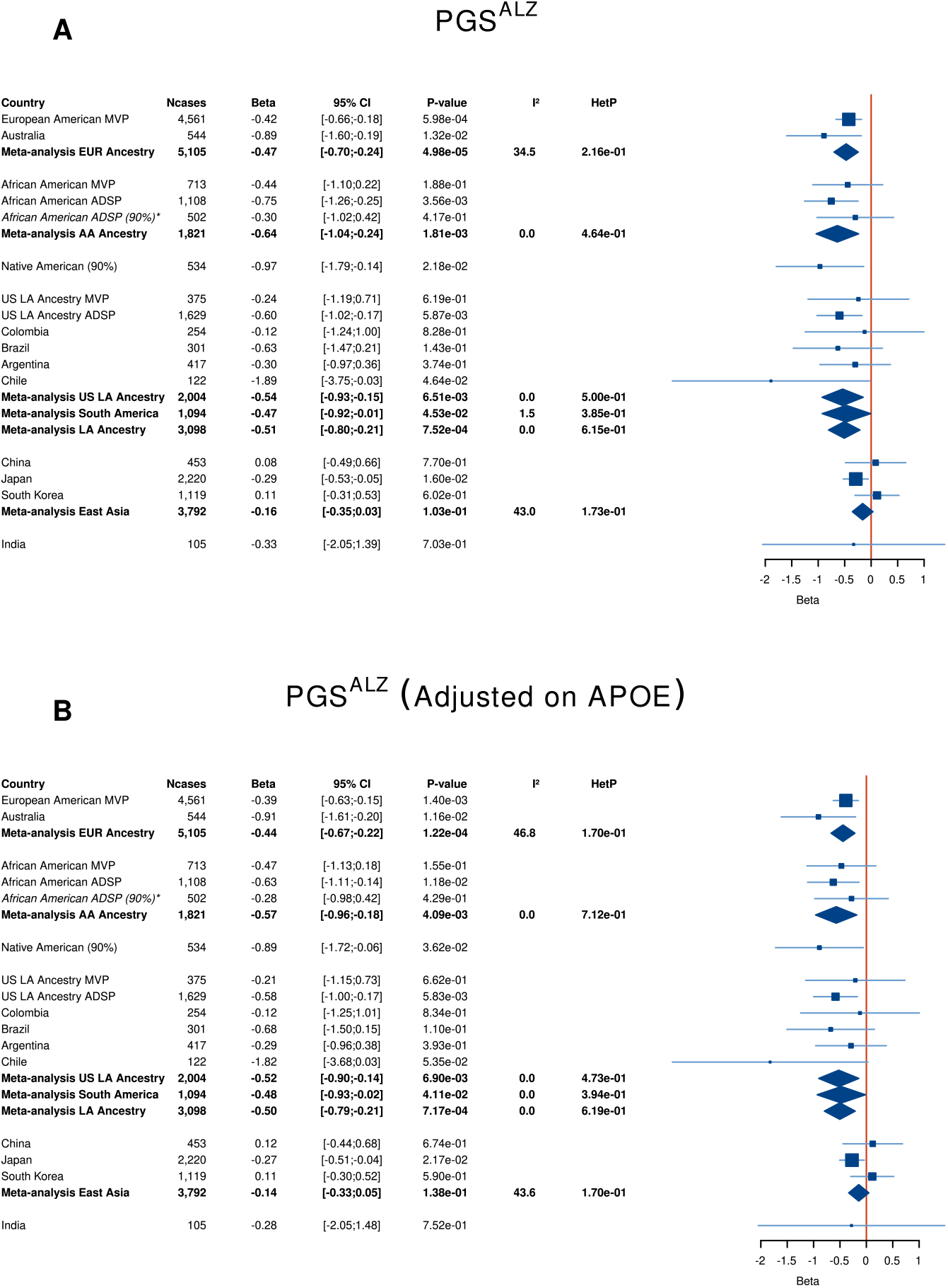
Association between (A) PGS^ALZ^ or (B) PGS^ALZ^ (adjusted for *APOE*) and age at onset of AD in multi-ancestry populations. N_cases_, number of cases. The African-American-ancestry meta-analysis (more than 75% of the population with African-American ancestry) included the MVP and ADSP datasets. The East Asia meta-analysis included datasets from China, Korea, and Japan. The Latin American (LA) ancestry (self-reporting) meta-analysis included the MMVP and ADSP datasets. The South America meta-analysis included the datasets from Argentina, Brazil, Chile, and Colombia. * not used in the meta-analysis.

## Notes

### Funding Statement

The work for this manuscript was further supported by the CoSTREAM project (www.costream.eu) and funding from the European Union Horizon 2020 research and innovation programme under grant agreement No 667375. This work is also funded by la fondation pour la recherche medicale (FRM) (EQU202003010147) Italian Ministry of Health (Ricerca Corrente); Ministero dell'Istruzione, del l'Universita e della Ricerca MIUR project (Dipartimenti di Eccellenza 2018 2022) to Department of Neuroscience (Rita Levi Montalcini), University of Torino (IR), and AIRAlzh Onlus-ANCC-COOP (SB); Partly supported by (Ministero della Salute), I.R.C.C.S. Research Program, Ricerca Corrente 2018-2020, Linea n. 2 (Meccanismi genetici, predizione e terapie innovative delle malattie complesse) and by the (5 x 1000) voluntary contribution to the Fondazione I.R.C.C.S. Ospedale (Casa Sollievo della Sofferenza); and RF-2018-12366665, Fondi per la ricerca 2019 (Sandro Sorbi) and Italian Ministry of Health, Ricerca Corrente (RG, IRCCS Istituto Centro San Giovanni di Dio Fatebenefratelli Brescia); Copenhagen General Population Study (CGPS): We thank staff and participants of the CGPS for their important contributions. Karolinska Institutet AD cohort: Dr. C.G. and co-authors of the Karolinska Institutet AD cohort report grants from Swedish Research Council (VR) 2015-02926, 2018-02754, 2015-06799, Swedish Alzheimer Foundation, Stockholm County Council ALF and resarch school, Karolinska Institutet StratNeuro, Swedish Demensfonden, and Swedish brain foundation, during the conduct of the study. ADGEN: This work was supported by Academy of Finland (grant numbers 307866); Sigrid Juselius Foundation; the Strategic Neuroscience Funding of the University of Eastern Finland; EADB project in the JPNDCO-FUND program (grant number 301220). CBAS: Supported by the Ministry of Health, Czech Republic―conceptual development of research organization, University Hospital Motol, Prague, Czech Republic Grant No. 00064203; Institutional Support of Excellence 2. LF UK Grant No. 699012; and The project National Institute for Neurological Research (Programme EXCELES, ID Project No. LX22NPO5107) - Funded by the European Union Next Generation EU. CNRMAJ-Rouen: This study received fundings from the Centre National de Reference Malades Alzheimer Jeunes (CNRMAJ). The Finnish Geriatric Intervention Study for the Prevention of Cognitive Impairment and Disability (FINGER) data collection was supported by grants from the Academy of Finland, La Carita Foundation, Juho Vainio Foundation, Novo Nordisk Foundation, Finnish Social Insurance Institution, Ministry of Education and Culture Research Grants, Yrj o Jahnsson Foundation, Finnish Cultural Foundation South Osthrobothnia Regional Fund, and EVO/State Research Funding grants of University Hospitals of Kuopio, Oulu and Turku, Seinajoki Central Hospital and Oulu City Hospital, Alzheimer Research & Prevention Foundation USA, AXA Research Fund, Knut and Alice Wallenberg Foundation Sweden, Center for Innovative Medicine (CIMED) at Karolinska Institutet Sweden, and Stiftelsen Stockholms sjukhem Sweden. FINGER cohort genotyping was funded by EADB project in the JPND CO-FUND (grant number 301220). Research at the Belgian EADB site is funded in part by the Alzheimer Research Foundation (SAO-FRA), The Research Foundation Flanders (FWO), and the University of Antwerp Research Fund. FK is supported by a fellowship of the University of Antwerp Research Fund. SNAC-K is financially supported by the Swedish Ministry of Health and Social Affairs, the participating County Councils and Municipalities, and the Swedish Research Council. BDR Bristol: We would like to thank the South West Dementia Brain Bank (SWDBB) for providing brain tissue for this study. The SWDBB is part of the Brains for Dementia Research programme, jointly funded by Alzheimer Research UK and Alzheimer Society and is supported by BRACE (Bristol Research into Alzheimer and Care of the Elderly) and the Medical Research Council. BDR Manchester: We would like to thank the Manchester Brain Bankfor providing brain tissue for this study. The Manchester Brain Bank is part of the Brains for Dementia Research programme, jointly funded by Alzheimer Research UK and Alzheimer Society. BDR KCL: Human post-mortem tissue was provided by the London Neurodegenerative Diseases Brain Bank which receives funding from the UK Medical Research Council and as part of the Brains for Dementia Research programme, jointly funded by Alzheimer Research UK and the Alzheimer Society. The CFAS Wales study was funded by the ESRC (RES-060-25-0060) and HEFCW as (Maintaining function and well-being in later life: a longitudinal cohort study). We are grateful to the NISCHR Clinical Research Centre for their assistance in tracing participants and in interviewing and in collecting blood samples, and to general practices in the study areas for their cooperation. MRC: We thank all individuals who participated in this study. Cardiff University was supported by the Alzheimer Society (AS; grant RF014/164) and the Medical Research Council (MRC; grants G0801418/1, MR/K013041/1, MR/L023784/1) (R.S. is an AS Research Fellow). Cardiff University was also supported by the European Joint Programme for Neurodegenerative Disease (JPND; grant MR/L501517/1), Alzheimer Research UK (ARUK; grant ARUK-PG2014-1), the Welsh Assembly Government (grant SGR544:CADR), Brain for dementia Research and a donation from the Moondance Charitable Foundation. Cardiff University acknowledges the support of the UK Dementia Research Institute, of which J.W. is an associate director. Cambridge University acknowledges support from the MRC. Patient recruitment for the MRC Prion Unit/UCL Department of Neurodegenerative Disease collection was supported by the UCLH/UCL Biomedical Centre and NIHR Queen Square Dementia Biomedical Research Unit. The University of Southampton acknowledges support from the AS. King College London was supported by the NIHR Biomedical Research Centre for Mental Health and the Biomedical Research Unit for Dementia at the South London and Maudsley NHS Foundation Trust and by King College London and the MRC. ARUK and the Big Lottery Fund provided support to Nottingham University. A.Ram. : Part of the work was funded by the JPND EADB grant (German Federal Ministry of Education and Research (BMBF) grant: 01ED1619A). A. Ram. is also supported by the German Research Foundation (DFG) grants Nr: RA 1971/6-1, RA1971/7-1, and RA 1971/8-1. German Study on Ageing, Cognition and Dementia in Primary Care Patients (AgeCoDe): This study/publication is part of the German Research Network on Dementia (KND), the German Research Network on Degenerative Dementia (KNDD; German Study on Ageing, Cognition and Dementia in Primary Care Patients; AgeCoDe), and the Health Service Research Initiative (Study on Needs, health service use, costs and health-related quality of life in a large sample of oldestold primary care patients (85+ AgeQualiDe)) and was funded by the German Federal Ministry of Education and Research (grants KND: 01GI0102, 01GI0420, 01GI0422, 01GI0423, 01GI0429, 01GI0431, 01GI0433, 01GI0434; grants KNDD: 01GI0710, 01GI0711, 01GI0712, 01GI0713, 01GI0714, 01GI0715, 01GI0716; grants Health Service Research Initiative: 01GY1322A, 01GY1322B, 01GY1322C, 01GY1322D, 01GY1322E, 01GY1322F, 01GY1322G). VITA study: The support of the Ludwig Boltzmann Society and the AFI Germany have supported the VITA study. The former VITA study group should be acknowledged: W. Danielczyk, G. Gatterer, K Jellinger, S Jugwirth, KH Tragl, S Zehetmayer. Vogel Study: This work was financed by a research grant of the (Vogelstiftung Dr. Eckernkamp). HELIAD study: This study was supported by the grants: IIRG-09-133014 from the Alzheimer Association, 189 10276/8/9/2011 from the ESPA-EU program Excellence Grant (ARISTEIA) and the ΔΥ2β/οικ.51657/14.4.2009 of the Ministry for Health and Social Solidarity (Greece). Biobank Department of Psychiatry, UMG: Prof. Jens Wiltfang is supported by an Ilidio Pinho professorship and iBiMED (UID/BIM/04501/2013), and FCT project PTDC/DTP_PIC/5587/2014 at the University of Aveiro, Portugal. Lausanne study: This work was supported by grants from the Swiss National Research Foundation (SNF 320030_141179). PAGES study: Harald Hampel is an employee of Eisai Inc. During part of this work he was supported by the AXA Research Fund, the (Fondation partenariale Sorbonne Universite) and the (Fondation pour la Recherche sur Alzheimer), Paris, France. Mannheim, Germany Biobank: Department of geriatric Psychiatry, Central Institute for Mental Health, Mannheim, University of Heidelberg, Germany. Genotyping for the Swedish Twin Studies of Aging was supported by NIH/NIA grant R01 AG037985. Genotyping in TwinGene was supported by NIH/NIDDK U01 DK066134. WvdF is recipient of Joint Programming for Neurodegenerative Diseases (JPND) grants PERADES (ANR-13-JPRF-0001) and EADB (733051061). Gothenburg Birth Cohort (GBC) Studies: We would like to thank UCL Genomics for performing the genotyping analyses. The studies were supported by The Stena Foundation, The Swedish Research Council (2015-02830, 2013-8717), The Swedish Research Council for Health, Working Life and Wellfare (2013-1202, 2005-0762, 2008-1210, 2013-2300, 2013-2496, 2013-0475), The Brain Foundation, Sahlgrenska University Hospital (ALF), The Alzheimer Association (IIRG-03-6168), The Alzheimer Association Zenith Award (ZEN-01-3151), Eivind och Elsa K:son Sylvans Stiftelse, The Swedish Alzheimer Foundation. Clinical AD, Sweden: We would like to thank UCL Genomics for performing the genotyping analyses. Barcelona Brain Biobank: Brain Donors of the Neurological Tissue Bank of the Biobanc-Hospital Clinic-IDIBAPS and their families for their generosity. Hospital Clinic de Barcelona Spanish Ministry of Economy and Competitiveness-Instituto de Salud Carlos III and Fondo Europeo de Desarrollo Regional (FEDER), Union Europea, (Una manera de hacer Europa) grants (PI16/0235 to Dr. R. Sanchez-Valle and PI17/00670 to Dr. A.Antonelli). AA is funded by Departament de Salut de la Generalitat de Catalunya, PERIS 2016-2020 (SLT002/16/00329). Work at JP-T laboratory was possible thanks to funding from Ciberned and generous gifts from Consuelo Cervera Yuste and Juan Manuel Moreno Cervera. Sydney Memory and Ageing Study (Sydney MAS): We gratefully acknowledge and thank the following for their contributions to Sydney MAS: participants, their supporters and the Sydney MAS Research Team (current and former staff and students). Funding was awarded from the Australian National Health and Medical Research Council (NHMRC) Program Grants (350833, 568969, 109308). This work was supported by InnoMed (Innovative Medicines in Europe), an integrated project funded by the European Union of the Sixth Framework program priority (FP6-2004-LIFESCIHEALTH-5). Oviedo: This work was partly supported by Grant from Fondo de Investigaciones Sanitarias-Fondos FEDER EuropeanUnion to V.A. PI15/00878. Project MinE: The ProjectMinE study was supported by the ALS Foundation Netherlands and the MND association (UK) (Project MinE, www.projectmine.com). The SPIN cohort: We are indebted to patients and their families for their participation in the (Sant Pau Initiative on Neurodegeneration cohort), at the Sant Pau Hospital (Barcelona). This is a multimodal research cohort for biomarker discovery and validation that is partially funded by Generalitat de Catalunya (2017 SGR 547 to JC), as well as from the Institute of Health Carlos III-Subdireccion General de Evaluacion and the Fondo Europeo de Desarrollo Regional (FEDER-(Una manera de Hacer Europa)) (grants PI11/02526, PI14/01126, and PI17/01019 to JF; PI17/01895 to AL), and the Centro de Investigacion Biomedica en Red Enfermedades Neurodegenerativas programme (Program 1, Alzheimer Disease to AL). We would also like to thank the Fundacio Bancaria Obra Social La Caixa (DABNI project) to JF and AL; and Fundacion BBVA (to AL), for their support in funding this follow-up study. Adolfo Lopez de Munain is supported by Fundacion Salud 2000 (PI2013156), CIBERNED and Diputacion Foral de Gipuzkoa (Exp.114/17). P.S.J. is supported by CIBERNED and Carlos III Institute of Health, Spain (PI08/0139, PI12/02288, and PI16/01652, PI20/01011), jointly funded by Fondo Europeo de Desarrollo Regional (FEDER), Union Europea, (Una manera de hacer Europa). We thank Biobanco Valdecilla for their support. Dr. Fermin Moreno is supported by the Tau Consortium and has received funding from the Carlos III Health Institute (PI19/01637). Amsterdam dementia Cohort (ADC): Research of the Alzheimer center Amsterdam is part of the neurodegeneration research program of Amsterdam Neuroscience. The AlzheimerCenter Amsterdam is supported by Stichting Alzheimer Nederland and Stichting VUmc fonds. The clinical database structure was developed with funding from Stichting Dioraphte. Genotyping of the Dutch case-control samples was performed in the context of EADB (European Alzheimer&Dementia biobank) funded by the JPco-fuND FP-829-029 (ZonMW project number #733051061). This research is performed by using data from the Parelsnoer Institute an initiative of the Dutch Federation of University Medical Centres (www.parelsnoer.org). 100-Plus study: We are grateful for the collaborative efforts of all participating centenarians and their family members and/or relations. We thank the Netherlands Brain Bank for supplying DNA for genotyping. This work was supported by Stichting AlzheimerNederland (WE09.2014-03), Stichting Diorapthe, Horstingstuit foundation, Memorabel (ZonMW project number #733050814, #733050512) and Stichting VUmcFonds. Additional support for EADB cohorts: WF, SL, HH are recipients of ABOARD, a public-private partnership receiving funding from ZonMW (#73305095007) and Health∽Holland, Topsector Life Sciences & Health (PPP-allowance; #LSHM20106). The DELCODE study was funded by the German Center for Neurodegenerative Diseases (Deutsches Zentrum fur Neurodegenerative Erkrankungen (DZNE)), reference number BN012.
Gra@ce. We would like to thank patients and controls who participated in this project. The Genome Research @ Ace Alzheimer Center Barcelona project (GR@ACE) is supported by Grifols SA, Fundacion bancaria (La Caixa), Ace Alzheimer Center Barcelona and CIBERNED. Ace Alzheimer Center Barcelona is one of the participating centers of the Dementia Genetics Spanish Consortium (DEGESCO). AR and MB receive support from the European Union / EFPIA Innovative Medicines Initiative joint undertaking ADAPTED and MOPEAD projects (grant numbers 115975 and 115985, respectively). MB and AR are also supported by national grants PI13/02434, PI16/01861, PI17/01474, PI19/01240, PI19/01301 and PI22/01403. Accion Estrategica en Salud is integrated into the Spanish National R+D+I Plan and funded by ISCIII Subdireccion General de Evaluacion and the Fondo Europeo de Desarrollo Regional (FEDER (Una manera de hacer Europa)). AR is also funded by JPco-fuND-2 (Multinational research projects on Personalized Medicine for Neurodegenerative Diseases), PREADAPT project (ISCIII grant: AC19/00097), EURONANOMED III Joint Transnational call for proposals (2017) for European Innovative Research & Technological Development Projects in Nanomedicine (ISCIII grant: AC17/00100), the ISCIII national grant PMP22/00022, funded by the European Union (NextGenerationEU), The support of CIBERNED (ISCIII) under the grants CB06/05/2004 and CB18/05/00010. The support from the ADAPTED and MOPEAD projects, European Union/EFPIA Innovative Medicines Initiative Joint (grant numbers 115975 and 115985, respectively); from PREADAPT project, Joint Program for Neurodegenerative Diseases (JPND) grant N AC19/00097; from HARPONE project, Agency for Innovation and Entrepreneurship (VLAIO) grant N PR067/21, Janssen. And DESCARTES project funded by German Research Foundation (DFG).
I.dR. is supported by a national grant from the Instituto de Salud Carlos III FI20/00215. Ace Alzheimer Center Barcelona research receives support from Roche, Janssen, Life Molecular Imaging, Araclon Biotech, Alkahest, Laboratorio de Analisis Echevarne, and IrsiCaixa. Some control samples and data from patients included in this study were provided in part by the National DNA Bank Carlos III (www.bancoadn.org, University of Salamanca, Spain) and Hospital Universitario Virgen de Valme (Sevilla, Spain); they were processed following standard operating procedures with the appropriate approval of the Ethical and Scientific Committee.
EADI. This work has been developed and supported by the LABEX (laboratory of excellence program investment for the future) DISTALZ grant (Development of Innovative Strategies for a Transdisciplinary approach to ALZheimer disease) including funding from MEL (Metropole europenne de Lille), ERDF (European Regional Development Fund) and Conseil Regional Nord Pas de Calais. This work was supported by INSERM, the National Foundation for Alzheimer disease and related disorders, the Institut Pasteur de Lille and the Centre National de Recherche en Genomique Humaine, CEA, the JPND PERADES, the Laboratory of Excellence GENMED (Medical Genomics) grant no. ANR-10-LABX-0013 managed by the National Research Agency (ANR) part of the Investment for the Future program, and the FP7 AgedBrainSysBio. The Three-City Study was performed as part of collaboration between the Institut National de la Sante et de la Recherche Medicale (Inserm), the Victor Segalen Bordeaux II University and Sanofi-Synthelabo. The Fondation pour la Recherche Medicale funded the preparation and initiation of the study. The 3C Study was also funded by the Caisse Nationale Maladie des Travailleurs Salaries, Direction Generale de la Sante, MGEN, Institut de la Longevite, Agence Francaise de Securite Sanitaire des Produits de Sante, the Aquitaine and Bourgogne Regional Councils, Agence Nationale de la Recherche, ANR supported the COGINUT and COVADIS projects. Fondation de France and the joint French Ministry of Research/INSERM (Cohortes et collections de donnes biologiques) programme. Lille Genopole received an unconditional grant from Eisai. The Three-city biological bank was developed and maintained by the laboratory for genomic analysis LAG-BRC - Institut Pasteur de Lille.
GERAD/PERADES. We thank all individuals who participated in this study. Cardiff University was supported by the Wellcome Trust, Alzheimer Society (AS; grant RF014/164), the Medical Research Council (MRC; grants G0801418/1, MR/K013041/1, MR/L023784/1), the European Joint Programme for Neurodegenerative Disease (JPND, grant MR/L501517/1), Alzheimer Research UK (ARUK, grant ARUK-PG2014-1), Welsh Assembly Government (grant SGR544:CADR), a donation from the Moondance Charitable Foundation, UK Dementia Platform (DPUK, reference MR/L023784/1), and the UK Dementia Research Institute at Cardiff. Cambridge University acknowledges support from the MRC. ARUK supported sample collections at the Kings College London, the South West Dementia Bank, Universities of Cambridge, Nottingham, Manchester and Belfast. King College London was supported by the NIHR Biomedical Research Centre for Mental Health and Biomedical Research Unit for Dementia at the South London and Maudsley NHS Foundation Trust and Kings College London and the MRC. Alzheimer Research UK (ARUK) and the Big Lottery Fund provided support to Nottingham University. Ulster Garden Villages, AS, ARUK, American Federation for Aging Research, NI R&D Office and the Royal College of Physicians/Dunhill Medical Trust provided support for Queen University, Belfast. The University of Southampton acknowledges support from the AS. The MRC and Mercer Institute for Research on Ageing supported the Trinity College group. DCR is a Wellcome Trust Principal Research fellow. The South West Dementia Brain Bank acknowledges support from Bristol Research into Alzheimer and Care of the Elderly. The Charles Wolfson Charitable Trust supported the OPTIMA group. Washington University was funded by NIH grants, Barnes Jewish Foundation and the Charles and Joanne Knight Alzheimer Research Initiative. Patient recruitment for the MRC Prion Unit/UCL Department of Neurodegenerative Disease collection was supported by the UCLH/UCL Biomedical Research Centre and their work was supported by the NIHR Queen Square Dementia BRU, the Alzheimer Research UK and the Alzheimer Society. LASER-AD was funded by Lundbeck SA. The AgeCoDe study group was supported by the German Federal Ministry for Education and Research grants 01 GI 0710, 01 GI 0712, 01 GI 0713, 01 GI 0714, 01 GI 0715, 01 GI 0716, 01 GI 0717. Genotyping of the Bonn case-control sample was funded by the German centre for Neurodegenerative Diseases (DZNE), Germany. The GERAD Consortium also used samples ascertained by the NIMH AD Genetics Initiative. HH was supported by a grant of the Katharina-Hardt-Foundation, Bad Homburg vor der H ohe, Germany. The KORA F4 studies were financed by Helmholtz Zentrum Munchen; German Research Center for Environmental Health; BMBF; German National Genome Research Network and the Munich Center of Health Sciences. The Heinz Nixdorf Recall cohort was funded by the Heinz Nixdorf Foundation and BMBF. We acknowledge use of genotype data from the 1958 Birth Cohort collection and National Blood Service, funded by the MRC and the Wellcome Trust which was genotyped by the Wellcome Trust Case Control Consortium and the Type-1 Diabetes Genetics Consortium, sponsored by the National Institute of Diabetes and Digestive and Kidney Diseases, National Institute of Allergy and Infectious Diseases, National Human Genome Research Institute, National Institute of Child Health and Human Development and Juvenile Diabetes Research Foundation International. The project is also supported through the following funding organisations under the aegis of JPND - www.jpnd.eu (United Kingdom, Medical Research Council (MR/L501529/1; MR/R024804/1) and Economic and Social Research Council (ES/L008238/1)) and through the Motor Neurone Disease Association. This study represents independent research part funded by the National Institute for Health Research (NIHR) Biomedical Research Centre at South London and Maudsley NHS Foundation Trust and King College London. Prof Jens Wiltfang is supported by an Ilidio Pinho professorship and iBiMED (UID/BIM/04501/2013), at the University of Aveiro, Portugal.
DemGene. The project has received funding from The Research Council of Norway (RCN) Grant Nos. 213837, 223273, 225989, 248778, and 251134 and EU JPND Program RCN Grant Nos. 237250, 311993, the South-East Norway Health Authority Grant No. 2013-123, the Norwegian Health Association, and KG Jebsen Foundation. The RCN FRIPRO Mobility grant scheme (FRICON) is co-funded by the European Union Seventh Framework Programme for research, technological development and demonstration under Marie Curie grant agreement No 608695. European Community grant PIAPP-GA-2011-286213 PsychDPC..
Bonn study. This group would like to thank Dr. Heike Koelsch for her scientific support. The Bonn group was funded by the German Federal Ministry of Education and Research (BMBF): Competence Network Dementia (CND) grant number 01GI0102, 01GI0711, 01GI042
China. This Chinese AD WGS cohort was supported in part by the National Key R&D Program of China (2021YFE0203000); the Areas of Excellence Scheme of the University Grants Committee (AoE/M-604/16); the Research Grants Council of Hong Kong (the Collaborative Research Fund [C6027-19GF] and the Theme-Based Research Scheme [T13-605/18W]); the Innovation and Technology Commission (InnoHK Funding Scheme; ITCPD/17-9, ITS/207/18FP, MRP/042/18X and MRP/097/20X); Chow Tai Fook Charity Foundation; the Guangdong Provincial Key S&T Program Grant (2018B030336001); the Guangdong Provincial Fund for Basic and Applied Basic Research (2019B1515130004); and the Fundamental Research Program of Shenzhen Virtual University Park (2021Szvup137).
Japan. This study is supported by AMED JP23dk0207060 and JP23wm0525019.
Korea. This research was supported by a National Research Foundation of Korea grant, funded by the Korean government (MSIT) (No. 2022R1A2C2009998); the ICT Creative Consilience program (No. IITP-2022-2020-0-01821) supervised by the Institute for Information & Communications Technology Planning & Evaluation (IITP); the Korea Health Technology R&D Project through the Korea Health Industry Development Institute, funded by the Ministry of Health and Welfare, Republic of Korea (Nos. HU22C0042, HU21C0111, and HI19C1132); the Ministry of Science and ICT, Republic of Korea (No. 2019RIA5A2026045); the National Institute of Health research project (No. 2021-ER1006-01); and Future Medicine 20*30 Project of the Samsung Medical Center (No. SMX1220021). This study was provided with biospecimens and data from the biobank of Chronic Cerebrovascular Disease consortium. The consortium was supported and funded by the Korea Centers for Disease Control and Prevention (No. 4845-303).
Sub Saharan Africa (EPIDEMCA). Sub Saharan Africa (EPIDEMCA). This study was supported by a grant from the French National Research Agency (ANR-09-MNPS-009-01),the AXA research Fund (2012 Project Public Health Institute (Inserm) PREUX Pierre-Marie) and the Limoges University Hospital through its APREL scheme. The sponsors of the study had no role in study design, data collection, data interpretation or writing of the report.
We also thank the staffs of the Universities of Bangui (Central African Republic) and Marien Ngouabi in Brazzaville (ROC); Pasteur Institute in Bangui and (Laboratoire National de Sante Publique) in Brazzaville; Health ministries of the Central African Republic and the Republic of Congo, for their moral support; University of Limoges, Doctoral School of Limoges University, Inserm; Limousin Regional Council. We are very grateful to all the participants to this survey, the investigators, and staffs of Bangui and Brazzaville hospitals for their assistance
Argentina. This study was supported by funding from Alexander von Humboldt Foundation and International Society for Neurochemistry (ISN) to M.C.D.; the Agencia Nacional de Promocion Cientifica y Tecnologica (PID-2011-0059, PIBT/09-2013, PICT-2016-4647 and PICT2019-0656 to L.M.) and from EU-LAC Health-Neurodegeneration JOINT CALL 2016 (EULACH16 to L.M.).
Chile. The Funding of the ALEXANDROS study was provided by the Chilean National Fund for Science and Technology (FONDECYT) grant 1130947. The genotyping for the Chilean and Tunisian series were funded by Genome Research @ Ace Alzheimer Center Barcelona project (GR@ACE), supported by Grifols SA, Fundacion bancaria (La Caixa), Ace Alzheimer Center Barcelona and CIBERNED.
Brazil. The study was supported by the Brazilian National Council for Scientific and Technological Development (CNPq): processes 315133/2021-0, 309953/2018-9, 436735/2018-0) FAPEMIG:processes:APQ-02662-14, APQ-02662-14) and the Coordination for the Improvement of Higher Education Personnel (CAPES): process 88887.569376/2020-00.
Colombia. This was supported by Ministerio de Ciencia, Tecnologia e Innovacion (MINCIENCIAS) and the Universidad Nacional de Colombia. We also thanks all the participants who kindly agreed to participate in the study.
ADSP. The ADGC cohorts include Adult Changes in Thought (ACT) (U01 AG006781, U19 AG066567),the Alzheimer Disease Research Centers (ADRC) (P30AG062429, P30 AG066468, P30AG062421, P30AG066509, P30AG066514, P30AG066530, P30AG066507, P30AG066444, P30AG066518, P30AG066512, P30AG066462, P30AG072979, P30AG072972, P30AG072976, P30AG072975, P30AG072978, P30AG072977, P30AG066519, P30AG062677, P30AG079280, P30AG062422, P30AG066511, P30AG072946, P30AG062715, P30AG072973, P30AG066506, P30AG066508, P30AG066515, P30AG072947, P30AG072931, P30AG066546, P20AG068024, P20AG068053, P20AG068077, P20AG068082, P30AG072958, P30AG072959), the Chicago Health and Aging Project (CHAP) (R01 AG11101, RC4 AG039085, K23 AG030944), Indiana Memory and Aging Study (IMAS) (R01 AG019771), Indianapolis Ibadan (R01 AG009956, P30 AG010133), the Memory and Aging Project (MAP) (R01 AG17917), Mayo Clinic (MAYO) (R01 AG032990, U01AG046139, R01 NS080820, RF1 AG051504, P50 AG016574), Mayo Parkinson Disease controls (NS039764, NS071674, 5RC2HG005605), University of Miami (R01 AG027944, R01 AG028786, R01 AG019085, IIRG09133827, A2011048), the Multi-Institutional Research in Alzheimer Genetic Epidemiology Study (MIRAGE) (R01 AG09029, R01 AG025259), the National Centralized Repository for Alzheimer Disease and Related Dementias (NCRAD) (U24 AG021886), the National Institute on Aging Late Onset Alzheimer Disease Family Study (NIA-LOAD) (U24AG056270), the Religious Orders Study (ROS) (P30 AG10161, R01 AG15819), the Texas Alzheimer Research and Care Consortium (TARCC) (funded by the Darrell K Royal Texas Alzheimer Initiative), Vanderbilt University/Case Western Reserve University (VAN/CWRU) (R01AG019757, R01 AG021547, R01 AG027944, R01 AG028786, P01 NS026630, and Alzheimer Association), the Washington Heights-Inwood Columbia Aging Project (WHICAP) (RF1AG054023), the University of Washington Families (VA Research Merit Grant, NIA: P50AG005136, R01AG041797, NINDS: R01NS069719), the Columbia University Hispanic Estudio Familiar de Influencia Genetica de Alzheimer (EFIGA) (RF1 AG015473), the University of Toronto (UT)(funded by Wellcome Trust, Medical Research Council, Canadian Institutes of Health Research), and Genetic Differences (GD) (R01 AG007584). The CHARGE cohorts are supported in part by National Heart, Lung, and Blood Institute (NHLBI) infrastructure grant HL105756 (Psaty), RC2HL102419 (Boerwinkle) and the neurology working group is supported by the National Institute on Aging (NIA) R01 grant AG033193. The CHARGE cohorts The CHARGE cohorts participating in the ADSP include the following: Austrian Stroke Prevention Study (ASPS), ASPS-Family study, and the Prospective Dementia Registry-Austria (ASPS/PRODEM-Aus), the Atherosclerosis Risk in Communities (ARIC) Study, the Cardiovascular Health Study (CHS), the Erasmus Rucphen Family Study (ERF), the Framingham Heart Study (FHS), and the Rotterdam Study (RS). ASPS is funded by the Austrian Science Fond (FWF) grant number P20545-P05 and P13180 and the Medical University of Graz. The ASPS-Fam is funded by the Austrian Science Fund (FWF) project I904), the EU Joint Programme Neurodegenerative Disease Research (JPND) in frame of the BRIDGET project (Austria, Ministry of Science) and theMedical University of Graz and the Steiermarkische Krankenanstalten Gesellschaft. PRODEM-Austria is supported by the Austrian Research Promotion agency (FFG) (Project No.827462) and by the Austrian National Bank (Anniversary Fund, project 15435. ARIC research is .CC-BY-NC-ND 4.0 International licenseIt is made available under a is the author/funder, who has granted medRxiv a license to display the preprint in perpetuity. The collaborative study is also supported by NHLBI contracts (HHSN268201100005C,HHSN268201100006C, HHSN268201100007C, HHSN268201100008C, HHSN268201100009C,Neurocognitive data in ARIC is collected by U01 2U01HL096812, 2U01HL096814, 2U01HL096899, 2U01HL096902,2U01HL096917 from the NIH (NHLBI, NINDS, NIA and NIDCD), and with previous brain MRI examinations funded by R01-HL70825 from the NHLBI. CHS research was supported by contracts HHSN268201200036C, HHSN268200800007C, N01HC55222, N01HC85079, N01HC85080, N01HC85081, N01HC85082, N01HC85083, N01HC85086, and grants U01HL080295 and U01HL130114 from the NHLBI with additional contribution from the National Institute of Neurological Disorders and Stroke (NINDS). Additional support was provided by R01AG023629, R01AG15928, and R01AG20098 from the NIA. FHS research is supported by NHLBI contracts N01-HC-25195 and HHSN268201500001I. This study was also supported by additional grants from the NIA (R01s AG054076, AG049607 and AG033040 and NINDS (R01 NS017950). The ERF study as a part of EUROSPAN (European Special Populations Research Network) was supported by European Commission FP6 STRP grant number 018947 (LSHG-CT-2006-01947) and also received funding from the European Community Seventh Framework Programme (FP7/2007-2013)/grant agreement HEALTH-F4-2007-201413 by the European Commission under the programme (Quality of Life and Management of the Living Resources) of 5th Framework Programme (no.QLG2-CT-2002-01254). High-throughput analysis of the ERF data was supported by a joint grant from the Netherlands Organization for Scientific Research and the Russian Foundation for Basic Research (NWO-RFBR 047.017.043). The Rotterdam Study is funded by Erasmus Medical Center. Genetic data sets are also supported by the Netherlands Organization of Scientific Research NWO Investments (175.010.2005.011,584 911-03-012), the Genetic Laboratory of the Department of Internal Medicine, Erasmus MC, the Research Institute for Diseases in the Elderly (014-93-015; RIDE2), and the Netherlands Genomics Initiative (NGI)/Netherlands Organization for Scientific Research (NWO) Netherlands Consortium for Healthy Aging (NCHA), project 050-060-810. All studies are grateful to their participants, faculty and staff. The content of these manuscripts is solely the responsibility of the authors and does not necessarily represent the official views of the National Institutes of Health or the U.S. Department of Health and Human Services. In addition, funding to support and share the ADSP data are included here: NIA U54-AG052427 and NIA U24-AG041689.
The Sacramento Area Latino Study on Aging (SALSA). This study was funded by the National Institutes of Health, including the National Institute on Aging under Award Numbers K01AG056602 and R01AG12975 and the National Institute for Diabetes and Digestive and Kidney Diseases under Award Numbers R01 DK60753 and R01DK087864.
MVP. This research is based on data from the Million Veteran Program, Office of Research and Development, Veterans Health Administration, and was supported by VA BLR&D grants 1 I01 BX004192 (MVP015) and I01BX005749 (MVP040). The views expressed in this article are those of the authors and do not necessarily reflect the position or policy of the Department of Veterans Affairs or the US government.

### Author Declarations

Written informed consent was obtained from study participants or, for those with substantial cognitive impairment, from a caregiver, legal guardian or other proxy. Study protocols for all cohorts were reviewed and approved by the appropriate institutional review boards. All necessary patient/participant consent has been obtained and the appropriate institutional forms have been archived.

### Summary of Updates

- All the analyses were re-ran based on the increase in the AD risk associated with an increment of one standard deviation in the GRS rather than as the increase in the risk of AD associated with an increment of one allele of average risk in the GRS. - Nearly 7,000 samples with measurements of tau, p-tau and Abeta42 in cerebrospinal fluid were added - Where possible, analyses with or without calibration were systematically performed to correct for any shift in the GRS/PRS distributions in different populations. - New analyses were added such as sex stratification or a GRS (named GRS+) that included additional SNPs at the European GWAS-defined loci, potentially associated with risk of developing AD in non-European multi-ancestry populations. - PRS-CSx, a Bayesian polygenic modelling method was implemented to construct a cross-ancestry polygenic risk score.

## Reference

1. Osterman, M. D., Kinzy, T. G. & Bailey, J. N. C. Polygenic Risk Scores. Curr Protoc 1, e126 (2021).

2. Kachuri, L. et al. Principles and methods for transferring polygenic risk scores across global populations. Nat Rev Genet 25, 8–25 (2024).

3. Gatz, M. et al. Role of genes and environments for explaining Alzheimer disease. Arch Gen Psychiatry 63, 168–74 (2006).

4. Baker, E. & Escott-Price, V. Polygenic Risk Scores in Alzheimer’s Disease: Current Applications and Future Directions. Front Digit Health 2, 14 (2020).

5. Desikan, R. S. et al. Genetic assessment of age-associated Alzheimer disease risk: Development and validation of a polygenic hazard score. PLoS Med 14, e1002258 (2017).

6. Sabuncu, M. R. et al. The association between a polygenic Alzheimer score and cortical thickness in clinically normal subjects. Cereb Cortex 22, 2653–61 (2012).

7. Mormino, E. C. et al. Polygenic risk of Alzheimer disease is associated with early- and late-life processes. Neurology 87, 481–8 (2016).

8. Xicota, L. et al. Association of APOE-Independent Alzheimer Disease Polygenic Risk Score With Brain Amyloid Deposition in Asymptomatic Older Adults. Neurology 99, e462–e475 (2022).

9. Sleegers, K. et al. A 22-single nucleotide polymorphism Alzheimer’s disease risk score correlates with family history, onset age, and cerebrospinal fluid Aβ42. Alzheimers Dement 11, 1452–1460 (2015).

10. Hong, S. et al. Genome-wide association study of Alzheimer’s disease CSF biomarkers in the EMIF-AD Multimodal Biomarker Discovery dataset. Transl Psychiatry 10, 403 (2020).

11. Clark, K., Leung, Y. Y., Lee, W.-P., Voight, B. & Wang, L.-S. Polygenic Risk Scores in Alzheimer’s Disease Genetics: Methodology, Applications, Inclusion, and Diversity. J Alzheimers Dis 89, 1–12 (2022).

12. Sariya, S. et al. Polygenic Risk Score for Alzheimer’s Disease in Caribbean Hispanics. Ann Neurol 90, 366–376 (2021).

13. Jung, S.-H. et al. Transferability of Alzheimer Disease Polygenic Risk Score Across Populations and Its Association With Alzheimer Disease-Related Phenotypes. JAMA Netw Open 5, e2247162 (2022).

14. Dalmasso, M. C. et al. The first genome-wide association study in the Argentinian and Chilean populations identifies shared genetics with Europeans in Alzheimer’s disease. Alzheimers Dement 20, 1298–1308 (2024).

15. Kikuchi, M. et al. Polygenic effects on the risk of Alzheimer’s disease in the Japanese population. Alzheimers Res Ther 16, 45 (2024).

16. Bellenguez, C. et al. New insights into the genetic etiology of Alzheimer’s disease and related dementias. Nat Genet 54, 412–436 (2022).

17. Koyama, S. et al. Population-specific and trans-ancestry genome-wide analyses identify distinct and shared genetic risk loci for coronary artery disease. Nat Genet 52, 1169–1177 (2020).

18. Lennon, N. J. et al. Selection, optimization and validation of ten chronic disease polygenic risk scores for clinical implementation in diverse US populations. Nat Med 30, 480–487 (2024).

19. Keaton, J. M. et al. Genome-wide analysis in over 1 million individuals of European ancestry yields improved polygenic risk scores for blood pressure traits. Nat Genet 56, 778–791 (2024).

20. Ge, T. et al. Development and validation of a trans-ancestry polygenic risk score for type 2 diabetes in diverse populations. Genome Med 14, 70 (2022).

21. Kachuri, L. et al. Principles and methods for transferring polygenic risk scores across global populations. Nat Rev Genet 25, 8–25 (2024).

22. Xiang, R. et al. Recent advances in polygenic scores: translation, equitability, methods and FAIR tools. Genome Med 16, 33 (2024).

23. Jansen, I. E. et al. Genome-wide meta-analysis for Alzheimer’s disease cerebrospinal fluid biomarkers. Acta Neuropathol 144, 821–842 (2022).

24. Logue, M. W., Dasgupta, S. & Farrer, L. A. Genetics of Alzheimer’s Disease in the African American Population. J Clin Med 12, (2023).

25. Miyashita, A., Kikuchi, M., Hara, N. & Ikeuchi, T. Genetics of Alzheimer’s disease: an East Asian perspective. J Hum Genet 68, 115–124 (2023).

26. Kunkle, B. W. et al. Novel Alzheimer Disease Risk Loci and Pathways in African American Individuals Using the African Genome Resources Panel: A Meta-analysis. JAMA Neurol 78, 102–113 (2021).

27. Lake, J. et al. Multi-ancestry meta-analysis and fine-mapping in Alzheimer’s disease. Mol Psychiatry 28, 3121–3132 (2023).

28. Shigemizu, D. et al. Ethnic and trans-ethnic genome-wide association studies identify new loci influencing Japanese Alzheimer’s disease risk. Transl Psychiatry 11, 151 (2021).

29. Frisoni, G. B. et al. The probabilistic model of Alzheimer disease: the amyloid hypothesis revised. Nat Rev Neurosci 23, 53–66 (2022).

30. Wang, Y. et al. Polygenic prediction across populations is influenced by ancestry, genetic architecture, and methodology. Cell genomics 3, 100408 (2023).

31. Smith, J. L. et al. Multi-Ancestry Polygenic Risk Score for Coronary Heart Disease Based on an Ancestrally Diverse Genome-Wide Association Study and Population-Specific Optimization. Circ Genom Precis Med 17, e004272 (2024).

32. Khan, A. et al. Genome-wide polygenic score to predict chronic kidney disease across ancestries. Nat Med 28, 1412–1420 (2022).

33. Naslavsky, M. S. et al. Global and local ancestry modulate APOE association with Alzheimer’s neuropathology and cognitive outcomes in an admixed sample. Mol Psychiatry 27, 4800–4808 (2022).

34. Rajabli, F. et al. Ancestral origin of ApoE ε4 Alzheimer disease risk in Puerto Rican and African American populations. PLoS Genet 14, e1007791 (2018).

35. Bussies, P. L. et al. Use of local genetic ancestry to assess TOMM40-523’ and risk for Alzheimer disease. Neurol Genet 6, e404 (2020).

36. Rajabli, F. et al. A locus at 19q13.31 significantly reduces the ApoE ε4 risk for Alzheimer’s Disease in African Ancestry. PLoS Genet 18, e1009977 (2022).

37. Nagao, Y. Contribution of rare variants to heritability of a disease is much greater than conventionally estimated: modification of allele distribution model. J Hum Genet (2024) doi:10.1038/s10038-024-01281-2.

## Reference

38. Das, S. et al. Next-generation genotype imputation service and methods. Nat Genet 48, 1284–1287 (2016).

39. Taliun, D. et al. Sequencing of 53,831 diverse genomes from the NHLBI TOPMed Program. Nature 590, 290–299 (2021).

40. Chen, C.-Y. et al. Improved ancestry inference using weights from external reference panels. Bioinformatics 29, 1399–406 (2013).

41. Le Guen, Y. et al. Multiancestry analysis of the HLA locus in Alzheimer’s and Parkinson’s diseases uncovers a shared adaptive immune response mediated by HLA-DRB1*04 subtypes. Proc Natl Acad Sci U S A 120, e2302720120 (2023).

42. Fang, H. et al. Harmonizing Genetic Ancestry and Self-identified Race/Ethnicity in Genome-wide Association Studies. Am J Hum Genet 105, 763–772 (2019).

43. Abraham, G., Qiu, Y. & Inouye, M. FlashPCA2: principal component analysis of Biobank-scale genotype datasets. Bioinformatics 33, 2776–2778 (2017).

44. Ruan, Y. et al. Improving polygenic prediction in ancestrally diverse populations. Nat Genet 54, 573–580 (2022).

45. Ma, Y. & Zhou, X. Genetic prediction of complex traits with polygenic scores: a statistical review. Trends Genet 37, 995–1011 (2021).

46. Kurniansyah, N. et al. Evaluating the use of blood pressure polygenic risk scores across race/ethnic background groups. Nat Commun 14, 3202 (2023).

47. Chang, C. C. et al. Second-generation PLINK: rising to the challenge of larger and richer datasets. Gigascience 4, 7 (2015).

48. Hao, L. et al. Development of a clinical polygenic risk score assay and reporting workflow. Nat Med 28, 1006–1013 (2022).

49. Lee, S. H., Goddard, M. E., Wray, N. R. & Visscher, P. M. A better coefficient of determination for genetic profile analysis. Genet Epidemiol 36, 214–24 (2012).

50. Choi, S. W., Mak, T. S.-H. & O’Reilly, P. F. Tutorial: a guide to performing polygenic risk score analyses. Nat Protoc 15, 2759–2772 (2020).

51. Lee, S. H. & Wray, N. R. Novel genetic analysis for case-control genome-wide association studies: quantification of power and genomic prediction accuracy. PLoS One 8, e71494 (2013).

52. Baker, E. et al. What does heritability of Alzheimer’s disease represent? PLoS One 18, e0281440 (2023).

53. Willer, C. J., Li, Y. & Abecasis, G. R. METAL: fast and efficient meta-analysis of genomewide association scans. Bioinformatics 26, 2190–1 (2010).

