## Supplementary material for "Transferability of European-derived Alzheimer’s Disease Polygenic Risk Scores across Multi-Ancestry Populations": Supplemnetary Materials

Aude Nicolas et al.

TABLE OF CONTENTS


1. **Sample Description** **2**
2. **Supplementary figures 1-13** **15**
3. **Supplementary References** **28**
4. **Acknowledgements** **31**
5. **Supplementary list of authors** **40**

### 1. SAMPLE DESCRIPTION

Demographics of the different case-control studies are fully described in the supplementary Table 1

***<u>The European Alzheimer & Dementia Biobank dataset (EADB)</u>***

This consortium groups together 20,464 Alzheimer's disease (AD) cases and 22,244 controls after quality controls from 15 European countries (Belgium, Bulgaria, Czech Republic, Denmark, Finland, France, Germany, Greece, Italy, Portugal, Spain, Sweden, Switzerland, The Netherlands and the UK). These samples were genotyped using the ILLUMINA GSA array in three independent centers (France, Germany and the Netherlands) leading to define three nodes: EADB-France, EADB-Germany and EADB-Netherlands.

**EADB-France**
In the France node, samples were collected from nine countries (39 centers/studies), and after quality controls (QCs), we obtained 13,867 AD cases and 15,310 controls. All these samples were genotyped at the Centre National de Recherche en Génomique Humaine (CNRGH, Evry, France).

<u>Belgium:</u> The participants were part of a large prospective cohort[1] of Belgian AD patients and healthy elderly control individuals. The patients were ascertained at the memory clinic of Middelheim and Hoge Beuken (Hospital Network Antwerp, Belgium) and at the memory clinic of the University Hospitals of Leuven, Belgium. The control individuals were the partners of the patients or volunteers from the Belgian community. The study protocols were approved by the ethics committees of the Antwerp University Hospital and the participating neurological centers at the different hospitals of the BELNEU consortium and by the University of Antwerp.

<u>Czech Republic:</u> The Czech Brain Aging Study (CBAS)[2] is a longitudinal memory-clinic–based study recruiting subjects at risk of dementia (subjects referred for cognitive complaints-SCD, MCI). The CBAS+ study is a cross-sectional study of patients in the early stages of dementia. All subjects signed informed consent and both studies were approved by the local ethics committee.

<u>Denmark:</u> The Copenhagen General Population Study (CGPS) is a prospective study of the Danish general population initiated in 2003 and still recruiting. Individuals were selected randomly based on the national Danish Civil Registration System to reflect the adult Danish population aged 20-100. Data were obtained from a self-administered questionnaire reviewed together with an investigator at the day of attendance, a physical examination, and from blood samples including DNA extraction.

<u>Finland:</u> *The ADGEN cohort*[3]: a clinic-based collection of AD patients from Eastern and Northern Finland examined in the Department of Neurology in Kuopio University Hospital and the Department of Neurology in Oulu University Hospital. All the patients were diagnosed with probable AD according to the criteria of the National Institute of Neurological and Communicative Disorders and Stroke and the Alzheimer's disease and Related Disorders Association (NINCDS–ADRDA). The study was approved by the ethics committee of Kuopio University Hospital, Finland (420/2016). *The FINGER study*[4]: a Finnish multi-domain lifestyle RCT enrolling 1,260 older adults with an increased risk of dementia from the general population. The intensive lifestyle intervention lasted for two years, and follow-up extends currently up to seven years. The FINGER study was approved by the coordinating ethics committee of the Hospital District of Helsinki and Uusimaa (94/13/03/00/2009 and HUS/1204/2017), and all the participants gave written informed consent.

<u>France:</u> *The BALTAZAR multicenter (23 memory centers) prospective study*[5]: 1,040 participants from September 2010 to April 2015. They were classified as AD cases (n = 501) according to DSM IV-TR and NINCDS–ADRDA criteria as well as amnestic mild cognitive impairment (MCI) cases (a MCI, n = 417) and non-amnestic MCI cases (na MCI, n = 122)

according to Petersen's criteria. A comprehensive battery of cognitive tests was performed, including MMSE, verbal fluency, and FCSRT. All the participants or their legal guardians gave written informed consent. The study was approved by the Paris ethics committee (CPP Ile de France IV Saint Louis Hospital). *MEMENTO*: a clinic-based study[6] aimed at better understanding the natural history of AD, dementia, and related diseases. Between 2011 and 2014, 2,323 individuals presenting either recently diagnosed MCI or isolated cognitive complaints were enrolled in 26 memory centers in France. This study was performed in accordance with the guidelines of the Declaration of Helsinki. The MEMENTO study protocol has been approved by the local ethics committee (Comité de Protection des Personnes Sud-Ouest et Outre Mer III; approval number 2010-A01394-35). All the participants provided written informed consent. *The CNRMAJ-Rouen study*[7]: early onset AD patients (n = 870). The patients or their legal guardians provided written informed consent. This study was approved by the ethics committee of CPP Ile de France II.

Italy: The AD cases and controls were collated through Italy in different centers: Brescia, Cagliari, Florence, Milan, Rome, Pertugia, San Giovani Rotondo and Torino. AD cases were diagnosed according to DSM III-R,IV and NINCDS–ADRDA criteria. Controls were defined a minima as subjects without DMS-III-R dementia criteria and with integrity of their cognitive functions (MMS>25).

Spain: The Dementia Genetic Spanish Consortium (DEGESCO) is a national consortium comprising 23 research centers and hospitals across the country, that holds the institutional coverage of The Network Center for Biomedical Research in Neurodegenerative Diseases (CIBERNED). Created in 2013, DEGESCO's objective is the promotion and conduction of genetic studies aimed at understanding the genetic architecture of neurodegenerative dementias in the Spanish population and participates in coordinated actions in national and international frameworks. All DNA samples are in compliance with the Law of Biomedical Research (Law 14/2007) and the Royal Decree on Biobanks (RD 1716/2011). Patients included in the present study met clinical criteria for probable or possible disease established by the National Institute of Neurological and Communication Disorders and Stroke and the Alzheimer Disease and Related Disorders Association (NINCDS-ADRDA). Cognitively healthy controls were unrelated individuals who had a documented MMSE in the normal range. Contributing centers in the France node genotyping were Centro de Biología Molecular Severo Ochoa (CSIC-UAM (Madrid), the Institute Biodonostia, University of Basque Contry (EHU-UPV, San Sebastián), Institut de Biomedicina de Valencia CSIC (València), and Sant Pau Biomedical Research Institute (Barcelona).

Sweden: *Upsala.* The Swedish AD patients were ascertained at the Memory Disorder Unit at Uppsala University Hospital. For all patients, the diagnosis was established according to the National Institute on Neurological Disorders and Stroke, and the Alzheimer's Disease and Related Disorders Association (NINDS-ADRDA) guidelines[8]. Healthy control subjects were recruited from the same geographic region following advertisements in local newspapers and displayed no signs of dementia upon Mini Mental State Examination (MMSE). *Swedish National Study on Aging and Care in Kungsholmen (SNAC-K)* data was collected. The original SNAC-K population consisted of 4590 living and eligible persons who lived on the island of Kungsholmen in Central Stockholm, belonged to pre-specified age strata, and were randomly selected to take part in the study. Between 2001 and 2004, 3363 persons participated in the baseline assessment. They belonged to the age cohorts 60, 66, 72, 78, 81, 84, 87, 90, 93, and 96 years and 99 years and older. The examination consists of three parts: a nurse interview, a medical examination, and a neuropsychological testing session. Altogether, the examination takes about six hours. The participants are reexamined each time they reach the next age cohort. All parts of the SNAC-K project have been approved by the ethical committee at Karolinska Institutet or the regional ethical review board. Informed consent was collected from all the participants or, if the person was severely cognitively impaired, from their next of kin.

The UK: *MRC*. The sample set comprises individuals with AD and healthy controls recruited across the MRC Centre for Neuropsychiatric Genetics and Genomics, Cardiff University, Cardiff, UK; Institute of Psychiatry, London, UK; University of Cambridge, Cambridge, UK. The collection of the samples was through multiple channels, including specialist NHS services and clinics, research registers and Join Dementia Research (JDR) platform. The participants were assessed at home or in research clinics along with an informant, usually a spouse, family member or close friend, who provided information about and on behalf of the individual with dementia. Established measures were used to ascertain the disease severity: Bristol activities of daily living (BADL), Clinical Dementia Rating scale (CDR), Neuropsychiatric Inventory (NPI) and Global Deterioration Scale (GlDS). Individuals with dementia completed the Addenbrooke's Cognitive Examination (ACE-r), Geriatric Depression Scale (GeDS) and National Adult Reading Test (NART) too. Control participants were recruited from GP surgeries and by means of self-referral (including existing studies and Joint Dementia Research platform). For all other recruitment, all AD cases met criteria for either probable (NINCDS-ADRDA, DSM-IV) or definite (CERAD) AD. All elderly controls were screened for dementia using the Mini Mental State Examination (MMSE) or ADAS-cog, were determined to be free from dementia at neuropathological examination or had a Braak score of 2.5 or lower. Control samples were chosen to match case samples for age, gender, ethnicity and country of origin. Informed consent was obtained for all study participants, and the relevant independent ethical committees approved study protocols. *SOTON, University of Southampton, Southampton, UK*. All AD cases met criteria for either probable (NINCDS-ADRDA, DSM-IV) or definite (CERAD) AD. All elderly controls were screened for dementia using the MMSE or ADAS-cog, were determined to be free from dementia at neuropathological examination or had a Braak score of 2.5 or lower. *Nottingham and Manchester*, *University of Nottingham, Nottingham, UK and Manchester Brain Bank*. All AD cases met criteria for either probable (NINCDS-ADRDA, DSM-IV) or definite (CERAD) AD. All elderly controls were screened for dementia using the MMSE or ADAS-cog, were determined to be free from dementia at neuropathological examination or had a Braak score of 2.5 or lower. *KCL, London Neurodegenerative Diseases Brain Bank*. All AD cases met criteria for either probable (NINCDS-ADRDA, DSM-IV) or definite (CERAD) AD. All elderly controls were screened for dementia using the MMSE or ADAS-cog, were determined to be free from dementia at neuropathological examination or had a Braak score of 2.5 or lower. *PRION,* All AD cases met criteria for either probable (NINCDS-ADRDA, DSM-IV) or definite (CERAD) AD. All elderly controls were screened for dementia using the MMSE or ADAS-cog, were determined to be free from dementia at neuropathological examination or had a Braak score of 2.5 or lower. *CFAS Wales*, The Cognitive Function and Ageing Study Wales (CFAS-Wales) is a longitudinal population-based study of people aged 65 years and over in rural and urban areas of Wales that aims to investigate physical and cognitive health in older age and examine the interactions between health, social networks, activity, and participation. Individuals aged 65 years and over were randomly sampled from general medical practice lists between 2011 and 2013, stratified by age to ensure equal numbers in two age groups, 65-74 years and 75 and over. The baseline sample included 3593 older people and included those living in care homes as well as those living at home. Those who provided written consent to join the study were interviewed in their own homes by trained interviewers and could choose to have the interview conducted through the medium of either English or Welsh. Participants were followed up 2 years later. All AD cases met criteria for either probable (NINCDS-ADRDA, DSM-IV) or definite (CERAD) AD. All elderly controls were screened for dementia using the MMSE or CAMCOG, and were determined to be free from dementia. *UCL-DRC*. the UCL Alzheimer's disease cohort of the Dementia Research Centre (UCL - EOAD DRC) included patients seen at the Cognitive Disorders Clinics at The National Hospital for Neurology and Neurosurgery (Queen Square), or affiliated hospitals. Individuals were assessed clinically and diagnosed as having probable Alzheimer's disease based on contemporary clinical criteria in use at the time, including imaging and neuropsychological testing where appropriate.

**EADB-Germany**

In the German node, samples were collected from seven countries (11 centers/studies) and after QCs, we obtained 4,159 AD cases and 4,545 controls. All these samples were genotyped at Life&brain (Bonn, Germany).

Germany: DELCODE (the multicenter DZNE-Longitudinal Cognitive Impairment and Dementia Study). This is an observational longitudinal memory clinic-based multicenter study in Germany comprising 400 subjects with Subjective cognitive decline (SCD), 200 mild cognitive impairment (MCI) patients, 100 AD dementia patients, 200 control subjects without subjective or objective cognitive decline, and 100 first-degree relatives of patients with a documented diagnosis of AD dementia. All patient groups (SCD, MCI, AD) are referrals, including self-referrals, to the participating memory centers. The control group and the relatives of AD dementia patients are recruited by standardized public advertisement. Ten university-based memory centers are participating, all being collaborators of local DZNE sites. All patient groups (SCD, MCI, AD) were assessed clinically at the respective memory centers before entering DELCODE. The assessments include medical history, psychiatric and neurological examination, neuropsychological testing, blood laboratory work-up, cerebrospinal fluid (CSF) biomarkers, and routine MRI, all according to the local standards. The Consortium to Establish a Registry for Alzheimer's Disease (CERAD) neuropsychological test battery was applied at all memory centers to measure cognitive function. German age, sex, and education-adjusted norms of the CERAD neuropsychological battery are available online (www.memoryclinic.ch). Detail description is of recruitment protocol is reported elsewhere. *The VOGEL study*: The VOGEL study is a prospective, observational, long-term follow-up study with three time points of investigation within 6–8 years. This cohort includes dementia and healthy subjects. Residents of the city of Würzburg born between 1936 and 1941 were recruited. Every participant underwent physical, psychiatric, and laboratory examinations and performed intense neuropsychological testing as well as VSEP and NIRS according to the published procedures. A total of 604 subjects were included. *The Heidelberg/Mannheim memory clinic sample:* This cohort includes 61 subjects from whom 40 MCI patients were recruited and assessed between 2012 and 2016. Some of those patients converted to dementia by AD or other dementias. *The PAGES study:* This study includes 301 subjects. AD patients were recruited at the memory clinic of the Department of Psychiatry, University of Munich, Germany. Participants in whom dementia associated with AD was diagnosed fulfilled the criteria for probable AD according to the NINCDS–ADRDA. The control group included participants who were randomly selected from the general population of Munich. Controls who had central nervous system diseases or psychotic disorders or who had first-degree relatives with psychotic disorders were excluded. *The Technische Universität München study:* This cohort includes 359 healthy, AD, and other dementias patients recruited from the Centre for Cognitive Disorders. All the participants provided written informed consent. A biobank was submitted to the ethics committee of the Technical University of Munich, School of Medicine (Munich, Germany), which raised no objections and approved the biobank (reference number 347-14). *The Göttingen Universität study:* This study includes 111 in- and outpatients with a healthy or AD dementia status from the Department of Psychiatry of the University of Göttingen. The study's ethical statement was provided locally at the Göttingen University Medical Centre. *The German Dementia Competence Network (DCN) cohort:* Individuals from the DCN cohort were recruited from 14 university hospital memory clinics across Germany between 2003 and 2005[9]. The study was approved by the respective ethics committees, and written informed consent was obtained from all the participants prior to inclusion. *The German Study on Aging, Cognition, and Dementia (AgeCoDe):* The AgeCoDe study is a general practice (GP) registry-based longitudinal study in elderly individuals that recruited patients aged 75 years and above in six German cities from 2003 to 2004[10]. The study was approved by the respective ethics committees, and written informed consent was obtained from all the participants prior to inclusion.

Greece: *the HELIAD study*, comprising 49 AD cases and 1,150 controls. HELIAD is a population-based, multidisciplinary, collaborative study designed to estimate, in the Greek population over the age of 64 years, the prevalence and incidence of MCI, AD, other forms of dementia, and other neuropsychiatric conditions of aging and to investigate associations between nutrition and cognitive dysfunction or age-related neuropsychiatric diseases. The participants were selected through random sampling from the records of two Greek municipalities, Larissa and Marousi. All the participants signed informed consent in Greek.

Portugal: *the Lisbon study* from Portugal, totaling 78 AD cases and 74 controls. This cohort was recruited in 2008–2009 to investigate the connections between oxidative stress and lipid dyshomeostasis in AD. The project includes 190 subjects and was approved by the local ethics committee, and all the participants provided written informed consent. This study includes healthy and dementia-by-AD subjects.

Spain: Those samples are part of DEGESCO. DEGESCO Centers from whom DNA samples were genotyped in the German node (1,778 cases and 470 controls) were the Alzheimer Research Center and Memory Clinic, Fundació ACE, Institut Català de Neurociències Aplicades (Barcelona), the Neurology Service at University Hospital Marqués de Valdecilla (Santander), the Alzheimer's disease and other cognitive disorders, Neurology Department, at Hospital Clínic, IDIBAPS (Barcelona), the Molecular Genetics Laboratory, at the Hospital Universitario Central de Asturias (Oviedo), and Fundació Docència i Recerca Mútua de Terrassa and Movement Disorders Unit, Department of Neurology, University Hospital Mútua de Terrassa (Barcelona).

Switzerland: Two datasets from Switzerland and Austria were combined, totaling 182 AD cases and 388 controls. *The Lausanne study*: This study includes 137 community-dwelling participants aged 55+ years with cognitive impairment (memory clinic patients with MCI, dementia) or normal cognition (recruited by advertisement, word of mouth). The study's ethical statement was provided locally at the Department of Psychiatry, Geneva University Centre, Switzerland. *The VITA study:* This is a longitudinal study of 606 individuals (Vienna, Austria) who were 75 years old in 2000, followed up every 30–90 months. This cohort includes dementia and healthy subjects. All the participants gave written informed consent. The study conformed to the latest version of the Declaration of Helsinki and was approved by the ethics committee of the City of Vienna, Austria

**EADB-Netherlands**

In the Dutch node, samples were collected from six organizations in the Netherlands and after QCs, we obtained 2,438 AD cases and 2,389 controls. All these samples were genotyped at the Erasmus Medical University (Rotterdam, The Netherlands). The Medical Ethics Committee (METC) of the local institutes approved the studies. All the participants and/or their legal guardians gave written informed consent for participation in the clinical and genetic studies. Samples from the following institutes were included. 1) *Erasmus Medical Center:* most individuals were selected from population studies from the epidemiology department and accounted for most of the controls, while a smaller subset of samples originated from the neurology department, where AD was diagnosed according to the National Institute of Neurological and Communicative Disorders and Stroke-Alzheimer's Disease and Related Disorders Association (NINCDS-ADRDA) criteria for AD[11]. 2) *The Amsterdam Dementia Cohort (ADC)*[12]: This cohort comprises patients who visit the memory clinic of the VU University Medical Centre, the Netherlands. The diagnosis of probable AD is based on the clinical criteria formulated by the NINCDS–ADRDA and based on the NIA–AA. Diagnosis of MCI was made according to Petersen and NIA-AA. Controls presented with subjective cognitive decline at the memory clinic, but performed within normal limits on all clinical investigations. 3) *The 100-Plus study:* This study includes Dutch-speaking individuals who (i) can provide official evidence for being aged 100 years or older, (ii) self-report to be cognitively healthy, which is confirmed by a proxy, (iii) consent to the donation of a blood sample, (iv) consent to (at least) two home visits from a researcher, and (v) consent to undergo an interview and neuropsychological test battery[13]. 4) *Parelsnoer Institute:* a collaboration between 8 Dutch University Medical Centers in which clinical data and biomaterials from patients suffering from chronic diseases (so called "Pearls") are collected according to harmonized protocols. The Pearl Neurodegenerative Diseases[14] includes individuals diagnosed with dementia, mild cognitive impairment, and controls with subjective memory complaints. 5) *The Netherlands Brain Bank:* a non-profit organization that collects human brain tissue of donors with a variety of neurological and psychiatric disorders, but also of non-diseased donors. A clinical diagnosis of AD is based on the clinical criteria of probable AD[8,15]. The selected AD patients for this study all received a definitive diagnosis which was based on autopsy. 6) *Maastricht University Medical Center*: a subset of individuals that were referred to the memory clinic for cognitive complaints were included if they participated in the BioBank-Alzheimer Centrum Limburg (BB-ACL)[16]. Diagnosis of MCI was made according to the criteria of Petersen, and diagnosis of AD-type dementia was made according to the criteria of the DSM-4[17], and the NINCDS-ADRDA[8]. The Alzheimer Center Amsterdam is supported by Stichting Alzheimer Nederland and Stichting VUmc fonds. The clinical database structure was developed with funding from Stichting Dioraphte. Genotyping of the Dutch case-control samples was performed in the context of EADB (European Alzheimer DNA biobank) funded by the JPco-fuND FP-829-029 (ZonMW projectnumber 733051061).

***GR@ACE***

The GR@ACE study[19] recruited Alzheimer's disease (AD) patients from Ace Alzheimer Center Barcelona (Barcelona, Spain), and control individuals from three centers: Ace Alzheimer Center Barcelona (Barcelona, Spain), Valme University Hospital (Seville, Spain), and the Spanish National DNA Bank–Carlos III (University of Salamanca, Spain) (http://www.bancoadn.org). Additional cases and controls were obtained from dementia cohorts included in the Dementia Genetics Spanish Consortium (DEGESCO)[20]. At all sites, AD diagnosis was established by a multidisciplinary working group—including neurologists, neuropsychologists, and social workers—according to the DSM-IV criteria for dementia and the National Institute on Aging and Alzheimer's Association's (NIA–AA) 2011 guidelines for diagnosing AD. In our study, we considered as AD cases any individuals with dementia diagnosed with probable or possible AD at any point in their clinical course.
Genotyping was conducted using the Axiom 815K Spanish biobank array (Thermo Fisher) at the Spanish National Centre for Genotyping (CeGEN, Santiago de Compostela, Spain). The genotyping array not only is an adaptation of the Axiom biobank genotyping array but also contains rare population-specific variations observed in the Spanish population.

**The Rotterdam Study**

The Rotterdam Study is a prospective population-based middle-aged and elderly cohort that started in 1990 in the district of Ommoord, in Rotterdam, The Netherlands. The study includes 14,926 participants and has three subcohorts[21]. At start of the study, all inhabitants of the district of Ommoord who were aged 55 years and older were invited to participate. At baseline, in 1990-1993, of the 10,215 invited inhabitants, 7,983 agreed to participate in the baseline examination (response rate 78%). In 2000, the cohort was extended with 3,011 participants (67% of invitees). This extension consisted of all persons living in the study district who had become 55 years and older or had moved into the study district. A second extension was initiated in 2006, in which 3,932 participants (65% of invitees) who were 45 years and older were included. Study rounds consist of a home interview and visits with extensive investigations at the dedicated research centre. Rounds are repeated every 4-6 years. Participants are continuously monitored for diseases and mortality through linkage of the medical records from the general practitioners and municipality records. The Rotterdam Study has been approved by the Medical Ethics Committee of the Erasmus MC and by the Ministry of Health, Welfare and Sport of The Netherlands. All participants provided written informed consent to participate in the study and to obtain information from their treating physicians.
A total of 11,496 participants from the three subcohorts who were genotyped passed genotyping quality control (92% of all subjects with genotyping)[22]. Exclusion criteria were a call rate <98%, Hardy–Weinberg p-value $<10^{-6}$, minor allele frequency <0.01%, excess autosomal heterozygosity >0.336, sex mismatch, and outlying identity-by-state clustering estimates. Imputations were performed using the Haplotype Reference Consortium (HRC) panel[23].
Dementia ascertainment involved cognitive screening at the study research centre. We further assessed individuals with a Mini-mental state examination (MMSE) score <26 or Geriatric Mental State Schedule organic level >0[24], by administering the Cambridge Mental Disorders of the Elderly Examination by a research physician. We also interviewed spouses or informants. A consensus panel headed by a consultant neurologist established the final diagnosis according to standard criteria. We studied the outcomes of all-cause dementia

(DSM-III-R), and Alzheimer's disease (NINCDS–ADRDA). For the assessment of dementia, and type of dementia, the latest follow-up information with available data was used to determine the disease state. Follow-up for dementia was near complete until 1st January 2016. Within this period, participants were censored at date of dementia diagnosis, death, or loss to follow-up. For this study, we included participants from the first, second and third subcohort (N=11,070).

***European Alzheimer's Disease Initiative (EADI) Consortium***

EADI is composed of several case-control studies and one population-based cohort, the 3C study[25,26]. Case-control studies are comprised of AD cases and cognitively normal controls across France. The population-based cohort, the 3C study, is a prospective study of the relationship between vascular factors and dementia carried out in the three French cities Bordeaux, Montpellier and Dijon. The AD status was defined based on 12 years follow-up for Dijon participants, 14-15 years follow-up for Montpellier participants and 17-18 years follow-up for Bordeaux participants. All other non-demented subjects of 3C were included as controls. All AD cases, both in the case-control studies and the 3C study, were ascertained by neurologists and the clinical diagnosis of probable AD was established according to the DSM-III-R and NINCDS-ADRDA criteria. Samples that passed DNA quality control were genotyped with Illumina Human 610-Quad BeadChips.

***Genetic and Environmental Risk in AD (GERAD) Consortium/Defining Genetic, Polygenic, and Environmental Risk for Alzheimer's Disease (PERADES) Consortium***

The GERAD/PERADES sample comprises 3,177 Alzheimer's disease cases and 7,277 controls with available age and gender data[27]. Cases and elderly screened controls were recruited by the Medical Research Council (MRC) Genetic Resource for Alzheimer's disease (Cardiff University; Institute of Psychiatry, London; Cambridge University; Trinity College Dublin), the Alzheimer's Research Trust (ART) Collaboration (University of Nottingham; University of Manchester; University of Southampton; University of Bristol; Queen's University Belfast; the Oxford Project to Investigate Memory and Ageing (OPTIMA), Oxford University); Washington University, St Louis, United States; MRC PRION Unit, University College London; London and the South East Region Alzheimer's disease project (LASER-AD), University College London; Competence Network of Dementia (CND) and Department of Psychiatry, University of Bonn, Germany; the National Institute of Mental Health (NIMH) Alzheimer's disease Genetics Initiative. 6129 population controls were drawn from large existing cohorts with available GWAS data, including the 1958 British Birth Cohort (1958BC) (http://www.b58cgene.sgul.ac.uk), the KORA F4 Study and the Heinz Nixdorf Recall Study. All Alzheimer's disease cases met criteria for either probable (NINCDS-ADRDA, DSM-IV) or definite (CERAD) Alzheimer's disease. All elderly controls were screened for dementia using the MMSE or ADAS-cog, were determined to be free from dementia at neuropathological examination or had a Braak score of 2.5 or lower. Genotypes from all cases and 4617 controls were previously included in the AD GWAS by Harold and colleagues[27]. Genotypes for the remaining 2660 population controls were obtained from WTCCC2.

***The Norwegian DemGene Network***

This is a Norwegian network of clinical sites collecting cases from memory clinics based on a standardized examination of cognitive, functional, and behavioral measures and data on the progression of most patients. The Norwegian DemGene Network includes 2,224 cases and 3,089 healthy controls from different studies described elsewhere[28]. The cases were diagnosed according to recommendations from the NIA–AA, the NINCDS–ADRDA criteria, or the ICD-10 research criteria. The controls were screened with a standardized interview and cognitive tests. Additional controls from blood donors of the Oslo University Hospital, Ulleval Hospital, were included ($n$=4992, age between 18-65 years, 48% female). They were thoroughly screened for diseases and medication, and provided blood for DNA analysis, in line with approval from the Regional Committee for Medical and Health Research Ethics. Individuals from the DemGene study and blood donors were genotyped using either the

Human Omni Express-24 v1.1 chip (Illumina Inc., San Diego, CA) or the DeCodeGenetics_V1_20012591_A1 chip at deCODE Genetics (Reykjavik, Iceland).

***<u>The Neocodex–Murcia study (NxC)</u>***

This study includes 324 sporadic AD patients and 754 controls of unknown cognitive status from the Spanish general population collected by Neocodex[29,30]. AD patients were diagnosed as having possible or probable AD in accordance with the NINCDS–ADRDA criteria.

***<u>Bonn studies</u>***

<u>DietBB</u>: The DietBB sample included in this GWAS is a subsample extracted from the AgeCoDe cohort in the context of an ongoing genome-wide methylation analysis for dementia. In addition to methylation, the DietBB samples has genome-wide genotype data which was included in this study. The German study on aging, cognition and dementia (AgeCoDe)[10,31] study is a general practice (GP) registry-based longitudinal study in elderly individuals on the identification of predictors of dementia. Participants were recruited in six German cities (Bonn, Dusseldorf, Hamburg, Leipzig, Mannheim, and Munich) with a total of 138 GPs connected to the study sites. The inclusion criteria for this study were an age of 75 years and older, absence of dementia according to GP judgment, and at least one contact with the GP within the past 12 months. Exclusion criteria were GP consultations by home visits only, living in a nursing home, severe illness with an anticipated fatal outcome within 3 months, language barrier, deafness or blindness, and lack of ability to provide informed consent. Baseline recruitment was performed in 2002 and 2003. The study was approved by the local ethical committees of the Universities of Bonn, Hamburg, Dusseldorf, Heidelberg/Mannheim, and Leipzig, and the Technical University of Munich. A total of 3327 subjects provided informed consent for participation after being provided with a complete description of the study protocol. The study assessments were performed by trained interviewers at the subjects' home. Seventy individuals were excluded after baseline interview because of the presence of dementia according to standard assessment, and 40 subjects were excluded for age less than 75 years. In AgeCoDe, dementia was diagnosed according to the criteria set of DSM-IV in a consensus conference with the interviewer and an experienced geriatrician or geriatric psychiatrist. The etiological diagnosis of dementia in AD was established according to the National Institute of Neurological and Communicative Diseases and Stroke/Alzheimer's Disease and Related Disorders Association (NINCDS-ADRDA) criteria for probable AD[8]. Mixed dementia was diagnosed in cases of cerebrovascular events without temporal relationship to cognitive decline. Mixed dementia and dementia in AD were combined. Dementia diagnosis in subjects who were not interviewed personally was based on the Global Deterioration Scale[32] (score ≥4 points). In these cases, an etiological diagnosis was established only if the information provided was sufficient to judge etiology according to the criteria just described. For DietBB, cohort participants where included if they were dementia-free at baseline and available biomaterial for DNA analysis is available. This criterion led to the selection of 320 participants. In 120 of these participants, dementia of the AD-type occurred at any follow up. The additional 200 remain free of dementia until last follow up of AgeCoDe.

<u>Bonn OMNI cohort</u>: the Bonn OMNI cohort consists of AD patients and controls derived from a larger German GWAS cohort which was recruited from the following three sources: (i) the German Dementia Competence Network; (ii) the German study on Aging, Cognition, and Dementia in primary care patients (AgeCoDe); and (iii) the interdisciplinary Memory Clinic at the University Hospital of Bonn. The control sample comprised of individuals from the population-based study Heinz Nixdorf Recall (HNR) study cohort. This sample was previously used for replication in Lambert et al.[33]. *The German study on aging, cognition and dementia (AgeCoDe)*: see description above. *The German competence network cohort (DCN)*: The DCN cohort includes 1,095 patients with mild cognitive impairment (MCI) and 648 cases with mild Alzheimer's disease (AD) clinical dementia syndrome that were recruited from 14 university hospital memory clinics across Germany between 2003 and 2005[9]. Exclusion criteria were substance abuse or dependence, insufficient German language skills, multi-morbidity, comorbid condition with excess mortality, circumstances that would have made regular

attendance at follow-up visits questionable and lack of an informant. The diagnosis of mild dementia according to ICD-10 criteria required a decline of cognitive ability (at least 1 SD) from a previous level in at least 2 domains as evidenced by age-corrected standardized tests, impairment in activities of daily living (i.e. B-ADL > 6), changes in personality, drive, social behavior or control of emotion but no clouding of consciousness. These changes must have persisted for at least 3 months. The etiological diagnosis of AD was assigned according to NINCDS-ADRDA criteria[8]. *Memory clinic Bonn*: The interdisciplinary Memory Clinic of the Department of Psychiatry and Department of Neurology at the University Hospital in Bonn provided further patients. Diagnoses were assigned according the NINCDS/ADRDA criteria[8] and on the basis of clinical history, physical examination, neuropsychological testing (using the CERAD neuropsychological battery, including the MMSE), laboratory assessments, and brain imaging. *Control sample:* In the Heinz Nixdorf Recall (Risk Factors, Evaluation of Coronary Calcification, and Lifestyle) study, participants were randomly sampled in three cities in Germany. The study design has previously been described[34,35]. Briefly, 4814 participants aged 45 to 75 years were enrolled between 2000 and 2003 (t0, baseline). Cognitive performance of participants was evaluated at follow up scheduled 5 years after baseline (t1, n = 4157, 2005–2008) and then again at follow up 5 years after t1 (t2, n = 3087, 2010–2015). Controls sample was selected if participant did not present cognitive impairment as reported at the last available evaluation. Cognitive evaluation has been described extensively previously[36,37]. Herein, cognitive impairment at t1 was defined as a performance of one standard deviation (SD) below the age- and education-adjusted mean except for the clock-drawing test, where a performance ≥3 was rated as impaired (for a detailed description, see the study by Winkler et al.[36]). The study was approved by the University of Duisburg-Essen Institutional Review Board and followed established guidelines of good epidemiological practice.

***Sub Saharan Africa (EPIDEMCA)***

Between 2011 and 2012, a population-based study of dementia prevalence and its associated factors in Central Africa (EPIDEMCA) was carried out37. Around 2,000 participants aged 65 and above living in Central African Republic (CAR) and Republic of Congo (ROC) were interviewed. The full protocol of the EPIDEMCA study (including data collection, variable definition and dementia diagnosis) has been described elsewhere[17]. An annual follow-up (EPIDEMCA-FU) was carried out over 3 years (2013-2015) only in ROC. Dementia diagnosis was made according to the DSM-IV criteria and AD was diagnosed according to the clinical criteria proposed by the NINCDS-ADRDA (National Institute of Neurological and Communicative Disorders and Stroke and the AD and Related Disorders Association). The severity of dementia was evaluated by the CDR (Clinical Dementia Rating Scale). Mild cognitive impairment (MCI) was diagnosed according to Petersen criteria and MCI cases were excluded from controls. An experienced neurologist reviewed all clinical assessements, evaluation of dependence (Instrumental Activities of Daily Living adapted to the African context, leisure, other social and occupational activities) , and test performances (the Free and Cued Selective Reminding Test (oral version with image), Zazzo's cancellation task and Isaac's Set Test of verbal fluency) for all the participants assessed at the second stage of the baseline study / during follow-up studies to reach a consensus. In case of insufficient information to reach a diagnosis, the participant was coded unclassifiable. These samples were genotyped using the ILLUMINA GSA array in the EADB-France node. Ethics committees, "Comité d'Ethique de la Recherche en Sciences de Santé" in ROC (reference: 00000204/DGRST/CERSAA for EPIDEMCA and reference: 0000200/MRSIT/DGRST/CERSSA for EPIDEMCA-FU), as well as the "Comité de la Protection des Personnes Sud-Ouest Outre-Mer" in France (SOOM4-CE-3) approved the study protocol.

***Tunisia***

DNA samples from 254 subjects (58 AD patients and 196 healthy controls) were recruited from the Department of Neurology, University Hospital of Fattouma Bourguiba, Monastir, Tunisia; the Department of Neurology, University Hospital of Mahdia, Tunisia; and the Alzheimer Family

Assistance Center in Tunisia (AFA center) as previously described[38]. Written informed consent was obtained from all participants or their representatives according to the ethical standards of the Ethics Committee for Research in Life Sciences and Health of the ISBM (CER-SVS/ISBM) and the 1964 Helsinki Declaration. For each participant, age, demographic information, and family and medical histories were collected. All participants were unrelated over three generations. In addition, for AD patients, a detailed clinical assessment was carried out, including a Mini Mental State Examination (MMSE), a standard brain MRI, neuropsychological tests, and laboratory studies. All AD patients were included after at least 2 years of follow-up. Diagnoses of AD were made according to the criteria of the National Institute of Neurological and Communicative Disorders and Stroke and the Alzheimer's Disease and Related Disorders Association (NINCDS–ADRDA) for possible or probable AD. Controls were selected after passing an MMSE test and according to the following criteria: no family member with AD or any type of dementia or psychiatric disorder, a mentally healthy status, and no history of alcohol or drug use. This retrospective chart study involving human participants was in accordance with the ethical standards of the Ethics Committee for Research in Life Sciences and Health of the ISBM (CER-SVS/ISBM) and with the 1964 Helsinki Declaration and its later amendments or comparable ethical standards. All procedures performed in the present work were approved by Ethics Committee for Research in Life Sciences and Health of the ISBM (CER-SVS/ISBM), Monastir, Tunisia. The Axiom 815K Spanish biobank arrays (Thermo Fisher) at the Spanish National Centre for Genotyping (CeGEN, Santiago de Compostela, Spain) were used for genotyping.

***Argentina.***

The Argentinian samples were recruited in the context of the Alzheimer's Genetics in Argentina – ALZheimer ARgentina consortium (AGA-ALZAR) from the following centers: Medical Research Institute A. Lanari (C1427ARO, Buenos Aires City), Hospital de Clínicas José de San Martín (C1120AAF, Buenos Aires City), Hospital HIGA-Eva Perón (B1650NBN, General San Martín), Hospital El Cruce (B1888AAE, Florencio Varela), and several geriatric centers across Jujuy and Mendoza provinces, organized and coordinated by their respective Public Ministry of Health. The study (protocol CBFIL#22) was approved by the ethical committee (HHS IRB#00007572, IORG#006295, FWA00020769), and all participants and/or family members gave their informed consent. Diagnosis of AD followed diagnostic criteria from the National Institute of Neurological and Communicative Disorders and Stroke and the Alzheimer's disease and Related Disorders Association (NINCDS-ADRDA). Peripheral blood or saliva samples were processed to obtain DNA using the QIAmp DNA mini kit (Qiagen) and genotyped using the Illumina Infinium Global Screening Array (GSA) v.1.0 combined to a GSA shared custom content[39].

***Chile***

The Chilean samples recruited correspond to patients with AD and control subjects, from different studies. Control individuals were recruited from Alexandros longitudinal study[40] of community dwelling older adults (≥60 years old) of different demographic origin and socioeconomic level, mainly in the study of healthy life expectancy, free of disability and dementia. All participants were randomly selected from 18 Primary Health Care Centers and signed an informed consent on enrolment after they had received written and verbal information about the study. The ethical committee of Institute of Nutrition and Food Technology (INTA), University of Chile (Acta 23, 2012) approved the study protocol (FONDECYT nº1130947). Cognitive status was determined through the Mini-mental State Examination (MMSE) test with a cut-off point of 21/22, previously validated in Chile[41]. AD patients were recruited at Biomedica Research Group in Santiago. Subjects were comprehensively studied and diagnosed following the NINCDS-ADRDA criteria for AD. The GWAS study was approved by the Ethics Committe "Servicio de Salud Metropolitano Oriente" (SSMO). Additional AD cases and control individuals (32 AD and 20 controls) from Santiago were recruited from the GERO[42] (Geroscience Center for Brain Health and Metabolism) study at the Memory and Neuropsychiatric Center of the Hospital del Salvador and Faculty of

Medicine of the University of Chile. The FONDAP GERO project n°15150012 was also approved by the Ethics Committee of the SSMO. A total of 934 samples (n=800 DNA and n=134 frozen blood) were sent to Ace Alzheimer Center Barcelona (Barcelona, Spain) for processing. DNA was extracted from peripheral blood according to standard procedures using the Chemagic system (Perkin Elmer). For the starting DNA samples, a re-extraction protocol using the Chemagic system was also followed in order to purify the DNA samples. Only samples reaching DNA concentrations of >10 ng/µL and presenting high integrity were included for genotyping. Finally, AD cases (n=123) and controls (n=252) were randomized across sample plates to avoid batch effects. The Axiom 815K Spanish biobank arrays (Thermo Fisher) at the Spanish National Centre for Genotyping (CeGEN, Santiago de Compostela, Spain) were used for genotyping.

***Brazil***

Individuals were recruited at the geriatric outpatient clinic, University Hospital of the Federal University of Minas Gerais (UFMG). This Clinic serves as a Reference Center for the diagnosis and treatment of dementia as well as the diagnosis and treatment of frail older adults. The study was approved by the local ethics committee, and written informed consent was obtained from all the participants prior to inclusion. All participants underwent the same research protocol, which consisted of assessment of sociodemographic characteristics, medical comorbidities, medication history and lifestyle habits as well as memory complaints. The cognitive assessment protocol included the following instruments: MMSE, Clinical Dementia Rating Scale, Verbal Fluency Test, CERAD battery word list, Brief Cognitive Screening, clock drawing test, Neuropsychiatric Inventory, functional activities questionnaire, diagnostic criteria for depression (DSM-5), Geriatric Depression Scale 15 items version (GDS).

The neuropsychological battery used was previously validated for assessing older adults with low formal education[43]. All tests were validated for use with Brazilian elderly and cutoff points were considered according to their education level. Patients also underwent routine blood tests (e.g., hematology, biochemistry, thyroid-stimulating hormone, vitamin B12 and folate levels, syphilis, and HIV serology) and neuroimaging studies (computerized tomography or nuclear magnetic resonance). The inclusion criteria consisted of older adults aged 60 or over, with a clinical diagnosis of probable Alzheimer's disease dementia and cognitively unimpaired older adults who agreed to take part in the project and signed the consent form. The exclusion criteria included cognitive impairment due to causes other than AD dementia, patients with moderate to severe mental disorders, hydrocephalus and intracranial mass documented by CT scan or NMR, abnormalities in serum folate and/or vitamin B12, serology for syphilis or thyroid hormone levels, and individuals with severe sensory impairment, that made it difficult to administer and respond to the cognitive assessment instruments and those who failed to attend the appointment for the application of the clinical and cognitive protocols. The samples were genotyped by Life & Brain (Bonn, Germany) using the Illumina Infinium Global Screening Array (GSA) v.1.0.

***Colombia***

The study sample consisted of 241 AD patients and 254 control subjects for a total of 495 individuals from the cundiboyacense region of Colombia. All of the participants were recruited and evaluated at neurology and neuropsychology services by the Neuroscience Group of the National University of Colombia. Diagnosis of AD was determined following the criteria established by the National Institute of Neurological and Communicative Disorders and Stroke and the Alzheimer's Disease and Related Disorders Association (NINCDS-ADRDA). The control group consisted of individuals with no personal or family history of neurodegenerative and/or psychiatric diseases. All patients and controls signed an informed consent to participate in the study.

***ADSP.*** Alzheimer's Disease Sequencing Project (ADSP) is a collaborative project aiming at identifying new variants, genes, and therapeutic targets in AD[44]. The joint-called whole-genome sequencing data from the Alzheimer's Disease Sequencing Project (ADSP) accession id NG00067.v10 were downloaded from the NIAGADS DSS website (https://dss.niagads.org/datasets/ng00067/), with 36,361 whole-genome genomes[45]. The global ancestry of each individual genome was determined with SNPweights v.2.1[46] using a set of ancestry-weighted variants computed on reference populations from the 1000 Genomes Project as in[47]. By applying a global ancestry percentage cutoff of >75%, the samples were assigned to the 5 super-populations: South-Asians, East-Asians, Americans, Africans, and Europeans, and classified as Admixed ancestries otherwise. The 83 independent variants were extracted from the individual genomes with vcftools v0.1.17[48]. Technical duplicates or twins were identified using KING v2.2.5 (Kinship-based INference for Gwas)[49] and only one genome among the set of identical pairs was retained for further analysis. The phenotype cognitively unimpaired or Alzheimer's disease case was obtained from the ADSP, and participants who did not fall in either of these two categories were excluded from further analysis (see cohorts descriptions at https://dss.niagads.org/datasets/ng00067/). Principal components accounting for genetic ancestry were computed per ancestry group using smartpca[50].

***Japan***

Participants of Japanese ancestry were recruited by the Japanese Genetic Study Consortium of Alzheimer's Disease[52] composed of 48 medical institutes and the Japanese Alzheimer's Disease Neuroimaging Initiative[53] composed of 38 clinical sites across Japan. Probable AD cases were ascertained on the basis of the criteria of the National Institute of Neurological and Communicative Disorders, and Stroke-Alzheimer's Disease and Related Disorders (NINCDS/ADRDA). The Mini-Mental State Examination, Clinical Dementia Rating, and/or Function Assessment Staging were primarily used for evaluation of cognitive impairment. Elders living in an unassisted manner in the local community with no signs of dementia were used as controls. DNA was extracted from peripheral blood leukocytes using standard protocols. GWAS genotyping was performed using Illumina Infinium Asian Screening Array.

***Korea (GARD)***

Participants of Korean ancestry were recruited from 34 referral hospitals in the Republic of Korea. Participants were diagnosed with AD dementia based on detailed neuropsychological test results[54]. The participants' diagnoses was those of at the latest assessment point. AD dementia was defined in accordance with the core clinical criteria for probable AD dementia according to the National Institute on Aging-Alzheimer Association. Participants were excluded when they had (1) a causative genetic mutation for AD in known genes, such as PSEN1, PSEN2, or APP; (2) structural abnormalities detected on brain magnetic resonance imaging, such as severe cerebral ischemia, territorial infarction, or brain tumors; or (3) other medical or psychiatric diseases that may cause cognitive impairment. DNA samples were genotyped using the Asian screening array (ASA) chip (Illumina). A subset of 125 samples was genotyped using a customized Korea Biobank array (KBA) chip (Affymetrix). Quality control for SNV data was conducted using the PLINK software and imputation was conducted using the Minimac4 software at the University of Michigan Imputation Server[54].

***CHINA***

The Chinese AD WGS cohort (N = 1239 comprising 453 patients with AD and 786 HCs) have been previously described[55]. Patients with AD were diagnosed according to the Diagnostic and Statistical Manual of Mental Disorders, Fifth Edition (DSM-5). The phenotypic data of the participants analyzed in this study were based on the participants' most recent diagnostic records (as of December 2019). The study was approved by the Clinical Research & Ethics Committees of Joint Chinese University of Hong Kong-New Territories East cluster for Prince of Wales Hospital (CREC Ref no. 2015.461), Kowloon Central Cluster/Kowloon East Cluster for Queen Elizabeth Hospital (KC/KE-15-0024/FR-3), and Human Participants Research Panel of the Hong Kong University of Science and Technology (CRP#180 and CRP#225). All participants provided written informed consent for both study participation and sample collection. We sent genomic DNA to Novogene for library preparation and WGS. The genomic

DNA libraries were sequenced using an Illumina NovaSeq 6000 System, which generated 150-bp paired-end reads. We performed quality control by examining the G/C content and per-base sequencing quality statistics. We subjected raw sequencing data to fastp for quality control, read trimming, and filtration[56]. We used BWA-mem to align clean reads to the GRCh37 human reference genome and then subjected the reads to GATK for duplicate removal and base quality score recalibration[57,58]. We performed variant calling and filtering of SNPs and INDELs by using GATK's germline joint calling as well as variant quality score recalibration with 95% and 90% sensitivity-level cutoffs for SNPs and INDELs, respectively7. Finally, we subjected filtered SNPs and INDELs to Beagle 4.0 for genotype refinement[59].
After variant calling, we performed standard quality control analysis. All participants showed consistent sex according to their genotypes and phenotypes. We calculated relatedness by performing KING analysis using plink2 and applied a kinship coefficient cutoff of 0.08 to remove second-degree relatives[60]. We performed principal component analysis and removed outliers according to the bigsnpr instruction manual[61] After sample-level quality control, we further filtered genetic variants by Hardy–Weinberg equilibrium test P-values >1 × 10−6.

***MVP***

The Million Veteran Program is a United States Office of Veterans Affairs (VA) Office of Research and Development-funded effort to identify factors that impact the health of US Veterans[62]. MVP volunteers are US Veterans who donate a blood sample for genetic analysis, agreed to have their electronic medical record (EMR) accessed for research purposes and in many cases complete surveys covering a wide range of health conditions and lifestyle factors[63]. Genome-wide genotype data are currently available for approximately 650K MVP participants (MVP Phase 4 genotype release). The generation of MVP genotype data for MVP participants is described in detail elsewhere[64]. Briefly, genotypes were assessed using the custom-designed MVP 1.0 custom Axiom array. QC procedures include relatedness, sex-concordance checks, and advanced marker filtering. Imputation was performed using TOPMed reference data. The ancestry of MVP participants was determined using the harmonized ancestry and race/ethnicity (HARE) method[65], which is similar to other genotype-based ancestry calling methods, except that concordance is checked between self-report and genetically inferred ancestry, and those with discrepant ancestry groups are not assigned a HARE category. Within-group PCs for ancestry were computed using FlashPCA25. Similar to several recent MVP publications[66], we utilized an International Classification of Diseases (ICD)-code-based method for classifying subjects as being AD, AD and related dementias (ADRD), or all-cause Dementia cases and controls. MVP participants were identified as a case at one of these levels if they had two or more qualifying ICD codes, with the first occurring after age 60. Controls were drawn from MVP participants age 65 or greater who had no ICD codes for any type of dementia or mild cognitive impairment, had not been prescribed AD medication, and who did not report AD/dementia in either parent on the MVP Baseline survey[67].

***American CSF samples***

Samples included in this study are from 23 different American cohorts[68]. Genotyping QC was performed within each cohort and involved stringent filtering criteria as previously described[69]. Prior to analysis, all related samples were removed, and only non-Hispanic white population were kept. In total, 7229 samples were included of which 3701 (51.20%) were males. The average of samples included was 68.67 (±11.39) years.

**Supplementary Figure 1:** GRS distribution (Finland, Sweden, Norway, Denmark, UK, Netherlands)

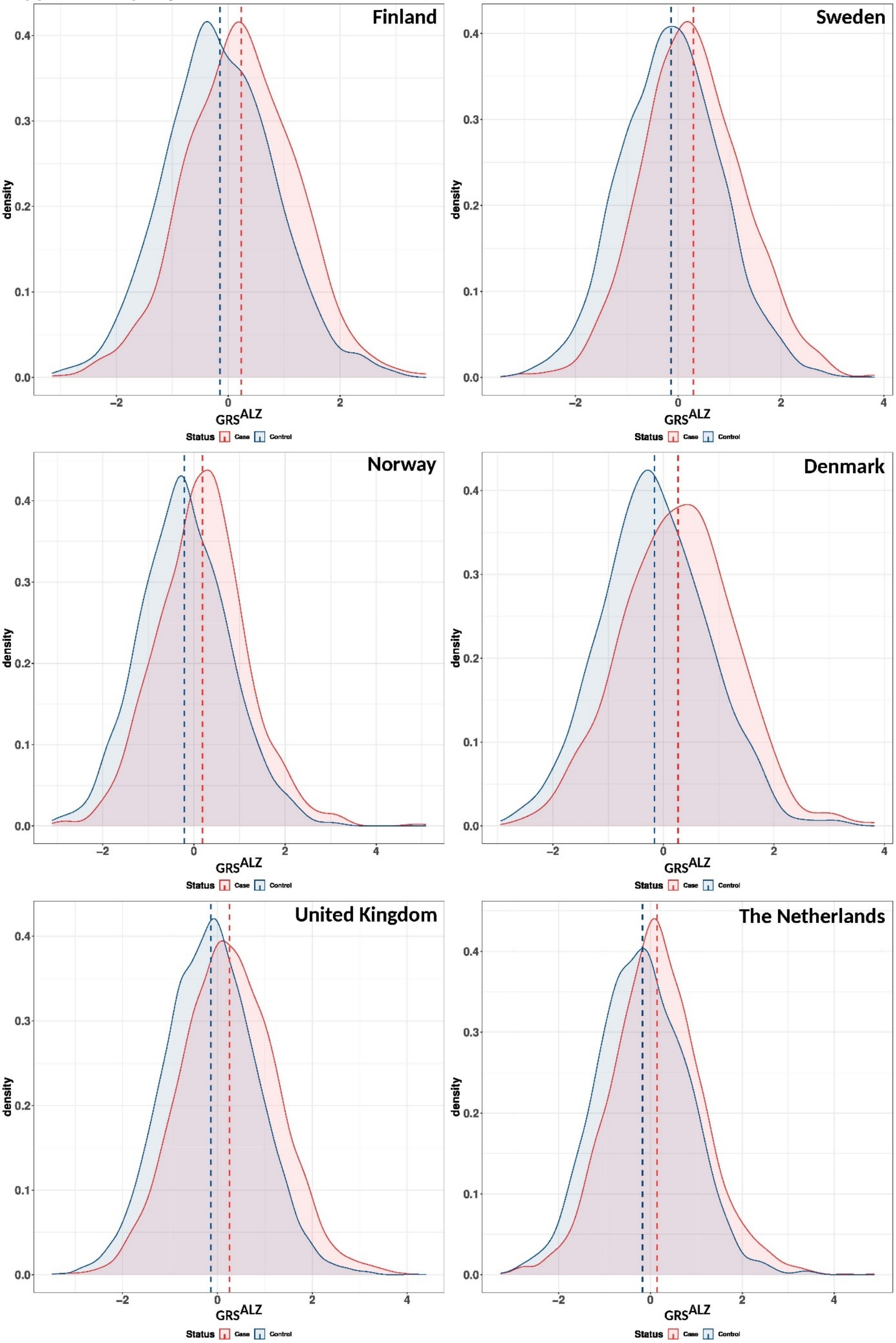

**Supplementary Figure 2:** GRS distribution (Belgium, Germany, Switzerland Czech Republic, Greece, Italy)

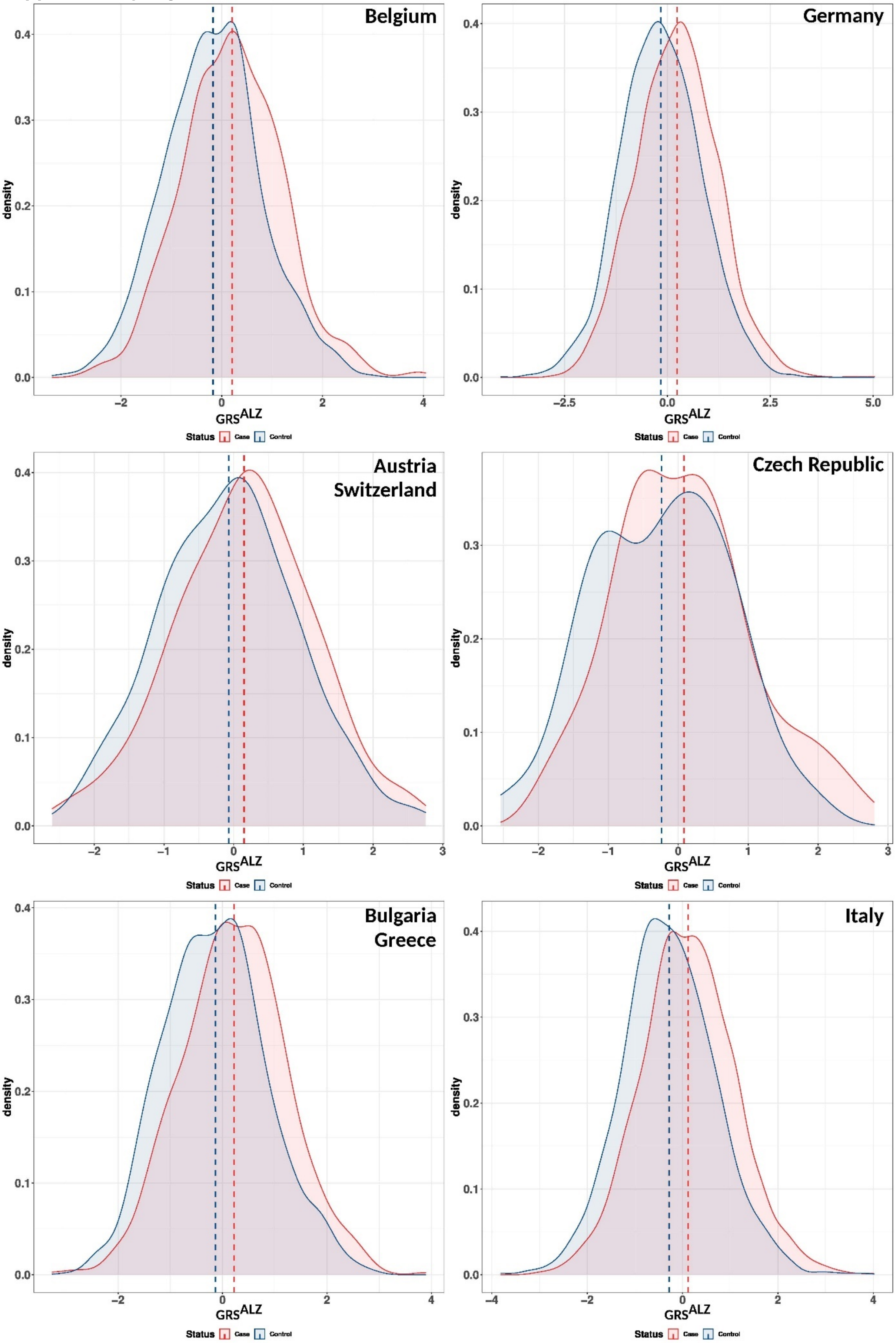

**Supplementary Figure 3:** GRS distribution (France, Spain, Portugal)

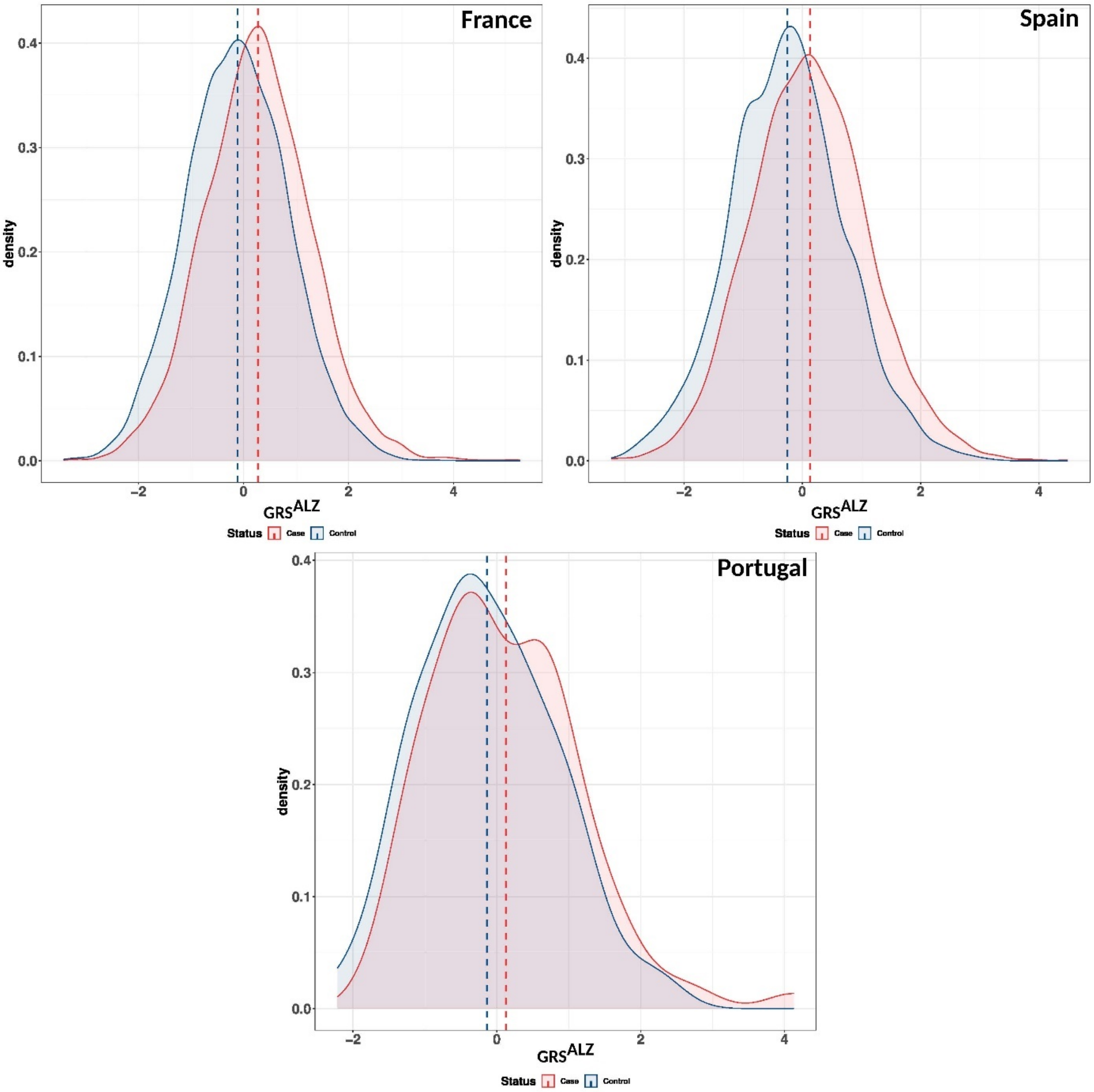

**Supplementary Figure 4:** GRS$^{ALZ}$ association with AD risk across Europe. Ncase, number of cases; Ncontrols, number of controls; OR, Odds ratio. The lines in the Forrest plots indicate the 95% confidence interval for the ORs.

GRS$^{ALZ}$

$I^2$ = 14.4 , HetP = 2.93e-01

| Country | Ncases | Ncontrols | OR | 95% CI | P-value |
|---|---|---|---|---|---|
| Finland | 1,118 | 1,770 | 1.44 | [1.33-1.56] | 3.14e-18 |
| Norway | 1,327 | 1,179 | 1.56 | [1.44-1.71] | 6.07e-24 |
| Sweden | 1,466 | 3,078 | 1.60 | [1.50-1.72] | 1.60e-42 |
| Denmark | 398 | 650 | 1.57 | [1.36-1.81] | 5.29e-10 |
| United Kingdom | 4,622 | 8,414 | 1.55 | [1.48-1.61] | 8.78e-85 |
| The Netherlands | 2,428 | 2,024 | 1.48 | [1.38-1.58] | 3.80e-28 |
| Belgium | 1,211 | 1,431 | 1.55 | [1.41-1.72] | 1.58e-18 |
| Germany | 2,190 | 3,141 | 1.53 | [1.44-1.63] | 5.71e-43 |
| Austria/Switzerland | 175 | 372 | 1.26 | [1.04-1.53] | 1.66e-02 |
| Czech Republic | 177 | 60 | 1.56 | [1.08-2.29] | 1.97e-02 |
| Bulgaria/Greece | 735 | 1,195 | 1.38 | [1.20-1.61] | 1.44e-05 |
| Italy | 2,973 | 1,251 | 1.53 | [1.43-1.65] | 2.52e-31 |
| France | 3,789 | 9,026 | 1.50 | [1.44-1.57] | 3.19e-85 |
| Spain | 3,103 | 1,615 | 1.64 | [1.53-1.76] | 1.48e-41 |
| Portugal | 80 | 74 | 1.31 | [0.94-1.85] | 1.14e-01 |
| **Meta-analysis** | **25,792** | **35,280** | **1.53** | **[1.50-1.56]** | **2.60e-412** |

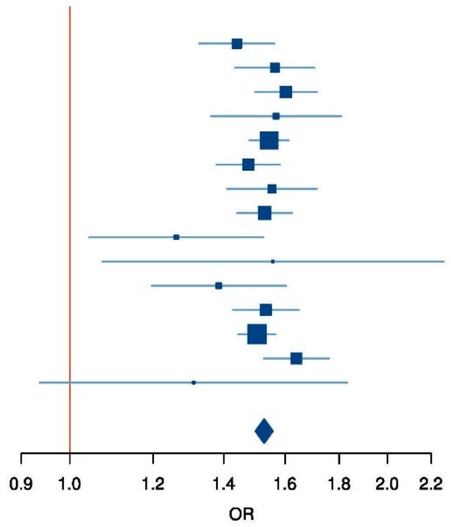

**Supplementary Figures 5:** calibrated GRS$^{ALZ}$ association with AD risk across Europe. Ncase, number of cases; Ncontrols, number of controls; OR, Odds ratio. The lines in the Forrest plots indicate the 95% confidence interval for the ORs.

**A**

calibrated GRS$^{ALZadjAPOE}$

$I^2$ = 18.6 , HetP = 2.45e-01

| Country | Ncases | Ncontrols | OR | 95% CI | P-value |
|---|---|---|---|---|---|
| Finland | 1,118 | 1,770 | 1.42 | [1.30-1.55] | 1.21e-14 |
| Norway | 1,327 | 1,179 | 1.55 | [1.41-1.70] | 1.91e-19 |
| Sweden | 1,466 | 3,078 | 1.62 | [1.51-1.75] | 2.19e-37 |
| Denmark | 398 | 650 | 1.53 | [1.32-1.78] | 3.11e-08 |
| United Kingdom | 4,622 | 8,414 | 1.53 | [1.46-1.60] | 6.32e-73 |
| The Netherlands | 2,428 | 2,024 | 1.46 | [1.36-1.57] | 4.96e-24 |
| Belgium | 1,211 | 1,431 | 1.57 | [1.42-1.75] | 2.28e-17 |
| Germany | 2,190 | 3,141 | 1.51 | [1.42-1.62] | 1.60e-36 |
| Austria/Switzerland | 175 | 372 | 1.24 | [1.03-1.51] | 2.70e-02 |
| Czech Republic | 177 | 60 | 1.76 | [1.17-2.72] | 8.55e-03 |
| Bulgaria/Greece | 735 | 1,195 | 1.39 | [1.20-1.62] | 1.41e-05 |
| Italy | 2,973 | 1,251 | 1.52 | [1.41-1.63] | 2.05e-27 |
| France | 3,789 | 9,026 | 1.49 | [1.43-1.56] | 7.69e-73 |
| Spain | 3,103 | 1,615 | 1.61 | [1.49-1.74] | 1.75e-35 |
| Portugal | 80 | 74 | 1.25 | [0.89-1.78] | 2.12e-01 |
| **Meta-analysis** | **25,792** | **35,280** | **1.51** | **[1.48-1.55]** | **1.69e-351** |

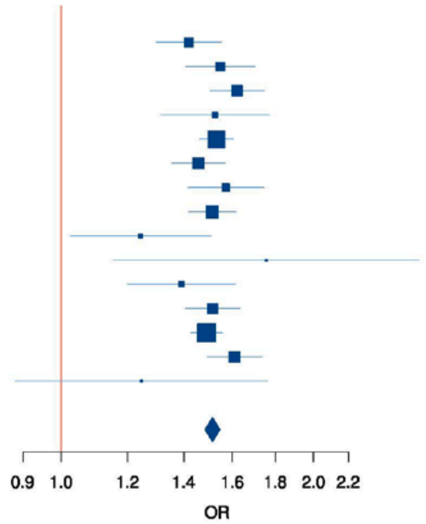


**A**

calibrated GRS$^{ALZ}$

$I^2$ = 11.7 , HetP = 3.22e-01

| Country | Ncases | Ncontrols | OR | 95% CI | P-value |
|---|---|---|---|---|---|
| Finland | 1,118 | 1,770 | 1.45 | [1.33-1.57] | 1.32e-18 |
| Norway | 1,327 | 1,179 | 1.56 | [1.43-1.70] | 2.01e-23 |
| Sweden | 1,466 | 3,078 | 1.60 | [1.50-1.72] | 1.25e-42 |
| Denmark | 398 | 650 | 1.56 | [1.36-1.80] | 7.45e-10 |
| United Kingdom | 4,622 | 8,414 | 1.54 | [1.48-1.61] | 1.78e-84 |
| The Netherlands | 2,428 | 2,024 | 1.48 | [1.38-1.59] | 2.59e-28 |
| Belgium | 1,211 | 1,431 | 1.56 | [1.41-1.72] | 1.50e-18 |
| Germany | 2,190 | 3,141 | 1.53 | [1.44-1.62] | 9.93e-43 |
| Austria/Switzerland | 175 | 372 | 1.25 | [1.04-1.52] | 2.10e-02 |
| Czech Republic | 177 | 60 | 1.52 | [1.05-2.24] | 3.08e-02 |
| Bulgaria/Greece | 735 | 1,195 | 1.39 | [1.20-1.61] | 1.22e-05 |
| Italy | 2,973 | 1,251 | 1.54 | [1.43-1.65] | 2.13e-31 |
| France | 3,789 | 9,026 | 1.50 | [1.44-1.57] | 5.36e-85 |
| Spain | 3,103 | 1,615 | 1.63 | [1.52-1.75] | 1.05e-40 |
| Portugal | 80 | 74 | 1.29 | [0.93-1.82] | 1.40e-01 |
| **Meta-analysis** | **25,792** | **35,280** | **1.53** | **[1.50-1.56]** | **1.29e-410** |

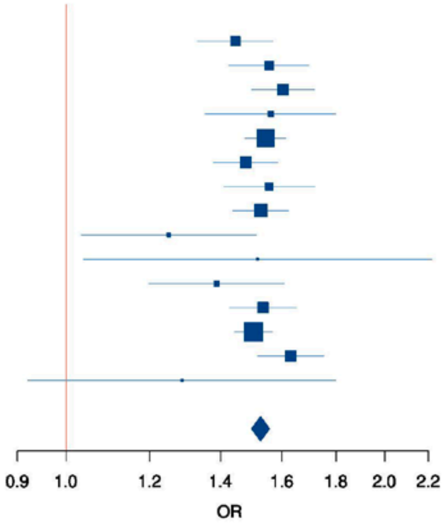

**Supplementary Figure 6:** GRS$^{ALZ}$ association with AD risk in Men and Women. Ncase, number of cases; Ncontrols, number of controls; OR, Odds ratio. The lines in the Forrest plots indicate the 95% confidence interval for the ORs.

GRS$^{ALZ}$

Females : $I^2$ = 20.4 , HetP = 2.26e-01 ; Males : $I^2$ = 0 , HetP = 7.60e-01

| Country | Gender | Ncases | Ncontrols | OR | 95% CI | P-value |
|---|---|---|---|---|---|---|
| Finland | Females | 716 | 914 | 1.40 | [1.26-1.56] | 8.12e-10 |
| | Males | 402 | 856 | 1.51 | [1.33-1.72] | 2.28e-10 |
| Norway | Females | 840 | 622 | 1.54 | [1.37-1.73] | 2.01e-13 |
| | Males | 487 | 557 | 1.61 | [1.41-1.85] | 2.98e-12 |
| Sweden | Females | 926 | 1,898 | 1.65 | [1.52-1.80] | 5.13e-30 |
| | Males | 540 | 1,180 | 1.52 | [1.37-1.70] | 3.26e-14 |
| Denmark | Females | 227 | 354 | 1.66 | [1.38-2.01] | 1.15e-07 |
| | Males | 171 | 296 | 1.45 | [1.17-1.81] | 9.21e-04 |
| United Kingdom | Females | 2,633 | 4,323 | 1.54 | [1.45-1.64] | 3.47e-46 |
| | Males | 1,989 | 4,091 | 1.55 | [1.45-1.65] | 4.82e-40 |
| The Netherlands | Females | 1,354 | 885 | 1.45 | [1.31-1.61] | 2.06e-12 |
| | Males | 1,074 | 1,139 | 1.50 | [1.36-1.64] | 4.17e-17 |
| Belgium | Females | 783 | 890 | 1.55 | [1.37-1.76] | 1.23e-11 |
| | Males | 428 | 541 | 1.56 | [1.33-1.83] | 2.96e-08 |
| Germany | Females | 1,347 | 1,755 | 1.55 | [1.44-1.68] | 2.16e-27 |
| | Males | 843 | 1,386 | 1.50 | [1.37-1.65] | 3.07e-17 |
| Austria/Switzerland | Females | 112 | 204 | 1.23 | [0.96-1.58] | 1.00e-01 |
| | Males | 63 | 168 | 1.33 | [0.99-1.81] | 6.53e-02 |
| Czech Republic | Females | 107 | 40 | 1.60 | [1.00-2.64] | 5.56e-02 |
| | Males | 70 | 20 | 1.50 | [0.85-2.82] | 1.76e-01 |
| Bulgaria/Greece | Females | 445 | 676 | 1.50 | [1.24-1.82] | 4.05e-05 |
| | Males | 290 | 519 | 1.22 | [0.97-1.54] | 8.84e-02 |
| Italy | Females | 2,015 | 715 | 1.53 | [1.39-1.67] | 2.64e-19 |
| | Males | 958 | 536 | 1.58 | [1.40-1.77] | 2.65e-14 |
| France | Females | 2,383 | 5,540 | 1.47 | [1.39-1.54] | 9.79e-49 |
| | Males | 1,406 | 3,486 | 1.58 | [1.47-1.69] | 1.04e-38 |
| Spain | Females | 2,076 | 1,027 | 1.60 | [1.46-1.75] | 1.47e-24 |
| | Males | 1,027 | 588 | 1.65 | [1.46-1.89] | 1.99e-14 |
| Portugal | Females | 60 | 56 | 1.07 | [0.74-1.56] | 7.27e-01 |
| | Males | 20 | 18 | 3.10 | [1.28-10.09] | 2.70e-02 |
| **Meta-analysis** | **Females** | **16,024** | **19,899** | **1.52** | **[1.48-1.56]** | **3.65e-235** |
| | **Males** | **9,768** | **15,381** | **1.54** | **[1.49-1.59]** | **8.62e-175** |

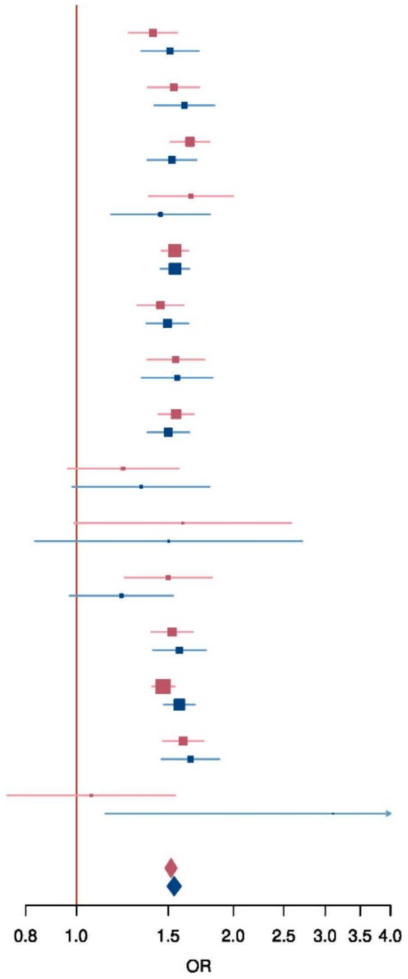

**Supplementary Figure 7:** Association of GRS$^{alz}$ with the level of Aβ42, Tau and p-TAU in the cerebrospinal fluid in European-ancestry populations. Ncase, number of cases; Ncontrols, number of controls, OR, Odds ratio. The lines in the Forrest plots indicate the 95% confidence interval for the ORs. If HetP <0.05, random-effect is shown for the meta-analysis results.

**A**

$A\beta_{42}$ ($PGS^{ALZ}$)

Europe : $I^2$ = 71.0 , HetP = 1.52e-04 ; Global : $I^2$ = 69.5 , HetP = 1.64e-04

| Country | N | Beta | 95% CI | P-value |
|---|---|---|---|---|
| Finland | 226 | -0.180 | [-0.31;-0.05] | 8.40e-03 |
| Sweden | 264 | -0.138 | [-0.26;-0.02] | 2.88e-02 |
| The Netherlands | 3,106 | -0.148 | [-0.18;-0.12] | 4.27e-18 |
| Germany 1 | 314 | -0.102 | [-0.21;0.00] | 5.66e-02 |
| Germany 2 | 75 | 0.028 | [-0.22;0.28] | 8.29e-01 |
| Germany 3 | 145 | -0.140 | [-0.32;0.04] | 1.21e-01 |
| Germany 4 | 112 | 0.109 | [-0.08;0.30] | 2.56e-01 |
| Belgium | 598 | 0.031 | [-0.05;0.11] | 4.46e-01 |
| France 1 | 439 | -0.068 | [-0.16;0.03] | 1.65e-01 |
| France 2 | 135 | 0.081 | [-0.09;0.26] | 3.68e-01 |
| Spain | 408 | -0.216 | [-0.31;-0.12] | 2.10e-05 |
| **Meta-analysis Europe (random effect)** | **5,822** | **-0.084** | **[-0.14;-0.02]** | **6.07e-03** |
| European American | 7,229 | -0.091 | [-0.12;-0.06] | 7.22e-09 |
| **Meta-analysis (random effect)** | **13,051** | **-0.088** | **[-0.13;-0.04]** | **1.83e-04** |
| **Adjusted for status** | | | | **4.49e-03** |

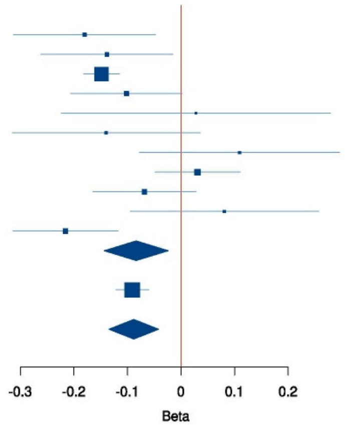


**B**

pTau ($PGS^{ALZ}$)

Europe : $I^2$ = 0 , HetP = 8.37e-01 ; Global : $I^2$ = 0 , HetP = 8.65e-01

| Country | N | Beta | 95% CI | P-value |
|---|---|---|---|---|
| Finland | 226 | 0.097 | [-0.08;0.28] | 2.92e-01 |
| Sweden | 264 | 0.143 | [0.02;0.26] | 1.89e-02 |
| The Netherlands | 3,106 | 0.119 | [0.09;0.15] | 3.16e-12 |
| Germany 1 | 314 | 0.051 | [-0.06;0.16] | 3.45e-01 |
| Germany 2 | 75 | 0.020 | [-0.25;0.29] | 8.86e-01 |
| Germany 3 | 145 | 0.034 | [-0.14;0.21] | 7.08e-01 |
| Germany 4 | 112 | -0.039 | [-0.23;0.16] | 6.98e-01 |
| Belgium | 598 | 0.085 | [0.00;0.16] | 3.84e-02 |
| France 1 | 439 | 0.105 | [0.01;0.20] | 3.18e-02 |
| France 2 | 135 | 0.052 | [-0.14;0.24] | 5.94e-01 |
| Spain | 408 | 0.120 | [0.02;0.22] | 2.27e-02 |
| **Meta-analysis Europe** | **5,822** | **0.105** | **[0.08;0.13]** | **5.46e-16** |
| European American | 7,229 | 0.123 | [0.07;0.18] | 4.53e-06 |
| **Meta-analysis** | **13,051** | **0.108** | **[0.09;0.13]** | **1.55e-20** |
| **Adjusted for status** | | | | **1.61e-07** |

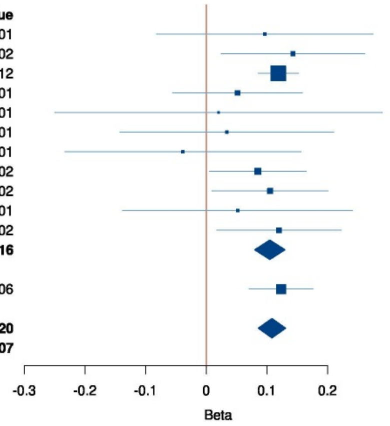


**C**

Tau ($PGS^{ALZ}$)

Europe : $I^2$ = 0 , HetP = 6.40e-01 ; Global : $I^2$ = 0 , HetP = 6.91e-01

| Country | N | Beta | 95% CI | P-value |
|---|---|---|---|---|
| Finland | 226 | 0.089 | [-0.08;0.26] | 3.05e-01 |
| Sweden | 264 | 0.150 | [0.03;0.27] | 1.17e-02 |
| The Netherlands | 3,106 | 0.135 | [0.10;0.17] | 1.90e-15 |
| Germany 1 | 314 | 0.092 | [-0.01;0.20] | 7.95e-02 |
| Germany 2 | 75 | 0.090 | [-0.15;0.33] | 4.68e-01 |
| Germany 3 | 145 | 0.078 | [-0.10;0.26] | 3.97e-01 |
| Germany 4 | 112 | -0.100 | [-0.28;0.08] | 2.82e-01 |
| Belgium | 598 | 0.113 | [0.03;0.19] | 5.51e-03 |
| France 1 | 439 | 0.111 | [0.01;0.21] | 2.54e-02 |
| France 2 | 135 | 0.117 | [-0.06;0.30] | 2.07e-01 |
| Spain | 408 | 0.157 | [0.06;0.26] | 2.16e-03 |
| **Meta-analysis Europe** | **5,822** | **0.123** | **[0.10;0.15]** | **5.45e-22** |
| European American | 7,229 | 0.109 | [0.07;0.15] | 3.56e-08 |
| **Meta-analysis** | **13,051** | **0.119** | **[0.10;0.14]** | **1.39e-28** |
| **Adjusted for status** | | | | **1.06e-10** |

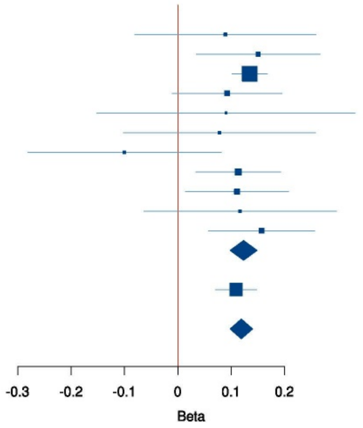

**Supplementary Figure 8:** GRS distribution (European American MVP, Asutralia, Maghreb, Sub Saharan Africa, African American MVP, African American ADSP)

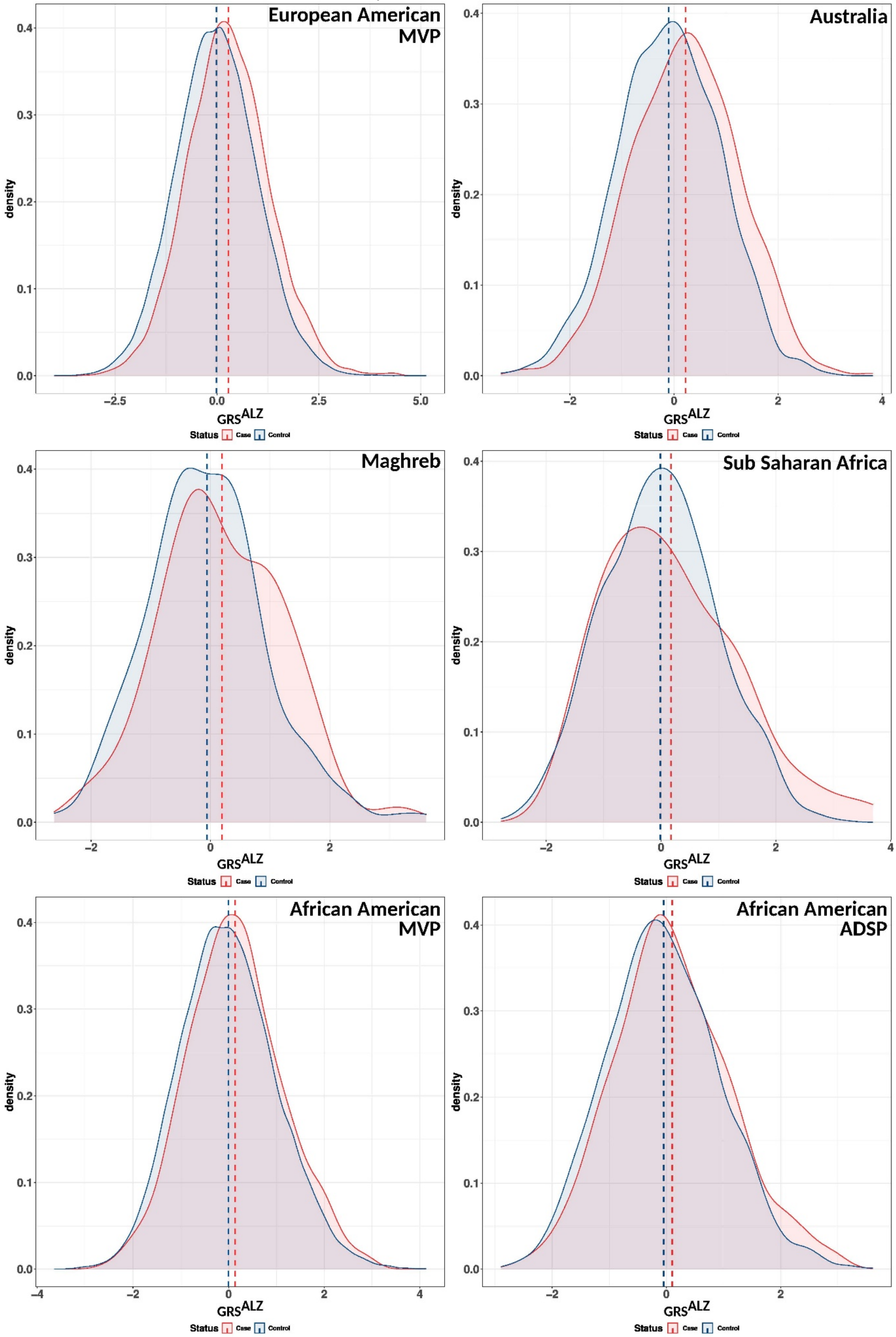

**Supplementary Figure 9:** GRS distribution (US LA ancestry MVP, US LA ancestry ADSP, Colombia,, Brazil, Argentina, Chile)

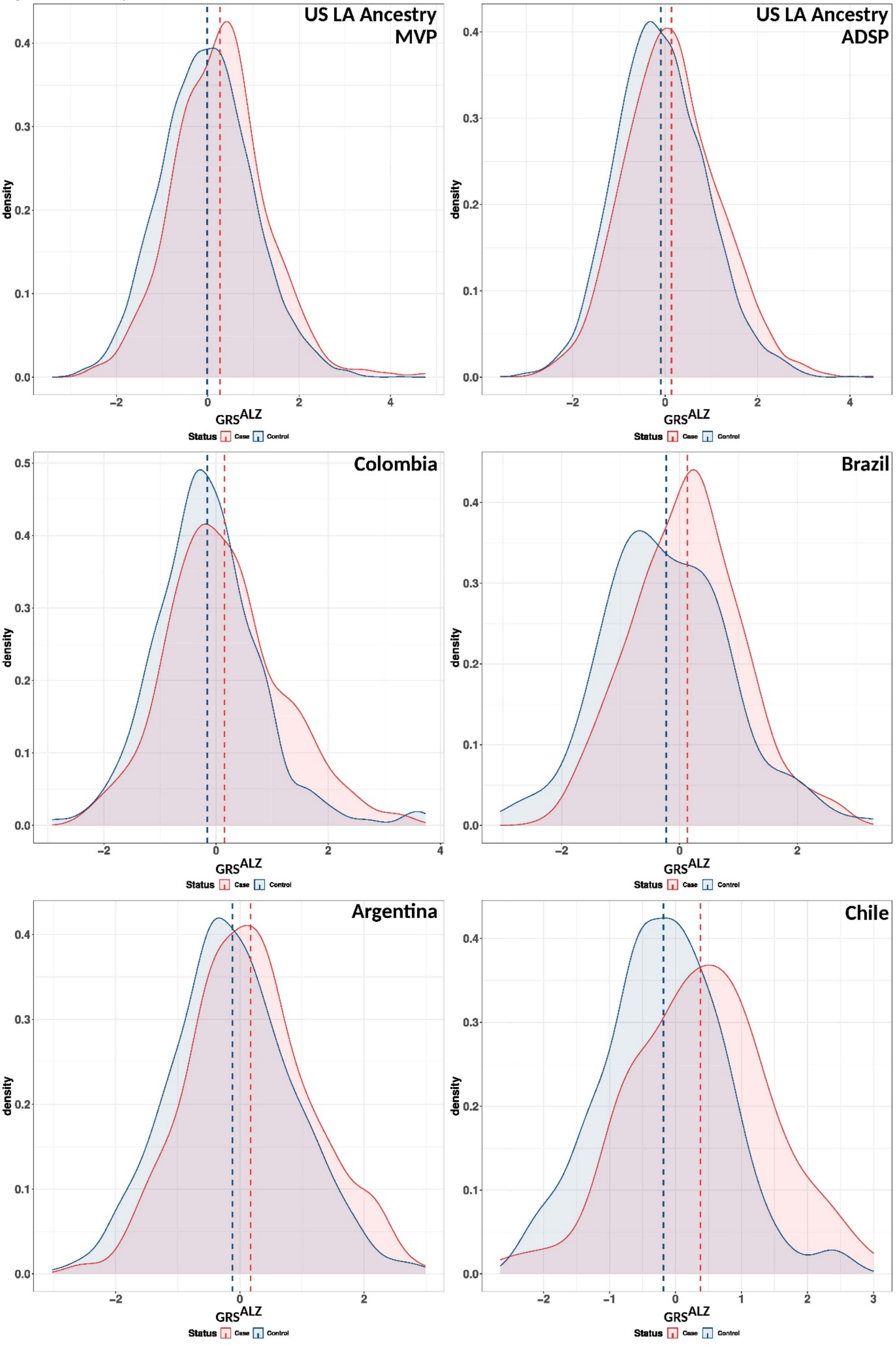

**Supplementary Figure 10:** GRS distribution (China, Japan, South Korea, India)

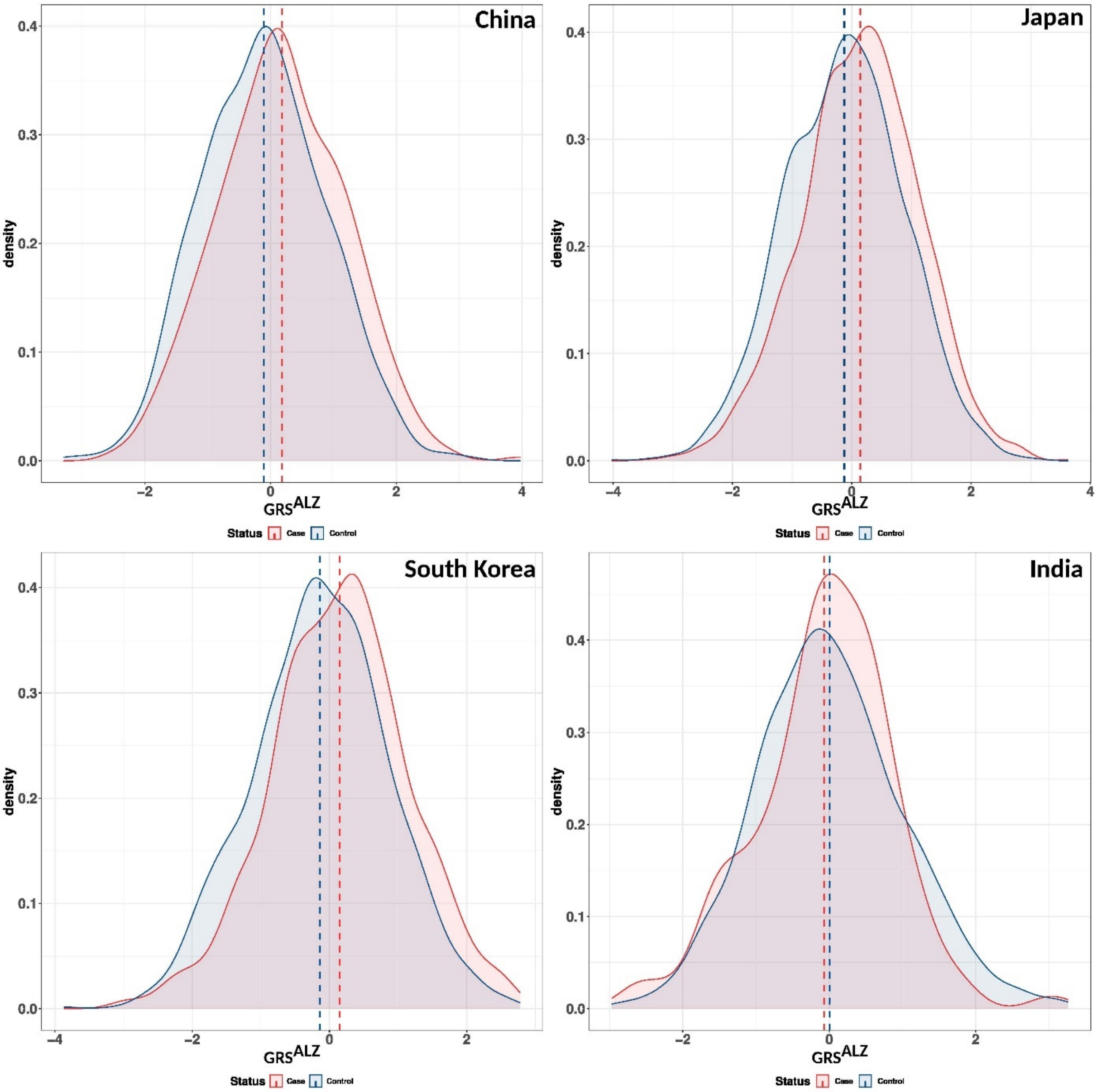

**Supplementary Figure 11:** Association of $GRS^{ALZ}$ with the risk of developing AD in multi-ancestry populations. European ancestry meta-analysis includes MVP and Australia. African-American-ancestry (more than 75% AA ancestry) meta-analysis includes MVP and ADSP. East-Asia meta-analysis includes China, Korea and Japan. Latino-American ancestry (self-reporting) meta-analysis includes MVP, ADSP and Salsa. South-America meta-analysis includes Argentina, Brazil, Chile and Colombia. Ncase, number of cases; Ncontrols, number of controls, OR, Odds ratio. The lines in the Forrest plots indicate the 95% confidence interval for the ORs. If HetP <0.05, random-effect is shown for the meta-analysis results.

$GRS^{ALZ}$

| Country | Ncases | Ncontrols | OR | 95% CI | P−value | HetIsq | HetPVal |
|---|---|---|---|---|---|---|---|
| European American MVP | 4,561 | 84,587 | 1.39 | [1.34−1.44] | 2.40e−81 | | |
| Australia | 544 | 1,190 | 1.39 | [1.25−1.54] | 1.04e−09 | | |
| **Meta−analysis EUR Ancestry** | **5,105** | **85,777** | **1.39** | **[1.34−1.43]** | **1.87e−89** | **0** | **9.99e−01** |
| Maghreb | 58 | 187 | 1.46 | [1.01−2.15] | 4.81e−02 | | |
| Sub−Saharan Africa | 85 | 1,091 | 1.11 | [0.89−1.39] | 3.48e−01 | | |
| African American MVP | 713 | 19,405 | 1.20 | [1.10−1.30] | 5.12e−05 | | |
| African American ADSP | 1,108 | 2,325 | 1.15 | [1.07−1.24] | 1.72e−04 | | |
| **Meta−analysis AA Ancestry** | **1,821** | **21,730** | **1.17** | **[1.11−1.24]** | **4.04e−08** | **0** | **5.31e−01** |
| US LA Ancestry MVP | 375 | 7,166 | 1.30 | [1.15−1.46] | 3.38e−05 | | |
| US LA Ancestry ADSP | 1,629 | 2,456 | 1.26 | [1.18−1.35] | 3.58e−12 | | |
| Colombia | 254 | 241 | 1.27 | [1.04−1.57] | 2.14e−02 | | |
| Brazil | 301 | 178 | 1.57 | [1.28−1.95] | 2.39e−05 | | |
| Argentina | 417 | 601 | 1.32 | [1.14−1.53] | 1.51e−04 | | |
| Chile | 122 | 252 | 1.77 | [1.39−2.28] | 5.60e−06 | | |
| **Meta−analysis US LA Ancestry** | **2,004** | **9,622** | **1.27** | **[1.20−1.35]** | **6.12e−16** | **0** | **7.10e−01** |
| **Meta−analysis South America** | **1,094** | **1,272** | **1.42** | **[1.29−1.56]** | **5.44e−13** | **50.7** | **1.08e−01** |
| **Meta−analysis LA Ancestry** | **3,098** | **10,894** | **1.31** | **[1.25−1.37]** | **1.57e−26** | **50.1** | **7.45e−02** |
| China | 453 | 786 | 1.35 | [1.19−1.54] | 2.33e−06 | | |
| Japan | 2,220 | 2,472 | 1.30 | [1.22−1.38] | 1.76e−17 | | |
| South Korea | 1,119 | 1,172 | 1.35 | [1.24−1.47] | 5.57e−12 | | |
| **Meta−analysis East Asia** | **3,792** | **4,430** | **1.32** | **[1.26−1.38]** | **1.32e−32** | **0** | **6.75e−01** |
| India | 105 | 1,500 | 0.97 | [0.79−1.19] | 7.85e−01 | | |

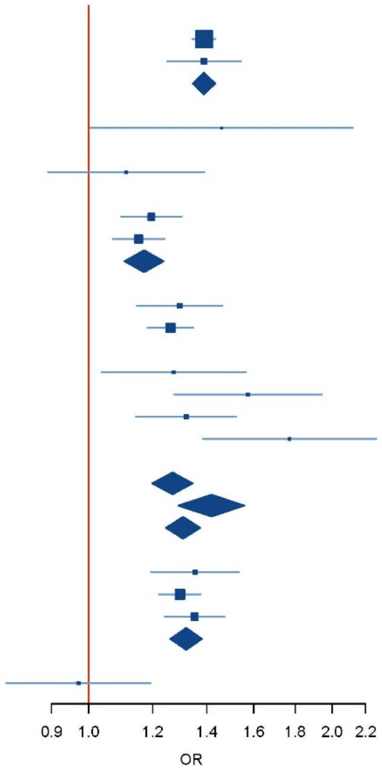

**Supplementary Figure 12.** Association of GRS$^{APOE}$ with age at onset in multi-ancestry populations. European ancestry meta-analysis includes MVP and Australia. African-American-ancestry (more than 75% AA ancestry) meta-analysis includes MVP and ADSP. East-Asia meta-analysis includes China, Korea and Japan. Latino-American ancestry (self-reporting) meta-analysis includes MVP, ADSP and Salsa. South-America meta-analysis includes Argentina, Brazil, Chile and Colombia. Ncase, The lines in the Forrest plots indicate the 95% confidence interval for the ORs. If HetP <0.05, random-effect is shown for the meta-analysis results.

GRS$^{APOE}$

| Country | Ncases | Beta | 95% CI | P−value | I² | HetP |
|---|---|---|---|---|---|---|
| Finland | 1,118 | −1.53 | [−1.98;−1.07] | 6.03e−11 | | |
| Norway | 1,327 | −1.25 | [−1.72;−0.78] | 2.54e−07 | | |
| Sweden | 1,466 | −3.18 | [−3.67;−2.68] | 2.31e−34 | | |
| Denmark | 398 | −1.46 | [−2.15;−0.77] | 4.06e−05 | | |
| United Kingdom | 4,622 | −1.06 | [−1.31;−0.80] | 1.11e−15 | | |
| The Netherlands | 2,428 | −0.68 | [−1.10;−0.27] | 1.20e−03 | | |
| Belgium | 1,211 | −1.13 | [−1.42;−0.84] | 2.29e−14 | | |
| Germany | 2,190 | −1.83 | [−2.17;−1.49] | 6.54e−25 | | |
| Austria/Switzerland | 175 | −0.42 | [−1.20;0.36] | 2.88e−01 | | |
| Bulgaria/Greece | 735 | 0.01 | [−0.53;0.55] | 9.77e−01 | | |
| Italy | 2,973 | −0.80 | [−1.10;−0.49] | 3.00e−07 | | |
| France | 3,789 | −1.32 | [−1.63;−1.02] | 1.15e−17 | | |
| Spain | 3,103 | −1.12 | [−1.41;−0.82] | 1.54e−13 | | |
| **Meta−Analysis Europe (random effect)** | **25,535** | **−1.22** | **[−1.55;−0.89]** | **2.77e−13** | **89.5** | **6.63e−18** |
| European American MVP | 4,561 | −0.89 | [−1.08;−0.69] | 4.80e−19 | | |
| Australia | 544 | −0.52 | [−1.22;0.17] | 1.41e−01 | | |
| **Meta−analysis EUR Ancestry** | **5,105** | **−0.86** | **[−1.05;−0.67]** | **1.70e−19** | **0** | **3.22e−01** |
| African American MVP | 713 | −1.01 | [−1.57;−0.45] | 4.27e−04 | | |
| African American ADSP | 1,108 | −2.33 | [−2.81;−1.85] | 1.73e−20 | | |
| **Meta−analysis AA Ancestry (random effect)** | **1,821** | **−1.68** | **[−2.97;−0.38]** | **1.11e−02** | **91.9** | **4.51e−04** |
| US LA Ancestry MVP | 375 | −0.50 | [−1.22;0.22] | 1.75e−01 | | |
| US LA Ancestry ADSP | 1,629 | −1.77 | [−2.15;−1.39] | 2.81e−19 | | |
| Colombia | 254 | 0.15 | [−0.86;1.17] | 7.65e−01 | | |
| Brazil | 301 | −1.27 | [−2.02;−0.52] | 9.38e−04 | | |
| Argentina | 417 | −0.76 | [−1.32;−0.20] | 7.92e−03 | | |
| Chile | 122 | −1.61 | [−3.36;0.14] | 7.13e−02 | | |
| **Meta−analysis US LA Ancestry (random effect)** | **2,004** | **−1.17** | **[−2.41;0.07]** | **6.49e−02** | **89.4** | **2.15e−03** |
| **Meta−analysis South America** | **1,094** | **−0.81** | **[−1.21;−0.41]** | **6.74e−05** | **48.4** | **1.21e−01** |
| **Meta−analysis LA Ancestry (random effect)** | **3,098** | **−0.96** | **[−1.57;−0.35]** | **1.98e−03** | **77.0** | **5.92e−04** |
| China | 453 | −0.87 | [−1.37;−0.37] | 6.68e−04 | | |
| Japan | 2,220 | −0.84 | [−1.05;−0.63] | 9.19e−15 | | |
| South Korea | 1,119 | −1.23 | [−1.59;−0.87] | 2.78e−11 | | |
| **Meta−analysis East Asia** | **3,792** | **−0.93** | **[−1.10;−0.76]** | **1.09e−26** | **43.2** | **1.72e−01** |
| India | 105 | 0.37 | [−1.10;1.85] | 6.15e−01 | | |

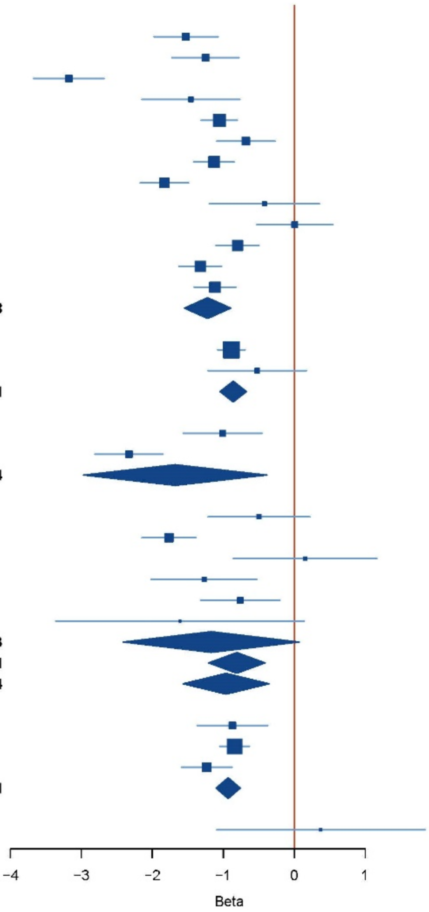

**Supplementary Figure 13:** Workflow indicating the summary statistics and populations used todevelop and test GRS, GRS+ and PRS.

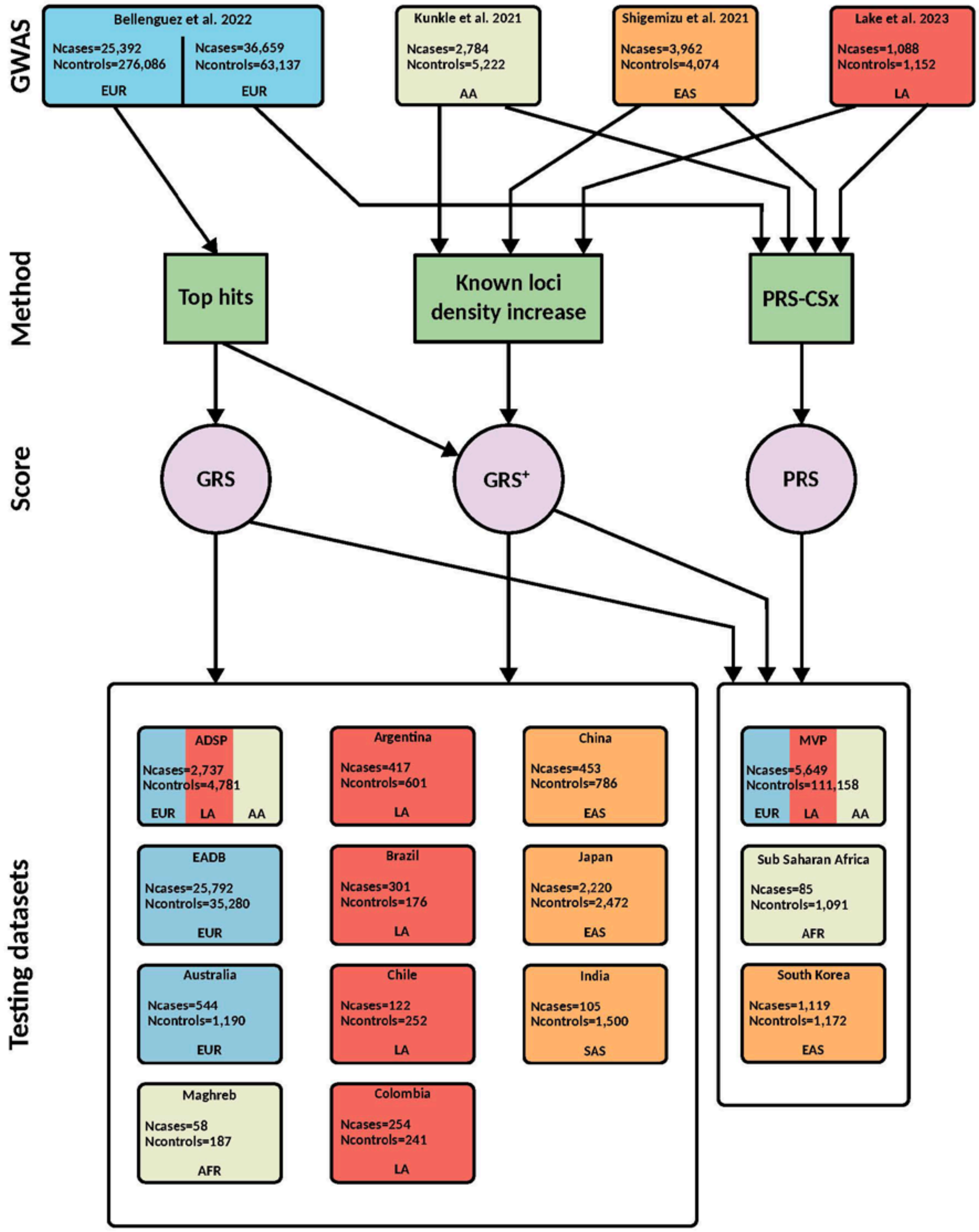

#### REFERENCES


1. De Roeck, A. et al. An intronic VNTR affects splicing of ABCA7 and increases risk of Alzheimer's disease. Acta Neuropathol. 135, 827–837 (2018).
2. Sheardova, K. et al. Czech Brain Aging Study (CBAS): Prospective multicentre cohort study on risk and protective factors for dementia in the Czech Republic. BMJ Open 9, (2019).
3. Steinberg, S. et al. Loss-of-function variants in ABCA7 confer risk of Alzheimer's disease. Nat. Genet. 47, 445–447 (2015).
4. Ngandu, T. et al. A 2 year multidomain intervention of diet, exercise, cognitive training, and vascular risk monitoring versus control to prevent cognitive decline in at-risk elderly people (FINGER): A randomised controlled trial. Lancet 385, 2255–2263 (2015).
5. Hanon, O. et al. Plasma amyloid levels within the Alzheimer's process and correlations with central biomarkers. Alzheimer's Dement. 14, 858–868 (2018).
6. Dufouil, C. et al. Cognitive and imaging markers in non-demented subjects attending a memory clinic: Study design and baseline findings of the MEMENTO cohort. Alzheimer's Res. Ther. 9, (2017).
7. Nicolas, G. et al. Screening of dementia genes by whole-exome sequencing in early-onset Alzheimer disease: Input and lessons. Eur. J. Hum. Genet. 24, 710–716 (2016).
8. McKhann, G. et al. Clinical diagnosis of Alzheimer's disease: report of the NINCDS-ADRDA Work Group under the auspices of Department of Health and Human Services Task Force on Alzheimer's Disease. Neurology 34, 939–44 (1984).
9. Kornhuber, J. et al. Early and differential diagnosis of dementia and mild cognitive impairment: Dement. Geriatr. Cogn. Disord. 27, 404–417 (2009).
10. Luck, T. et al. Mild cognitive impairment in general practice: Age-specific prevalence and correlate results from the German study on ageing, cognition and dementia in primary care patients (AgeCoDe). Dement. Geriatr. Cogn. Disord. 24, 307–316 (2007).
11. McKhann, G. M. et al. The diagnosis of dementia due to Alzheimer's disease: Recommendations from the National Institute on Aging-Alzheimer's Association workgroups on diagnostic guidelines for Alzheimer's disease. Alzheimer's Dement. 7, 263–269 (2011).
12. Van Der Flier, W. M. & Scheltens, P. Amsterdam dementia cohort: Performing research to optimize care. Journal of Alzheimer's Disease vol. 62 1091–1111 (2018).
13. Holstege, H. et al. The 100-plus Study of cognitively healthy centenarians: rationale, design and cohort description. Eur. J. Epidemiol. 33, (2018).
14. Aalten, P. et al. The Dutch Parelsnoer Institute - Neurodegenerative diseases; methods, design and baseline results. BMC Neurol. 14, (2014).
15. Dubois, B. et al. Research criteria for the diagnosis of Alzheimer's disease: revising the NINCDS-ADRDA criteria. Lancet Neurology vol. 6 734–746 (2007).
16. Ramakers, I. et al. Biobank Alzheimer Center Limburg cohort: design and cohort characteristics. Prep.
17. (APA), A. P. A. Diagnostic and statistical manual of mental disorders. (1994).
18. Sachdev, P. S. et al. The Sydney Memory and Ageing Study (MAS): Methodology and baseline medical and neuropsychiatric characteristics of an elderly epidemiological non-demented cohort of Australians aged 70-90 years. Int. Psychogeriatrics 22, 1248–1264 (2010).
19. Moreno-Grau, S. et al. Genome-wide association analysis of dementia and its clinical endophenotypes reveal novel loci associated with Alzheimer's disease and three causality networks: The GR@ACE project. Alzheimer's Dement. 15, 1333–1347 (2019).
20. Ruiz, A. et al. Assessing the role of the TREM2 p.R47H variant as a risk factor for Alzheimer's disease and frontotemporal dementia. Neurobiol. Aging 35, 444.e1–4 (2014).
21. Ikram, M. A. et al. The Rotterdam Study: 2018 update on objectives, design and main results. Eur. J. Epidemiol. 32, 807–850 (2017).
22. Niemeijer, M. N. et al. ABCB1 gene variants, digoxin and risk of sudden cardiac death in a general population. Heart 101, 1973–1979 (2015).
23. Loh, P.-R. et al. Reference-based phasing using the Haplotype Reference Consortium panel. Nat. Genet. 48, 1443–1448 (2016).
24. De Bruijn, R. F. A. G. et al. Determinants, MRI correlates, and prognosis of mild cognitive impairment: The rotterdam study. in Journal of Alzheimer's Disease vol. 42 S239–S249 (IOS Press, 2014).
25. 3C Study Group. Vascular factors and risk of dementia: design of the Three-City Study and baseline characteristics of the study population. Neuroepidemiology 22, 316–25 (2003).

26. Lambert, J.-C. et al. Genome-wide association study identifies variants at CLU and CR1 associated with Alzheimer's disease. Nat. Genet. 41, 1094–9 (2009).
27. Harold, D. et al. Genome-wide association study identifies variants at CLU and PICALM associated with Alzheimer's disease. Nat. Genet. 41, 1088–93 (2009).
28. Jansen, I. E. et al. Genome-wide meta-analysis identifies new loci and functional pathways influencing Alzheimer's disease risk. Nat. Genet. 51, 404–413 (2019).
29. Gayán, J. et al. Genetic Structure of the Spanish Population. BMC Genomics 11, (2010).
30. Antúnez, C. et al. The membrane-spanning 4-domains, subfamily A (MS4A) gene cluster contains a common variant associated with Alzheimer's disease. Genome Med. 3, (2011).
31. Jessen, F. et al. AD dementia risk in late MCI, in early MCI, and in subjective memory impairment. Alzheimer's Dement. 10, 76–83 (2014).
32. Reisberg, B., Ferris, S. H., De Leon, M. J. & Crook, T. The global deterioration scale for assessment of primary degenerative dementia. Am. J. Psychiatry 139, 1136–1139 (1982).
33. Lambert, J. C. et al. Meta-analysis of 74,046 individuals identifies 11 new susceptibility loci for Alzheimer's disease. Nat. Genet. 45, 1452–8 (2013).
34. Schmermund, A. et al. Assessment of clinically silent atherosclerotic disease and established and novel risk factors for predicting myocardial infarction and cardiac death in healthy middle-aged subjects: Rationale and design of the Heinz Nixdorf RECALL study. Am. Heart J. 144, 212–218 (2002).
35. Stang, A. et al. Baseline recruitment and analyses of nonresponse of the Heinz Nixdorf Recall Study: Identifiability of phone numbers as the major determinant of response. Eur. J. Epidemiol. 20, 489–496 (2005).
36. Winkler, A. *et al.* Association of diabetes mellitus and mild cognitive impairment in middle-aged men and women. *J. Alzheimer's Dis.* **42**, 1269–1277 (2014).
37. Guerchet, M. *et al*. Epidemiology of dementia in Central Africa (EPIDEMCA): protocol for a multicentre population-based study in rural and urban areas of the Central African Republic and the Republic of Congo. *Springerplus* **3**, 338 (2014).
38. Ben Salem, N. *et al*. Mitochondrial DNA and Alzheimer's disease: a first case-control study of the Tunisian population. *Mol Biol Rep*. **49**, 1687-1700 (2022).
39. Dalmasso, M.C. *et al*. Transethnic meta-analysis of rare coding variants in PLCG2, ABI3, and TREM2 supports their general contribution to Alzheimer's disease. *Transl. Psychiatry* **9**, 55 (2019).
40. Albala, C. et al. Low Leptin Availability as a Risk Factor for Dementia in Chilean Older People. *Dement. Geriatr. Cogn. Dis. Extra.* **6**, 295-302 (2016).
41. Quiroga, P. *et al*. Validación de un test de tamizaje para el diagnóstico de demencia asociada a edad, en Chile [Validation of a screening test for age associated cognitive impairment, in Chile]. *Rev. Med. Chil*. **132**, 467-78 (2004).
42. Slachevsky, A. *et al*. GERO Cohort Protocol, Chile, 2017-2022: Community-based Cohort of Functional Decline in Subjective Cognitive Complaint elderly. *BMC Geriatr*. **20**, 505 (2020).
43. de Paula, J.J. *et al*. The Stick Design Test on the assessment of older adults with low formal education: evidences of construct, criterion-related and ecological validity. *Int. Psychogeriatr.* **25**, 2057-65 (2013).
44. Beecham, G.W. *et al.* The Alzheimer's Disease Sequencing Project: Study design and sample selection. *Neurol. Genet*. **3**, e194 (2017).
45. Leung, Y.Y. *et al.* VCPA: genomic variant calling pipeline and data management tool for Alzheimer's Disease Sequencing Project. *Bioinformatics* **35**, 1768-1770 (2019).
46. Chen, C.Y. *et al*. Improved ancestry inference using weights from external reference panels. *Bioinformatics* **2,** 1399-406 (2013).
47. Le Guen, Y. et al. Multiancestry analysis of the HLA locus in Alzheimer's and Parkinson's diseases uncovers a shared adaptive immune response mediated by HLA-DRB1*04 subtypes. *Proc. Natl. Acad. Sci.* 120, e2302720120 (2023).
48. Danecek, P. *et al.* 1000 Genomes Project Analysis Group. The variant call format and VCFtools. *Bioinformatics* **27**, 2156-8 (2011).
49. Manichaikul, A. *et al.* Robust relationship inference in genome-wide association studies. *Bioinformatics* **26**, 2867-73 (2010).
50. Patterson, N. et al. Population structure and eigenanalysis. *PLoS Genet*. **2**, e190 (2006).

51. Martinez-Miller, E.E. *et al*. Longitudinal Associations of US Acculturation With Cognitive Performance, Cognitive Impairment, and Dementia. *Am. J. Epidemiol.* **189**, 1292-1305 (2020).
52. Miyashita, A. *et al*. SORL1 is genetically associated with late-onset Alzheimer's disease in Japanese, Koreans and Caucasians. *PLoS One* **8**, e58618 (2013).
53. Iwatsubo, T. *et al*. Japanese and North American Alzheimer's Disease Neuroimaging Initiative studies: Harmonization for international trials. *Alzheimers Dement*. **14**, 1077-1087 (2018).
54. Jung, S.H. *et al*. Transferability of Alzheimer Disease Polygenic Risk Score Across Populations and Its Association With Alzheimer Disease-Related Phenotypes. *JAMA Netw. Open* **5**, e2247162 (2022).
55. Zhou, X. *et al*. Genetic and polygenic risk score analysis for Alzheimer's disease in the Chinese population. *Alzheimers Dement*. **12**, e12074 (2020).
56. Chen, S., Zhou, Y., Chen, Y. & Gu, J. fastp: an ultra-fast all-in-one FASTQ preprocessor. Bioinformatics 34, i884–i890 (2018).
57. Li, H. Aligning sequence reads, clone sequences and assembly contigs with BWA-MEM. (2013).
58. DePristo, M. A. et al. A framework for variation discovery and genotyping using next-generation DNA sequencing data. Nature Genetics 2011 43:5 43, 491–498 (2011).
59. Browning, S. R. & Browning, B. L. Rapid and Accurate Haplotype Phasing and Missing-Data Inference for Whole-Genome Association Studies By Use of Localized Haplotype Clustering. Am J Hum Genet 81, 1084 (2007).
60. Chang, C. C. et al. Second-generation PLINK: rising to the challenge of larger and richer datasets. Gigascience 4, 7 (2015).
61. Privé, F., Luu, K., Blum, M. G. B., McGrath, J. J. & Vilhjálmsson, B. J. Efficient toolkit implementing best practices for principal component analysis of population genetic data. Bioinformatics 36, 4449–4457 (2020).
62. Gaziano, J.M. *et al*. Million Veteran Program: A mega-biobank to study genetic influences on health and disease. *J. Clin. Epidemiol*. **70**, 214-23 (2016).
63. Nguyen, X.T. *et al*. VA Million Veteran Program. Data Resource Profile: Self-reported data in the Million Veteran Program: survey development and insights from the first 850 736 participants. *Int. J. Epidemiol*. **52**, e1-e17 (2023).
64. Hunter-Zinck, H. *et al.* Genotyping Array Design and Data Quality Control in the Million Veteran Program. *Am. J. Hum. Genet*. **106**, 535-548 (2020).
65. Fang H*, et al*. Harmonizing Genetic Ancestry and Self-identified Race/Ethnicity in Genome-wide Association Studies. Am J Hum Genet. 2019 Oct 3;105(4):763-772.
66. Sherva, R. *et al*. African ancestry GWAS of dementia in a large military cohort identifies significant risk loci. *Mol. Psychiatry*. **28**, 1293-1302 (2023).
67. Cho, K. *et al.* Dementia Coding, Workup, and Treatment in the VA New England Healthcare System. *Int. J. Alzheimers Dis*. **2014**, 821894 (2014).
68. Timsina J, et al. Harmonization of CSF and imaging biomarkers in Alzheimer's disease: Need and practical applications for genetics studies and preclinical classification. *Neurobiol Dis*. **190**,106373 (2024).
69. Ali M, et al. Large multi-ethnic genetic analyses of amyloid imaging identify new genes for Alzheimer disease. *Acta Neuropathol Commun*. **11**, 68 (2023).

#### 2. ACKNOWLEDGEMENTS

**Additional support for EADB cohorts**

The work for this manuscript was further supported by the CoSTREAM project (www.costream.eu) and funding from the European Union's Horizon 2020 research and innovation programme under grant agreement No 667375. This work is also funded by la fondation pour la recherché médicale (FRM) (EQU202003010147) Italian Ministry of Health (Ricerca Corrente); Ministero dell'Istruzione, del l'Università e della Ricerca–MIUR project "Dipartimenti di Eccellenza 2018–2022" to Department of Neuroscience "Rita Levi Montalcini", University of Torino (IR), and AIRAlzh Onlus-ANCC-COOP (SB); Partly supported by "Ministero della Salute", I.R.C.C.S. Research Program, Ricerca Corrente 2018-2020, Linea n. 2 "Meccanismi genetici, predizione e terapie innovative delle malattie complesse" and by the "5 x 1000" voluntary contribution to the Fondazione I.R.C.C.S. Ospedale "Casa Sollievo della Sofferenza"; and RF-2018-12366665, Fondi per la ricerca 2019 (Sandro Sorbi) and Italian Ministry of Health, Ricerca Corrente (RG, IRCCS Istituto Centro San Giovanni di Dio Fatebenefratelli Brescia); Copenhagen General Population Study (CGPS): We thank staff and participants of the CGPS for their important contributions. Karolinska Institutet AD cohort: Dr. C.G. and co-authors of the Karolinska Institutet AD cohort report grants from Swedish Research Council (VR) 2015-02926, 2018-02754, 2015-06799, Swedish Alzheimer Foundation, Stockholm County Council ALF and resarch school, Karolinska Institutet StratNeuro, Swedish Demensfonden, and Swedish brain foundation, during the conduct of the study. ADGEN: This work was supported by Academy of Finland (grant numbers 307866); Sigrid Jusélius Foundation; the Strategic Neuroscience Funding of the University of Eastern Finland; EADB project in the JPNDCO-FUND program (grant number 301220). CBAS: Supported by the Ministry of Health, Czech Republic—conceptual development of research organization, University Hospital Motol, Prague, Czech Republic Grant No. 00064203; Institutional Support of Excellence 2. LF UK Grant No. 699012; and The project National Institute for Neurological Research (Programme EXCELES, ID Project No. LX22NPO5107) - Funded by the European Union – Next Generation EU. CNRMAJ-Rouen: This study received fundings from the Centre National de Référence Malades Alzheimer Jeunes (CNRMAJ). The Finnish Geriatric Intervention Study for the Prevention of Cognitive Impairment and Disability (FINGER) data collection was supported by grants from the Academy of Finland, La Carita Foundation, Juho Vainio Foundation, Novo Nordisk Foundation, Finnish Social Insurance Institution, Ministry of Education and Culture Research Grants, Yrjö Jahnsson Foundation, Finnish Cultural Foundation South Osthrobothnia Regional Fund, and EVO/State Research Funding grants of University Hospitals of Kuopio, Oulu and Turku, Seinäjoki Central Hospital and Oulu City Hospital, Alzheimer's Research & Prevention Foundation USA, AXA Research Fund, Knut and Alice Wallenberg Foundation Sweden, Center for Innovative Medicine (CIMED) at Karolinska Institutet Sweden, and Stiftelsen Stockholms sjukhem Sweden. FINGER cohort genotyping was funded by EADB project in the JPND CO-FUND (grant number 301220). Research at the Belgian EADB site is funded in part by the Alzheimer Research Foundation (SAO-FRA), The Research Foundation Flanders (FWO), and the University of Antwerp Research Fund. FK is supported by a fellowship of the University of Antwerp Research Fund. SNAC-K is financially supported by the Swedish Ministry of Health and Social Affairs, the participating County Councils and Municipalities, and the Swedish Research Council. BDR Bristol: We would like to thank the South West Dementia Brain Bank (SWDBB) for providing brain tissue for this study. The SWDBB is part of the Brains for Dementia Research programme, jointly funded by Alzheimer's Research UK and Alzheimer's Society and is supported by BRACE (Bristol Research into Alzheimer's and Care of the Elderly) and the Medical Research Council. BDR Manchester: We would like to thank the Manchester Brain Bankfor providing brain tissue for this study. The Manchester Brain Bank is part of the Brains for Dementia Research programme, jointly funded by Alzheimer’s Research UK and Alzheimer’s Society. BDR KCL: Human post-mortem tissue was provided by the London Neurodegenerative Diseases Brain Bank which receives funding from the UK Medical Research Council and as part of the Brains for Dementia Research programme, jointly funded by Alzheimer’s Research UK and the Alzheimer’s Society. The CFAS Wales study was funded by the ESRC (RES-060-25-0060) and HEFCW as ‘Maintaining function and well-being in later life: a longitudinal cohort study’. We are grateful to the NISCHR Clinical Research Centre for their assistance in tracing participants and in interviewing and in collecting blood samples, and to general practices in the study areas for their cooperation. MRC: We thank all individuals who participated in this study. Cardiff University was supported by the Alzheimer's Society (AS; grant RF014/164) and the Medical Research Council (MRC; grants G0801418/1, MR/K013041/1, MR/L023784/1) (R.S. is an AS Research Fellow). Cardiff University was also supported by the European Joint Programme for Neurodegenerative Disease (JPND; grant MR/L501517/1), Alzheimer's Research UK (ARUK; grant ARUK-PG2014-1), the Welsh Assembly Government (grant SGR544:CADR), Brain’s for dementia Research and a donation from the Moondance Charitable Foundation. Cardiff University acknowledges the support of the UK Dementia Research Institute, of which J.W. is an associate director. Cambridge University acknowledges support from the MRC. Patient recruitment for the MRC Prion Unit/UCL Department of Neurodegenerative Disease collection was supported by the UCLH/UCL Biomedical Centre and NIHR Queen Square Dementia Biomedical Research Unit. The University of Southampton acknowledges support from the AS. King's College London was supported by the NIHR Biomedical Research Centre for Mental Health and the Biomedical Research Unit for Dementia at the South London and Maudsley NHS Foundation Trust and by King's College London and the MRC. ARUK and the Big Lottery Fund provided support to Nottingham University. A.Ram. : Part of the work was funded by the JPND EADB grant (German Federal Ministry of Education and Research (BMBF) grant: 01ED1619A). A. Ram. is also supported by the German Research Foundation (DFG) grants Nr: RA 1971/6-1, RA1971/7-1, and RA 1971/8-1. German Study on Ageing, Cognition and Dementia in Primary Care Patients (AgeCoDe): This study/publication is part of the German Research Network on Dementia (KND), the German Research Network on Degenerative Dementia (KNDD; German Study on Ageing, Cognition and Dementia in Primary Care Patients; AgeCoDe), and the Health Service Research Initiative (Study on Needs, health service use, costs and health-related quality of life in a large sample of oldestold primary care patients (85+; AgeQualiDe)) and was funded by the German Federal Ministry of Education and Research (grants KND: 01GI0102, 01GI0420, 01GI0422, 01GI0423, 01GI0429, 01GI0431, 01GI0433, 01GI0434; grants KNDD: 01GI0710, 01GI0711, 01GI0712, 01GI0713, 01GI0714, 01GI0715, 01GI0716; grants Health Service Research Initiative: 01GY1322A, 01GY1322B, 01GY1322C, 01GY1322D, 01GY1322E, 01GY1322F, 01GY1322G). VITA study: The support of the Ludwig Boltzmann Society and the AFI Germany have supported the VITA study. The former VITA study group should be acknowledged: W. Danielczyk, G. Gatterer, K Jellinger, S Jugwirth, KH Tragl, S Zehetmayer. Vogel Study: This work was financed by a research grant of the ‘‘Vogelstiftung Dr. Eckernkamp’’. HELIAD study: This study was supported by the grants: IIRG-09-133014 from the Alzheimer’s Association, 189 10276/8/9/2011 from the ESPA-EU program Excellence Grant (ARISTEIA) and the ΔΥ2β/οικ.51657/14.4.2009 of the Ministry for Health and Social Solidarity (Greece). Biobank Department of Psychiatry, UMG: Prof. Jens Wiltfang is supported by an Ilídio Pinho professorship and iBiMED (UID/BIM/04501/2013), and FCT project PTDC/DTP_PIC/5587/2014 at the University of Aveiro, Portugal. Lausanne study: This work was supported by grants from the Swiss National Research Foundation (SNF 320030_141179). PAGES study: Harald Hampel is an employee of Eisai Inc. During part of this work he was supported by the AXA Research Fund, the “Fondation partenariale Sorbonne Université” and the “Fondation pour la Recherche sur Alzheimer”, Paris, France. Mannheim,

Germany Biobank: Department of geriatric Psychiatry, Central Institute for Mental Health, Mannheim, University of Heidelberg, Germany. Genotyping for the Swedish Twin Studies of Aging was supported by NIH/NIA grant R01 AG037985. Genotyping in TwinGene was supported by NIH/NIDDK U01 DK066134. WvdF is recipient of Joint Programming for Neurodegenerative Diseases (JPND) grants PERADES (ANR-13-JPRF-0001) and EADB (733051061). Gothenburg Birth Cohort (GBC) Studies: We would like to thank UCL Genomics for performing the genotyping analyses. The studies were supported by The Stena Foundation, The Swedish Research Council (2015-02830, 2013-8717), The Swedish Research Council for Health, Working Life and Wellfare (2013-1202, 2005-0762, 2008-1210, 2013-2300, 2013-2496, 2013-0475), The Brain Foundation, Sahlgrenska University Hospital (ALF), The Alzheimer's Association (IIRG-03-6168), The Alzheimer's Association Zenith Award (ZEN-01-3151), Eivind och Elsa K:son Sylvans Stiftelse, The Swedish Alzheimer Foundation. Clinical AD, Sweden: We would like to thank UCL Genomics for performing the genotyping analyses. Barcelona Brain Biobank: Brain Donors of the Neurological Tissue Bank of the Biobanc-Hospital Clinic-IDIBAPS and their families for their generosity. Hospital Clínic de Barcelona Spanish Ministry of Economy and Competitiveness-Instituto de Salud Carlos III and Fondo Europeo de Desarrollo Regional (FEDER), Unión Europea, "Una manera de hacer Europa" grants (PI16/0235 to Dr. R. Sánchez-Valle and PI17/00670 to Dr. A.Antonelli). AA is funded by Departament de Salut de la Generalitat de Catalunya, PERIS 2016-2020 (SLT002/16/00329). Work at JP-T laboratory was possible thanks to funding from Ciberned and generous gifts from Consuelo Cervera Yuste and Juan Manuel Moreno Cervera. Sydney Memory and Ageing Study (Sydney MAS): We gratefully acknowledge and thank the following for their contributions to Sydney MAS: participants, their supporters and the Sydney MAS Research Team (current and former staff and students). Funding was awarded from the Australian National Health and Medical Research Council (NHMRC) Program Grants (350833, 568969, 109308). This work was supported by InnoMed (Innovative Medicines in Europe), an integrated project funded by the European Union of the Sixth Framework program priority (FP6-2004- LIFESCIHEALTH-5). Oviedo: This work was partly supported by Grant from Fondo de Investigaciones Sanitarias-Fondos FEDER EuropeanUnion to V.A. PI15/00878. Project MinE: The ProjectMinE study was supported by the ALS Foundation Netherlands and the MND association (UK) (Project MinE, www.projectmine.com). The SPIN cohort: We are indebted to patients and their families for their participation in the "Sant Pau Initiative on Neurodegeneration cohort", at the Sant Pau Hospital (Barcelona). This is a multimodal research cohort for biomarker discovery and validation that is partially funded by Generalitat de Catalunya (2017 SGR 547 to JC), as well as from the Institute of Health Carlos III-Subdirección General de Evaluación and the Fondo Europeo de Desarrollo Regional (FEDER- "Una manera de Hacer Europa") (grants PI11/02526, PI14/01126, and PI17/01019 to JF; PI17/01895 to AL), and the Centro de Investigación Biomédica en Red Enfermedades Neurodegenerativas programme (Program 1, Alzheimer Disease to AL). We would also like to thank the Fundació Bancària Obra Social La Caixa (DABNI project) to JF and AL; and Fundación BBVA (to AL), for their support in funding this follow-up study. Adolfo López de Munain is supported by Fundación Salud 2000 (PI2013156), CIBERNED and Diputación Foral de Gipuzkoa (Exp.114/17). P.S.J. is supported by CIBERNED and Carlos III Institute of Health, Spain (PI08/0139, PI12/02288, and PI16/01652, PI20/01011), jointly funded by Fondo Europeo de Desarrollo Regional (FEDER), Unión Europea, "Una manera de hacer Europa". We thank Biobanco Valdecilla for their support. Dr. Fermín Moreno is supported by the Tau Consortium and has received funding from the Carlos III Health Institute (PI19/01637). Amsterdam dementia Cohort (ADC): Research of the Alzheimer center Amsterdam is part of the neurodegeneration research program of Amsterdam Neuroscience. The AlzheimerCenter Amsterdam is supported by Stichting Alzheimer Nederland and Stichting VUmc fonds. The clinical database structure was developed with funding from Stichting Dioraphte. Genotyping of the Dutch case-control samples was performed in the context of EADB (European Alzheimer&Dementia biobank) funded by the JPco-fuND FP-829-029 (ZonMW project number #733051061). This research is performed by using data from the Parelsnoer Institute an initiative of the Dutch Federation of University Medical Centres (www.parelsnoer.org). 100-Plus study: We are grateful for the collaborative efforts of all participating centenarians and their family members and/or relations. We thank the Netherlands Brain Bank for supplying DNA for genotyping. SvdL was funded for this study by NWO (#733050512, PROMO-GENODE: a PROspective study of MOnoGEnic causes Of Dementia) a substantial donation by Edwin Bouw Fonds and Dioraphte. Research of Alzheimer Center Amsterdam is part of the neurodegeneration research program of Amsterdam Neuroscience. This work was supported by Stichting AlzheimerNederland (WE09.2014-03), Stichting Diorapthe, Horstingstuit foundation, Memorabel (ZonMW project number #733050814, #733050512) and Stichting VUmcFonds. Additional support for EADB cohorts: WF, SL, HH are recipients of ABOARD, a public-private partnership receiving funding from ZonMW (#73305095007) and Health~Holland, Topsector Life Sciences & Health (PPP-allowance; #LSHM20106). The DELCODE study was funded by the German Center for Neurodegenerative Diseases (Deutsches Zentrum für Neurodegenerative Erkrankungen (DZNE)), reference number BN012.

**Gra@ce**. We would like to thank patients and controls who participated in this project. The Genome Research @ Ace Alzheimer Center Barcelona project (GR@ACE) is supported by Grifols SA, Fundación bancaria 'La Caixa', Ace Alzheimer Center Barcelona and CIBERNED. Ace Alzheimer Center Barcelona is one of the participating centers of the Dementia Genetics Spanish Consortium (DEGESCO). AR and MB receive support from the European Union / EFPIA Innovative Medicines Initiative joint undertaking ADAPTED and MOPEAD projects (grant numbers 115975 and 115985, respectively). MB and AR are also supported by national grants PI13/02434, PI16/01861, PI17/01474, PI19/01240, PI19/01301 and PI22/01403. Acción Estratégica en Salud is integrated into the Spanish National R+D+I Plan and funded by ISCIII –Subdirección General de Evaluación and the Fondo Europeo de Desarrollo Regional (FEDER–'Una manera de hacer Europa'). AR is also funded by JPco-fuND-2 "Multinational research projects on Personalized Medicine for Neurodegenerative Diseases", PREADAPT project (ISCIII grant: AC19/00097), EURONANOMED III Joint Transnational call for proposals (2017) for European Innovative Research & Technological Development Projects in Nanomedicine (ISCIII grant: AC17/00100), the ISCIII national grant PMP22/00022, funded by the European Union (NextGenerationEU), The support of CIBERNED (ISCIII) under the grants CB06/05/2004 and CB18/05/00010. The support from the ADAPTED and MOPEAD projects, European Union/EFPIA Innovative Medicines Initiative Joint (grant numbers 115975 and 115985, respectively); from PREADAPT project, Joint Program for Neurodegenerative Diseases (JPND) grant Nº AC19/00097; from HARPONE project, Agency for Innovation and Entrepreneurship (VLAIO) grant Nº PR067/21, Janssen. And DESCARTES project funded by German Research Foundation (DFG).
I.dR. is supported by a national grant from the Instituto de Salud Carlos III FI20/00215. Ace Alzheimer Center Barcelona research receives support from Roche, Janssen, Life Molecular Imaging, Araclon Biotech, Alkahest, Laboratorio de Análisis Echevarne, and IrsiCaixa. Some control samples and data from patients included in this study were provided in part by the National DNA Bank Carlos III (www.bancoadn.org, University of Salamanca, Spain) and Hospital Universitario Virgen de Valme (Sevilla, Spain); they were processed following standard operating procedures with the appropriate approval of the Ethical and Scientific Committee.

**EADI**. This work has been developed and supported by the LABEX (laboratory of excellence program investment for the future) DISTALZ grant (Development of Innovative Strategies for a Transdisciplinary approach to ALZheimer's disease) including funding from MEL (Metropole européenne de Lille), ERDF (European Regional Development Fund) and Conseil Régional Nord Pas de Calais. This work was supported by INSERM, the National Foundation for

Alzheimer's disease and related disorders, the Institut Pasteur de Lille and the Centre National de Recherche en Génomique Humaine, CEA, the JPND PERADES, the Laboratory of Excellence GENMED (Medical Genomics) grant no. ANR-10-LABX-0013 managed by the National Research Agency (ANR) part of the Investment for the Future program, and the FP7 AgedBrainSysBio. The Three-City Study was performed as part of collaboration between the Institut National de la Santé et de la Recherche Médicale (Inserm), the Victor Segalen Bordeaux II University and Sanofi-Synthélabo. The Fondation pour la Recherche Médicale funded the preparation and initiation of the study. The 3C Study was also funded by the Caisse Nationale Maladie des Travailleurs Salariés, Direction Générale de la Santé, MGEN, Institut de la Longévité, Agence Française de Sécurité Sanitaire des Produits de Santé, the Aquitaine and Bourgogne Regional Councils, Agence Nationale de la Recherche, ANR supported the COGINUT and COVADIS projects. Fondation de France and the joint French Ministry of Research/INSERM "Cohortes et collections de données biologiques" programme. Lille Génopôle received an unconditional grant from Eisai. The Three-city biological bank was developed and maintained by the laboratory for genomic analysis LAG-BRC - Institut Pasteur de Lille.

**GERAD/PERADES.** We thank all individuals who participated in this study. Cardiff University was supported by the Wellcome Trust, Alzheimer's Society (AS; grant RF014/164), the Medical Research Council (MRC; grants G0801418/1, MR/K013041/1, MR/L023784/1), the European Joint Programme for Neurodegenerative Disease (JPND, grant MR/L501517/1), Alzheimer's Research UK (ARUK, grant ARUK-PG2014-1), Welsh Assembly Government (grant SGR544:CADR), a donation from the Moondance Charitable Foundation, UK Dementia's Platform (DPUK, reference MR/L023784/1), and the UK Dementia Research Institute at Cardiff. Cambridge University acknowledges support from the MRC. ARUK supported sample collections at the Kings College London, the South West Dementia Bank, Universities of Cambridge, Nottingham, Manchester and Belfast. King's College London was supported by the NIHR Biomedical Research Centre for Mental Health and Biomedical Research Unit for Dementia at the South London and Maudsley NHS Foundation Trust and Kings College London and the MRC. Alzheimer's Research UK (ARUK) and the Big Lottery Fund provided support to Nottingham University. Ulster Garden Villages, AS, ARUK, American Federation for Aging Research, NI R&D Office and the Royal College of Physicians/Dunhill Medical Trust provided support for Queen's University, Belfast. The University of Southampton acknowledges support from the AS. The MRC and Mercer's Institute for Research on Ageing supported the Trinity College group. DCR is a Wellcome Trust Principal Research fellow. The South West Dementia Brain Bank acknowledges support from Bristol Research into Alzheimer's and Care of the Elderly. The Charles Wolfson Charitable Trust supported the OPTIMA group. Washington University was funded by NIH grants, Barnes Jewish Foundation and the Charles and Joanne Knight Alzheimer's Research Initiative. Patient recruitment for the MRC Prion Unit/UCL Department of Neurodegenerative Disease collection was supported by the UCLH/UCL Biomedical Research Centre and their work was supported by the NIHR Queen Square Dementia BRU, the Alzheimer's Research UK and the Alzheimer's Society. LASER-AD was funded by Lundbeck SA. The AgeCoDe study group was supported by the German Federal Ministry for Education and Research grants 01 GI 0710, 01 GI 0712, 01 GI 0713, 01 GI 0714, 01 GI 0715, 01 GI 0716, 01 GI 0717. Genotyping of the Bonn case-control sample was funded by the German centre for Neurodegenerative Diseases (DZNE), Germany. The GERAD Consortium also used samples ascertained by the NIMH AD Genetics Initiative. HH was supported by a grant of the Katharina-Hardt-Foundation, Bad Homburg vor der Höhe, Germany. The KORA F4 studies were financed by Helmholtz Zentrum München; German Research Center for Environmental Health; BMBF; German National Genome Research Network and the Munich Center of Health Sciences. The Heinz Nixdorf Recall cohort was funded by the Heinz Nixdorf Foundation and BMBF. We acknowledge use of genotype data from the 1958 Birth Cohort collection and National Blood Service, funded by the MRC and the Wellcome Trust which was genotyped by the Wellcome Trust Case Control Consortium and the Type-1 Diabetes Genetics Consortium, sponsored by the National Institute of Diabetes and Digestive and Kidney Diseases,

National Institute of Allergy and Infectious Diseases, National Human Genome Research Institute, National Institute of Child Health and Human Development and Juvenile Diabetes Research Foundation International. The project is also supported through the following funding organisations under the aegis of JPND - www.jpnd.eu (United Kingdom, Medical Research Council (MR/L501529/1; MR/R024804/1) and Economic and Social Research Council (ES/L008238/1)) and through the Motor Neurone Disease Association. This study represents independent research part funded by the National Institute for Health Research (NIHR) Biomedical Research Centre at South London and Maudsley NHS Foundation Trust and King's College London. Prof Jens Wiltfang is supported by an Ilídio Pinho professorship and iBiMED (UID/BIM/04501/2013), at the University of Aveiro, Portugal.

**DemGene**. The project has received funding from The Research Council of Norway (RCN) Grant Nos. 213837, 223273, 225989, 248778, and 251134 and EU JPND Program RCN Grant Nos. 237250, 311993, the South-East Norway Health Authority Grant No. 2013-123, the Norwegian Health Association, and KG Jebsen Foundation. The RCN FRIPRO Mobility grant scheme (FRICON) is co-funded by the European Union's Seventh Framework Programme for research, technological development and demonstration under Marie Curie grant agreement No 608695. European Community's grant PIAPP-GA-2011-286213 PsychDPC..

**Bonn study**. This group would like to thank Dr. Heike Koelsch for her scientific support. The Bonn group was funded by the German Federal Ministry of Education and Research (BMBF): Competence Network Dementia (CND) grant number 01GI0102, 01GI0711, 01GI042

***China.*** This Chinese AD WGS cohort was supported in part by the National Key R&D Program of China (2021YFE0203000); the Areas of Excellence Scheme of the University Grants Committee (AoE/M-604/16); the Research Grants Council of Hong Kong (the Collaborative Research Fund [C6027-19GF] and the Theme-Based Research Scheme [T13-605/18W]); the Innovation and Technology Commission (InnoHK Funding Scheme; ITCPD/17-9, ITS/207/18FP, MRP/042/18X and MRP/097/20X); Chow Tai Fook Charity Foundation; the Guangdong Provincial Key S&T Program Grant (2018B030336001); the Guangdong Provincial Fund for Basic and Applied Basic Research (2019B1515130004); and the Fundamental Research Program of Shenzhen Virtual University Park (2021Szvup137).

***Japan.*** This study is supported by AMED JP23dk0207060 and JP23wm0525019.

***Korea (GARD).*** This research was supported by a National Research Foundation of Korea grant, funded by the Korean government (MSIT) (No. 2022R1A2C2009998); the ICT Creative Consilience program (No. IITP-2022-2020-0-01821) supervised by the Institute for Information & Communications Technology Planning & Evaluation (IITP); the Korea Health Technology R&D Project through the Korea Health Industry Development Institute, funded by the Ministry of Health and Welfare, Republic of Korea (Nos. HU22C0042, HU21C0111, and HI19C1132); the Ministry of Science and ICT, Republic of Korea (No. 2019RIA5A2026045); the National Institute of Health research project (No. 2021-ER1006-01); and Future Medicine 20*30 Project of the Samsung Medical Center (No. SMX1220021). This study was provided with biospecimens and data from the biobank of Chronic Cerebrovascular Disease consortium. The consortium was supported and funded by the Korea Centers for Disease Control and Prevention (No. 4845-303). This research was also supported by the Basic Research Program through the Korea Brain Research Institute (No. 23-BR-03.05), the Healthcare AI Convergence Research & Development Program through the National IT Industry Promotion Agency of Korea (No. 1711196566) funded by the Ministry of Science and ICT, and Global-LAMP Program (No. RS-2023-00285353) through the National Research Foundation of Korea funded by the Ministry of Education

***Sub Saharan Africa (EPIDEMCA).*** ***Sub Saharan Africa (EPIDEMCA).*** This study was supported by a grant from the French National Research Agency (ANR-09-MNPS-009-01),the

AXA research Fund (2012 – Project – Public Health Institute (Inserm) – PREUX Pierre-Marie) and the Limoges University Hospital through its APREL scheme. The sponsors of the study had no role in study design, data collection, data interpretation or writing of the report.
We also thank the staffs of the Universities of Bangui (Central African Republic) and Marien Ngouabi in Brazzaville (ROC); Pasteur Institute in Bangui and “Laboratoire National de Santé Publique” in Brazzaville; Health ministries of the Central African Republic and the Republic of Congo, for their moral support; University of Limoges, Doctoral School of Limoges University, Inserm; Limousin Regional Council. We are very grateful to all the participants to this survey, the investigators, and staffs of Bangui and Brazzaville hospitals for their assistance

***Argentina.*** This study was supported by funding from Alexander von Humboldt Foundation and International Society for Neurochemistry (ISN) to M.C.D.; the Agencia Nacional de Promoción Científica y Tecnológica (PID-2011-0059, PIBT/09-2013, PICT-2016-4647 and PICT2019-0656 to L.M.) and from EU-LAC Health-Neurodegeneration JOINT CALL 2016 (EULACH16 to L.M.).

***Chile.*** The Funding of the ALEXANDROS study was provided by the Chilean National Fund for Science and Technology (FONDECYT) grant 1130947. The genotyping for the Chilean and Tunisian series were funded by Genome Research @ Ace Alzheimer Center Barcelona project (GR@ACE), supported by Grifols SA, Fundación bancaria ‘La Caixa’, Ace Alzheimer Center Barcelona and CIBERNED.

***Brazil.*** The study was supported by the Brazilian National Council for Scientific and Technological Development (CNPq): processes 315133/2021-0, 309953/2018-9, 436735/2018-0) FAPEMIG:processes:APQ-02662-14, APQ-02662-14) and the Coordination for the Improvement of Higher Education Personnel (CAPES): process 88887.569376/2020-00.

***Colombia.*** This was supported by Ministerio de Ciencia, Tecnología e Innovación (MINCIENCIAS) and the Universidad Nacional de Colombia. We also thanks all the participants who kindly agreed to participate in the study.

***ADSP.*** The ADGC cohorts include Adult Changes in Thought (ACT) (U01 AG006781, U19 AG066567),the Alzheimer’s Disease Research Centers (ADRC) (P30AG062429, P30 AG066468, P30AG062421, P30AG066509, P30AG066514, P30AG066530, P30AG066507, P30AG066444, P30AG066518, P30AG066512, P30AG066462, P30AG072979, P30AG072972, P30AG072976, P30AG072975, P30AG072978, P30AG072977, P30AG066519, P30AG062677, P30AG079280, P30AG062422, P30AG066511, P30AG072946, P30AG062715, P30AG072973, P30AG066506, P30AG066508, P30AG066515, P30AG072947, P30AG072931, P30AG066546, P20AG068024, P20AG068053, P20AG068077, P20AG068082, P30AG072958, P30AG072959), the Chicago Health and Aging Project (CHAP) (R01 AG11101, RC4 AG039085, K23 AG030944), Indiana Memory and Aging Study (IMAS) (R01 AG019771), Indianapolis Ibadan (R01 AG009956, P30 AG010133), the Memory and Aging Project (MAP) ( R01 AG17917), Mayo Clinic (MAYO) (R01 AG032990, U01AG046139, R01 NS080820, RF1 AG051504, P50 AG016574), Mayo Parkinson’s Disease controls (NS039764, NS071674, 5RC2HG005605), University of Miami (R01 AG027944, R01 AG028786, R01 AG019085, IIRG09133827, A2011048), the Multi-Institutional Research in Alzheimer’s Genetic Epidemiology Study (MIRAGE) (R01 AG09029, R01 AG025259), the National Centralized Repository for Alzheimer’s Disease and Related Dementias (NCRAD) (U24 AG021886), the National Institute on Aging Late Onset Alzheimer’s Disease Family Study (NIA- LOAD) (U24AG056270), the Religious Orders Study (ROS) (P30 AG10161, R01 AG15819), the Texas Alzheimer’s Research and Care Consortium (TARCC) (funded by the Darrell K Royal Texas Alzheimer’s Initiative), Vanderbilt University/Case Western Reserve University (VAN/CWRU) (R01AG019757, R01 AG021547, R01 AG027944, R01 AG028786, P01 NS026630, and Alzheimer’s Association), the Washington Heights-

Inwood Columbia Aging Project (WHICAP) (RF1AG054023), the University of Washington Families (VA Research Merit Grant, NIA: P50AG005136, R01AG041797, NINDS: R01NS069719), the Columbia University Hispanic Estudio Familiar de Influencia Genetica de Alzheimer (EFIGA) (RF1 AG015473), the University of Toronto (UT)(funded by Wellcome Trust, Medical Research Council, Canadian Institutes of Health Research), and Genetic Differences (GD) (R01 AG007584). The CHARGE cohorts are supported in part by National Heart, Lung, and Blood Institute (NHLBI) infrastructure grant HL105756 (Psaty), RC2HL102419 (Boerwinkle) and the neurology working group is supported by the National Institute on Aging (NIA) R01 grant AG033193. The CHARGE cohorts The CHARGE cohorts participating in the ADSP include the following: Austrian Stroke Prevention Study (ASPS), ASPS-Family study, and the Prospective Dementia Registry-Austria (ASPS/PRODEM-Aus), the Atherosclerosis Risk in Communities (ARIC) Study, the Cardiovascular Health Study (CHS), the Erasmus Rucphen Family Study (ERF), the Framingham Heart Study (FHS), and the Rotterdam Study (RS). ASPS is funded by the Austrian Science Fond (FWF) grant number P20545-P05 and P13180 and the Medical University of Graz. The ASPS-Fam is funded by the Austrian Science Fund (FWF) project I904), the EU Joint Programme – Neurodegenerative Disease Research (JPND) in frame of the BRIDGET project (Austria, Ministry of Science) and theMedical University of Graz and the Steiermärkische Krankenanstalten Gesellschaft. PRODEM-Austria is supported by the Austrian Research Promotion agency (FFG) (Project No.827462) and by the Austrian National Bank (Anniversary Fund, project 15435. ARIC research is .CC-BY-NC-ND 4.0 International licenseIt is made available under a is the author/funder, who has granted medRxiv a license to display the preprint in perpetuity. The collaborative study is also supported by NHLBI contracts (HHSN268201100005C,HHSN268201100006C, HHSN268201100007C, HHSN268201100008C, HHSN268201100009C,Neurocognitive data in ARIC is collected by U01 2U01HL096812, 2U01HL096814, 2U01HL096899, 2U01HL096902,2U01HL096917 from the NIH (NHLBI, NINDS, NIA and NIDCD), and with previous brain MRI examinations funded by R01-HL70825 from the NHLBI. CHS research was supported by contracts HHSN268201200036C, HHSN268200800007C, N01HC55222, N01HC85079, N01HC85080, N01HC85081, N01HC85082, N01HC85083, N01HC85086, and grants U01HL080295 and U01HL130114 from the NHLBI with additional contribution from the National Institute of Neurological Disorders and Stroke (NINDS). Additional support was provided by R01AG023629, R01AG15928, and R01AG20098 from the NIA. FHS research is supported by NHLBI contracts N01-HC-25195 and HHSN268201500001I. This study was also supported by additional grants from the NIA (R01s AG054076, AG049607 and AG033040 and NINDS (R01 NS017950). The ERF study as a part of EUROSPAN (European Special Populations Research Network) was supported by European Commission FP6 STRP grant number 018947 (LSHG-CT-2006-01947) and also received funding from the European Community's Seventh Framework Programme (FP7/2007-2013)/grant agreement HEALTH-F4- 2007-201413 by the European Commission under the programme "Quality of Life and Management of the Living Resources" of 5th Framework Programme (no.QLG2-CT-2002- 01254). High-throughput analysis of the ERF data was supported by a joint grant from the Netherlands Organization for Scientific Research and the Russian Foundation for Basic Research (NWO-RFBR 047.017.043). The Rotterdam Study is funded by Erasmus Medical Center. Genetic data sets are also supported by the Netherlands Organization of Scientific Research NWO Investments (175.010.2005.011,584 911-03-012), the Genetic Laboratory of the Department of Internal Medicine, Erasmus MC, the Research Institute for Diseases in the Elderly (014-93-015; RIDE2), and the Netherlands Genomics Initiative (NGI)/Netherlands Organization for Scientific Research (NWO) Netherlands Consortium for Healthy Aging (NCHA), project 050-060-810. All studies are grateful to their participants, faculty and staff. The content of these manuscripts is solely the responsibility of the authors and does not necessarily represent the official views of the National Institutes of Health or the U.S. Department of Health and Human Services. In addition, funding to support and share the ADSP data are included here: NIA U54-AG052427 and NIA U24-AG041689.

***The Sacramento Area Latino Study on Aging (SALSA).*** This study was funded by the National Institutes of Health, including the National Institute on Aging under Award Numbers K01AG056602 and R01AG12975 and the National Institute for Diabetes and Digestive and Kidney Diseases under Award Numbers R01 DK60753 and R01DK087864.

***MVP.*** This research is based on data from the Million Veteran Program, Office of Research and Development, Veterans Health Administration, and was supported by VA BLR&D grants 1 I01 BX004192 (MVP015) and I01BX005749 (MVP040). The views expressed in this article are those of the authors and do not necessarily reflect the position or policy of the Department of Veterans Affairs or the US government.

#### EADB, Supplementary list of authors

Sonia Moreno-Grau[215,10], Najaf Amin[228,229], Peter A. Holmans[230], Luca Kleineidam[231,232,233], Dag Aarsland[234,235], Pablo Garcia-Gonzalez[215,10], Carla Abdelnour[215,10], Emilio Alarcón-Martín[215,236], Daniel Alcolea[237,10], Montserrat Alegret[215,10], Ignacio Alvarez[238,239], Nicola J. Armstrong[155], Tsolaki Anthoula[55,240], Ildebrando Appollonio[241,242], Marina Arcaro[243], Silvana Archetti[244], Alfonso Arias Pastor[78,79], Lavinia Athanasiu[245], Henri Bailly[134], Nerisa Banaj[246], Miquel Baquero[247], Ana Belén Pastor[248], Luisa Benussi[164], Claudine Berr[249], Céline Besse[250], Valentina Bessi[251,252], Giuliano Binetti[164,253], Alessandra Bizarro[254], Rafael Blesa[237,10], Silvia Boschi[180], Paola Bossù[255], Geir Bråthen[256,257], Catherine Bresner[230], Henry Brodaty[155,258], Keeley J. Brookes[259], Dolores Buiza-Rueda[10,75], Katharina Bûrger[260,261], Vanessa Burholt[262,263], Miguel Calero[10,248,264], Geneviève Chene[131,132], Ángel Carracedo[265,266], Roberta Cecchetti[267], Laura Cervera-Carles[237,10], Camille Charbonnier[268], Caterina Chillotti[269], Simona Ciccone[270], Jordi Clarimon[237,10], Christopher Clark[271], Elisa Conti[241], Anaïs Corma-Gómez[272], Emanuele Costantini[273], Carlo Custodero[274], Delphine Daian[250], Efthimios Dardiotis[275], Peter Paul de Deyn[276], Teodoro del Ser[277], Nicola Denning[278], Janine Diehl-Schmid[279], Mónica Diez-Fairen[238,239], Paolo Dionigi Rossi[270], Srdjan Djurovic[245], Emmanuelle Duron[134], Sebastiaan Engelborghs[280,281,282,283], Ana Espinosa[215,10], Michael Ewers[260,261], Tagliavini Fabrizio[284], Sune Fallgaard Nielsen[285], Lucia Farotti[171], Chiara Fenoglio[286], Marta Fernández-Fuertes[272], Catarina B Ferreira[85], Evelyn Ferri[270], Bertrand Fin[250], Peter Fischer[287], Tormod Fladby[288], Klaus Fließbach[289,233], Juan Fortea[237,10], Tatiana M. Foroud[112], Silvia Fostinelli[164], Nick C. Fox[290], Emlio Franco-Macías[291], Ana Frank-García[10,63,292], Lutz Froelich[293], Jose Maria García-Alberca[10,80], Sebastian Garcia-Madrona[294], Guillermo Garcia-Ribas[294], Ina Giegling[295], Giaccone Giorgio[284], Oliver Goldhardt[279], Antonio González-Pérez[296], Giulia Grande[104], Emma Green[297], Tamar Guetta-Baranes[298], Annakaisa Haapasalo[299], Georgios Hadjigeorgiou[300], Harald Hampel[301,302], John Hardy[303], Annette M. Hartmann[295], Janet Harwood[230], Seppo Helisalmi[304,305], Michael T. Heneka[232,306], Isabel Hernández[215,10], Martin J. Herrmann[307], Per Hoffmann[53], Clive Holmes[308], Raquel Huerto Vilas[78,79], Marc Hulsman[23,309], Geert Jan Biessels[310], Charlotte Johansson[311,312], Lena Kilander[313], Anne Kinhult Ståhlbom[311,312], Miia Kivipelto[314,315,316,317], Anne Koivisto[304,318,319], Johannes Kornhuber[320], Mary H. Kosmidis[321], Carmen Lage[10,82], Erika J Laukka[104,322], Alessandra Lauria[254], Jenni Lehtisalo[304,323], Ondrej Lerch[88,89], Alberto Lleó[237,10], Adolfo Lopez de Munain[10,324], Seth Love[325], Malin Löwemark[313], Lauren Luckcuck[230], Juan Macías[272], Catherine A. MacLeod[326], Wolfgang Maier[327,233], Francesca Mangialasche[314], Marco Spallazzi[328], Marta Marquié[215,10], Rachel Marshall[230], Angel Martín Montes[10,63,292], Carmen Martínez Rodríguez[329], Simon Mead[330], Miguel Medina[10,248], Alun Meggy[278], Silvia Mendoza[80], Manuel Menéndez-González[329], Merel Mol[93], Laura Montrreal[215], Kevin Morgan[331], Markus M Möthen[53], Tiia Ngandu[323], Børge G. Nordestgaard[332,91], Robert Olaso[250], Adelina Orellana[215,10], Michela Orsini[273], Gemma Ortega[215,10], Alessandro Padovani[333], Alba Pérez-Cordón[215], Pierre Pericard[334], Juan A Pineda[272], Claudia Pisanu[335], Thomas Polak[307], Danielle Posthuma[24], Josef Priller[336,337], Olivier Quenez[268], Inés Quintela[265], Alberto Rábano[248,338,10], Marcel J.T. Reinders[339], Peter Riederer[340], Natalia Roberto[215], Arvid Rongve[341,342], Irene Rosas Allende[68,69], Maitée Rosende-Roca[215,10], Jose Luis Royo[343], Elisa Rubino[344], María Eugenia Sáez[296], Paraskevi Sakka[345], Ingvild Saltvedt[257,346], Ángela Sanabria[215,10], María Bernal Sánchez-Arjona[291], Florentino Sanchez-Garcia[347], Pascual Sánchez Juan[82,10], Sigrid B Sando[256,257], Michela Scamosci[267], Elio Scarpini[243,286], Martin Scherer[348], Matthias Schmid[306,349], Jonathan M. Schott[290], Alexey A Shadrin[245], Olivia Skrobot[325], Alina Solomon[304,314], Sandro Sorbi[251,177], Oscar Sotolongo-Grau[215], Annika Spottke[306,350], Eystein Stordal[351], Juan Pablo Tartan[215], Lluís Tárraga[215,10], Niccolo Tesí[23,309], Anbupalam Thalamuthu[155], Tegos Thomas[55], Anne Tybjærg-Hansen[91,352], Andre Uitterlinden[353], Abbe Ullgren[311], Ingun Ulstein[354], Sergi Valero[215,10], Christine Van Broeckhoven[355,356,357], Jasper Van Dongen[27,355,356], Rik Vandenberghe[358,359], Jean-Sébastien Vidal[134], Jonathan Vogelgsang[50,360], Michael Wagner[289,306], Leonie Weinhold[349], Gill Windle[326], Bob Woods[326], Mary Yannakoulia[361], Miren Zulaica[10,362]

[228]Department of Epidemiology, ErasmusMC

[229]Nuffield Department of Population Health Oxford University

[230]MRC Centre for Neuropsychiatric Genetics and Genomics, , Division of Psychological Medicine and Clinical Neuroscience, School of Medicine, Cardiff University, Cardiff, UK

[231]Division of Neurogenetics and Molecular Psychiatry, Department of Psychiatry and Psychotherapy, University of Cologne, Medical Faculty, 50937 Cologne, Germany.

[232]Department of Neurodegeneration and Geriatric Psychiatry, University of Bonn, Bonn, Germany.

[233]German Center for Neurodegenerative Diseases (DZNE Bonn), Bonn, Germany

[234]Centre of Age-Related Medicine, Stavanger University Hospital, Norway

[235]Institute of Psychiatry, Psychology & Neuroscience, PO 70, 16 De Crespigny Park, London, SE58AF

[236]Department of Surgery, Biochemistry and Molecular Biology, School of Medicine, University of Málaga, Málaga, Spain.

[237]Department of Neurology, II B Sant Pau, Hospital de la Santa Creu i Sant Pau, Universitat Autònoma de Barcelona, Barcelona, Spain.

[238]Fundació Docència i Recerca MútuaTerrassa and Movement Disorders Unit, Department of Neurology, University Hospital MútuaTerrassa, Terrassa 08221, Barcelona, Spain

[239]Memory Disorders Unit, Department of Neurology, Hospital Universitari Mutua de Terrassa, Terrassa, Barcelona, Spain.

[240]Alzheimer Hellas, Thessaloniki, Makedonia, Greece

[241]School of Medicine and Surgery, University of Milano-Bicocca, Italy

[242]Neurology Unit, "San Gerardo" hospital, Monza, Italy
[243]Fondazione IRCCS Ca' Granda, Ospedale Policlinico, Milan, Italy
[244]Department of Laboratory Diagnostics, III Laboratory of Analysis, Brescia Hospital, Brescia, Italy
[245]NORMENT Centre, University of Oslo, Oslo, Norway
[246]Laboratory of Neuropsychiatry, Department of Clinical and Behavioral Neurology, IRCCS Santa Lucia Foundation, Rome, Italy
[247]Servei de Neurologia, Hospital Universitari i Politècnic La Fe, Valencia, Spain.
[248]CIEN Foundation/Queen Sofia Foundation Alzheimer Center
[249]Univ. Montpellier, Inserm U1061, Neuropsychiatry: epidemiological and clinical research, PSNREC, Montpellier, France
[250]Université Paris-Saclay, CEA, Centre National de Recherche en Génomique Humaine, 91057, Evry, France
[251]Department of Neuroscience, Psychology, Drug Research and Child Health University of Florence, Florence Italy
[252]Azienda Ospedaliero-Universitaria Careggi, , Florence, Italy
[253]MAC - Memory Clinic, IRCCS Istituto Centro San Giovanni di Dio Fatebenefratelli, Brescia
[254]Geriatrics Unit Fondazione Policlinico A. Gemelli IRCCS, Rome , Italy
[255]Experimental Neuro-psychobiology Laboratory,Department of Clinical and Behavioral Neurology, IRCCS Santa Lucia Foundation, Rome, Italy
[256]Department of Neurology and Clinical Neurophysiology, University Hospital of Trondheim, Trondheim, Norway
[257]Department of Neuromedicine and Movement Science, Norwegian University of Science and Technology, Trondheim, Norway
[258]Dementia Centre for Research Collaboration, School of Psychiatry, University of New South Wales, Sydney, Australia
[259]Biosciences, School of Science and Technology, Nottingham Trent University, Nottingham UK
[260]Institute for Stroke and Dementia Research, Klinikum der Universität München, Ludwig-Maximilians-Universität LMU, Munich, Germany.
[261]German Center for Neurodegenerative Diseases (DZNE, Munich), Munich, Germany.
[262]Faculty of Medical & Health Sciences, University of Auckland, New Zealand
[263]Wales Centre for Ageing & Dementia Research, Swansea University, Wales, New Zealand
[264]UFIEC, Instituto de Salud Carlos III, , Madrid, Spain
[265]Grupo de Medicina Xenómica, Centro Nacional de Genotipado (CEGEN-PRB3-ISCIII). Universidade de Santiago de Compostela, Santiago de Compostela, Spain.
[266]Fundación Pública Galega de Medicina Xenómica- CIBERER-IDIS, University of Santiago de Compostela, Santiago de Compostela, Spain.
[267]Institute of Gerontology and Geriatrics, Department of Medicine and Surgery, University of Perugia Perugia (Italy)
[268]Normandie Univ, UNIROUEN, Inserm U1245 and CHU Rouen, Department of Genetics and CNR-MAJ, Rouen, France
[269]Unit of Clinical Pharmacology, University Hospital of Cagliari, Cagliari, Italy
[270]Geriatic Unit, Fondazione Cà Granda, IRCCS Ospedale Maggiore Policlinico, Milan, Italy
[271]Institute for Regenerative Medicine, University of Zürich, Schlieren, Switzerland
[272]Unidad Clínica de Enfermedades Infecciosas y Microbiología. Hospital Universitario de Valme, Sevilla, Spain
[273]Department of Neuroscience, Catholic University of Sacred Heart, Fondazione Policlinico Universitario A. Gemelli IRCCS, Rome, Italy
[274]University of Bari, “A. Moro”, Italy
[275]School of Medicine, University of Thessaly, Larissa, Greece
[276]Department of Neurology, University Medical Center Groningen, the Netherlands
[277]Department of Neurology/CIEN Foundation/Queen Sofia Foundation Alzheimer Center
[278]UKDRI@ Cardiff, School of Medicine, Cardiff University, Cardiff, UK
[279]Technical University of Munich, School of Medicine, Klinikum rechts der Isar, Department of Psychiatry and Psychotherapy
[280]Center for Neurosciences, Vrije Universiteit Brussel (VUB), Brussels, Belgium
[281]Reference Center for Biological Markers of Dementia (BIODEM), Institute Born-Bunge, University of Antwerp, Antwerp, Belgium
[282]Institute Born-Bunge, University of Antwerp, Antwerp, Belgium
[283]Department of Neurology, UZ Brussel, Brussels, Belgium
[284]Fondazione IRCCS, Istituto Neurologico Carlo Besta, Milan Italy
[285]Department of Clinical Biochemistry, Herlev and Gentofte Hospital, Herlev Denmark
[286]University of Milan, Milan, Italy
[287]Department of Psychiatry, Social Medicine Center East- Donauspital, Vienna, Austria
[288]Institute of Clinical Medicine, University of Oslo, Oslo, Norway.
[289]Department of Neurodegeneration and Geriatric Psychiatry, University of Bonn, 53127 Bonn, Germany.
[290]Dementia Research Centre, UCL Queen Square Institute of Neurology, London, United Kingdom
[291]Unidad de Demencias, Servicio de Neurología y Neurofisiología. Instituto de Biomedicina de Sevilla (IBiS), Hospital Universitario Virgen del Rocío/CSIC/Universidad de Sevilla, Seville, Spain
[292]Hospital Universitario la Paz, Madrid, Spain
[293]Department of geriatric Psychiatry, Central Institute for Mental Health, Mannheim, University of Heidelberg, Germany
[294]Hospital Universitario Ramon y Cajal, IRYCIS, Madrid
[295]Department of Psychiatry and Psychotherapy, Medical University of Vienna, Vienna, Austria
[296]CAEBI, Centro Andaluz de Estudios Bioinformáticos, Sevilla, Spain.
[297]Institute of Public Health, University of Cambridge, UK
[298]Human Genetics, School of Life Sciences, Life Sciences Building, University Park, University of Nottingham, Nottingham, UK
[299]A.I Virtanen Institute for Molecular Sciences, University of Eastern Finland, Kuopio, Finland
[300]Department of Neurology, Medical School, University of Cyprus, Cyprus
[301]Sorbonne University, GRC n° 21, Alzheimer Precision Medicine Initiative (APMI), AP-HP, Pitié-Salpêtrière Hospital, Boulevard de l'hôpital, F-75013, Paris, France

[302]Eisai Inc., Neurology Business Group, 100 Tice Blvd, Woodcliff Lake, NJ 07677, USA
[303]Reta Lila Weston Research Laboratories, Department of Molecular Neuroscience, UCL Institute of Neurology, London, UK.
[304]Insitute of Clinical Medicine - Neurology, University of Eastern, Kuopio, Finland
[305]Institute of Clinical Medicine – Internal Medicine, University of Eastern Finland, Kuopio, Finland
[306]German Center for Neurodegenerative Diseases (DZNE, Bonn), Bonn, Germany.
[307]Department of Psychiatry, Psychosomatics and Psychotherapy, Center of Mental Health, University Hospital, Wuerzburg
[308]Clinical and Experimental Science, Faculty of Medicine, University of Southampton, Southampton, UK.
[309]Section Genomics of Neurdegenerative Diseases and Aging, Department of Human Genetics Amsterdam UMC, Vrije Universiteit Amsterdam, Amsterdam UMC, Amsterdam, The Netherlands
[310]Department of Neurology, UMC Utrecht Brain Center, Utrecht, the Netherlands
[311]Karolinska Institutet, Center for Alzheimer Research, Department NVS, Division of Neurogeriatrics, Stockholm, Sweden
[312]Unit for Hereditary dementias, Karolinska University Hospital-Solna, Stockholm, Sweden
[313]Dept.of Public Health and Carins Sciences / Geriatrics, Uppsala University
[314]Division of Clinical Geriatrics, Center for Alzheimer Research, Care Sciences and Society (NVS), Karolinska Institutet, Stockholm, Sweden
[315]Institute of Public Health and Clinical Nutrition, University of Eastern Finland, Kuopio, Finland
[316]Neuroepidemiology and Ageing Research Unit, School of Public Health, Imperial College London, London, United Kingdom
[317]Stockholms Sjukhem, Research & Development Unit, Stockholm, Sweden
[318]Department of Neurology, Kuopio University Hospital, Kuopio, Finland
[319]Department of Neurosciences, University of Helsinki and Department of Geriatrics, Helsinki University Hospital, Helsinki, Finland
[320]Department of Psychiatry and Psychotherapy, Universitätsklinikum Erlangen, and Friedrich-Alexander Universität Erlangen-Nürnberg, Erlangen, Germany.
[321]Laboratory of Cognitive Neuroscience, School of Psychology, Aristotle University of Thessaloniki, Thessaloniki, Greece
[322]Stockholm Gerontology Research Center, Stockholm, Sweden
[323]Public Health Promotion Unit, Finnish Institute for Health and Welfare, Helsinki, Finland
[324]Department of Neurology. Hospital Universitario Donostia. OSAKIDETZA-Servicio Vasco de Salud, San Sebastian, Spain
[325]Translational Health Sciences, Bristol Medical School, University of Bristol, Bristol, BS16 1LE, UK
[326]School of Health Sciences, Bangor University, UK
[327]Department of Neurodegenerative Diseases and Geriatric Psychiatry, University Hospital Bonn, Bonn, Germany
[328]Unit of Neurology, University of Parma and AOU, Parma, Italy
[329]Servicio de Neurología HOspital Universitario Central de Asturias- Oviedo and Instituto de Investigación Biosanitaria del Principado de Asturias, Oviedo, Spain
[330]MRC Prion Unit at UCL, UCL Institute of Prion Diseases, London, W1W 7FF
[331]Human Genetics, School of Life Sciences, University of Nottingham, UK NG7 2UH
[332]Department of Clinical Biochemistry, Herlev and Gentofte Hospital, Herlev, Denmark
[333]Centre for Neurodegenerative Disorders, Department of Clinical and Experimental Sciences, University of Brescia, Brescia, Italy
[334]Univ. Lille, CNRS, Inserm, CHU Lille, Institut Pasteur de Lille, US 41-UMS 2014-PLBS, bilille, Lille, France.
[335]Department of Biomedical Sciences, University of Cagliari, Italy
[336]German Center for Neurodegenerative Diseases (DZNE), Berlin, Germany.
[337]Department of Neuropsychiatry and Laboratory of Molecular Psychiatry, Charité, Charitéplatz 1, 10117 Berlin, Germany
[338]BT-CIEN
[339]Delft Bioinformatics Lab, Delft University of Technology, Delft, The Netherlands
[340]Center of Mental Health, Clinic and Policlinic of Psychiatry, Psychosomatics and Psychotherapy, University Hospital of Würzburg, Wuerzburg, Germany
[341]Department of Research and Innovation, Helse Fonna, Haugesund Hospital, Haugesund, Norway.
[342]The University of Bergen, Institute of Clinical Medicine (K1), Bergen Norway
[343]Departamento de Especialidades Quirúrgicas, Bioquímicas e Inmunología, School of Medicine, University of Málaga, Málaga, Spain.
[344]Department of Neuroscience and Mental Health, AOU Città della Salute e della Scienza di Torino, Torino, Italy
[345]Athens Association of Alzheimer's disease and Related Disorders, Athens, Greece
[346]Department of Geriatrics, St. Olav's Hospital, Trondheim University Hospital, Norway
[347]Department of Immunology, Hospital Universitario Doctor Negrín, Las Palmas de Gran Canaria, Spain.
[348]Department of Primary Medical Care, University Medical Centre Hamburg-Eppendorf, 20246 Hamburg, Germany.
[349]Institute of Medical Biometry, Informatics and Epidemiology, University Hospital of Bonn, Bonn, Germany.
[350]Department of Neurology, University of Bonn, Bonn, Germany.
[351]Department of Psychiatry, Namsos Hospital, Namsos, Norway
[352]Department of Clinical Biochemistry, Rigshospitalet, Copenhagen, Denmark
[353]Department of Internal medicine and Biostatistics, ErasmusMC
[354]Department of Geriatric Medicine, Oslo University Hospital, Oslo, Norway
[355]Laboratory of Neurogenetics, Institute Born - Bunge, Antwerp, Belgium
[356]Department of Biomedical Sciences, University of Antwerp, Neurodegenerative Brain Diseases Group, Center for Molecular Neurology, VIB, Antwerp, Belgium
[357]Neurodegenerative Brain Diseases Group, VIB Center for Molecular Neurology, VIB, Antwerp, Belgium
[358]Laboratory for Cognitive Neurology, Department of Neurosciences, University of Leuven, Belgium
[359]Neurology Department, University Hospitals Leuven, Leuven, Belgium
[360]Department of Psychiatry, Harvard Medical School, McLean Hospital, Belmont, MA, USA

[361]Department of Nutrition and Diatetics, Harokopio University, Athens, Greece
[362]Neurosciences Area. Instituto Biodonostia. San Sebastian, Spain
