## Supplementary Informations for "Transferability of European-derived Alzheimer’s Disease Polygenic Risk Scores across Multi-Ancestry Populations"

Aude Nicolas et al.

TABLE OF CONTENTS


1. **Sample Description** **2**
2. **Supplementary figures 1-14** **15**
3. **Supplementary References** **29**
4. **Acknowledgements** **32**
5. **Supplementary list of authors** **41**

### 1. SAMPLE DESCRIPTION

Demographics of the different case-control studies are fully described in the supplementary Table 1

***ADSP.*** Alzheimer's Disease Sequencing Project (ADSP) is a collaborative project aiming at identifying new variants, genes, and therapeutic targets in AD[44]. The joint-called whole-genome sequencing data from the Alzheimer's Disease Sequencing Project (ADSP) accession id NG00067.v10 were downloaded from the NIAGADS DSS website (https://dss.niagads.org/datasets/ng00067/), with 36,361 whole-genome genomes[45]. The global ancestry of each individual genome was determined with SNPweights v.2.1[46] using a set of ancestry-weighted variants computed on reference populations from the 1000 Genomes Project as in[47]. By applying a global ancestry percentage cutoff of >75%, the samples were assigned to the 5 super-populations: South-Asians, East-Asians, Americans, Africans, and Europeans, and classified as Admixed ancestries otherwise. Indian samples were selected using their Identifying code "G-LSID" and a percentage of South-Asian genetic ancestry > 50. The African Americans or Native Americans were selected with an African ancestry above 75% or a Native American ancestry above 75%. As the Hispanic/Latinos group is based on a self-report, samples with African, Native American, South Asian or South East Asian genetic ancestries above 50% were filtered out. Then, in each genetic ancestry super-population, PC outliers of one of the first 4 PCs were excluded. The 83 independent variants were extracted from the individual genomes with vcftools v0.1.17[48]. The PCs significantly associated with the disease status, were selected as covariates, in the logistic regressions. The 83 independent variants were extracted from the individual genomes with vcftools v0.1.17[48]. Technical duplicates or twins were identified using KING v2.2.5 (Kinship-based INference for Gwas)[49] and only one genome among the set of identical pairs was retained for further analysis. The phenotype cognitively unimpaired or Alzheimer's disease case was obtained from the ADSP, and participants who did not fall in either of these two categories were excluded from further analysis (see cohorts descriptions at https://dss.niagads.org/datasets/ng00067/). Principal components accounting for genetic ancestry were computed per ancestry group using smartpca[50].

**Supplementary Figure 1:** $PGS^{ALZ}$ distribution (Finland, Sweden, Norway, Denmark, UK, Netherlands)

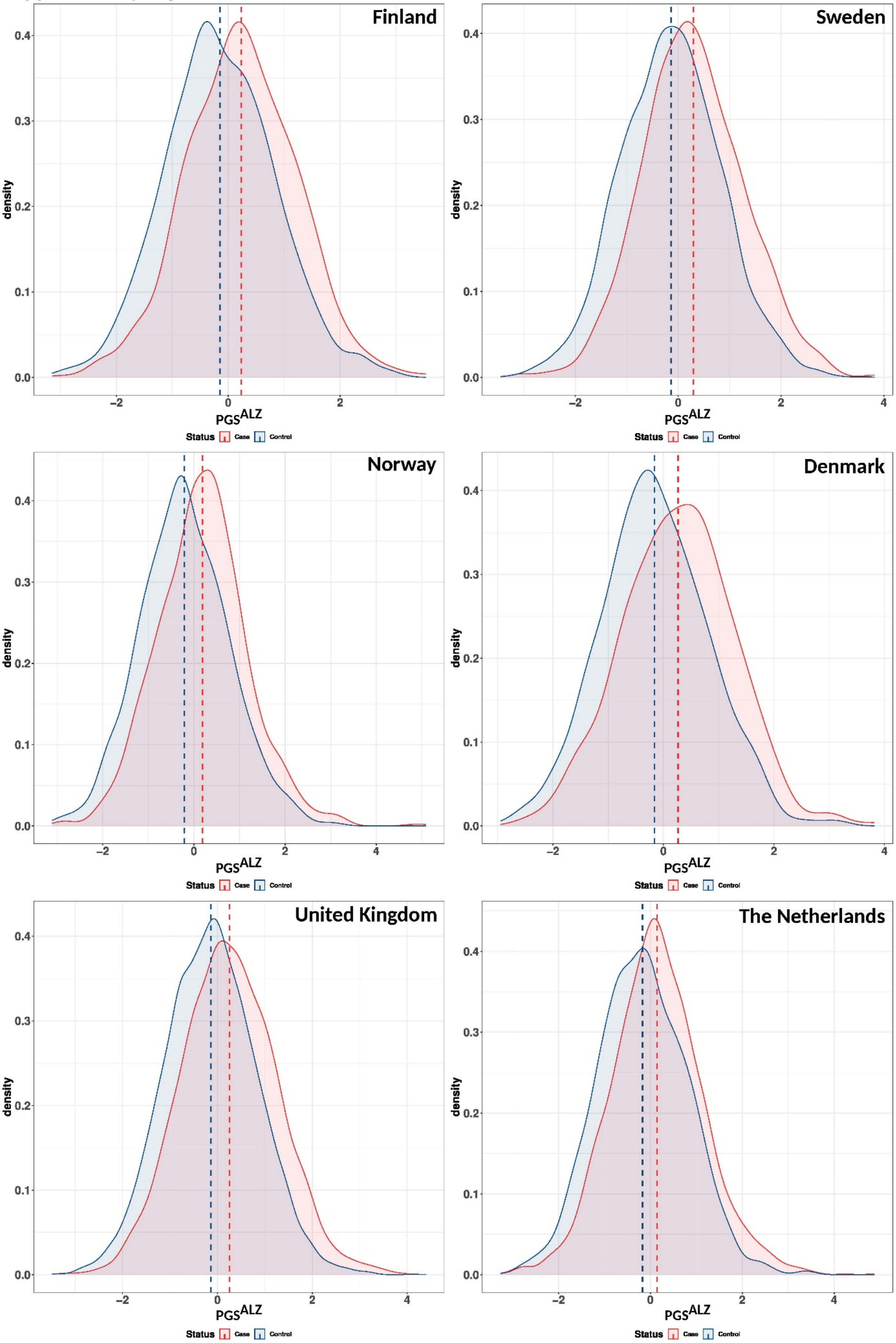

**Supplementary Figure 2:** PGS$^{ALZ}$ distribution (Belgium, Germany, Austria/Switzerland, Czech Republic, Greece, Italy)

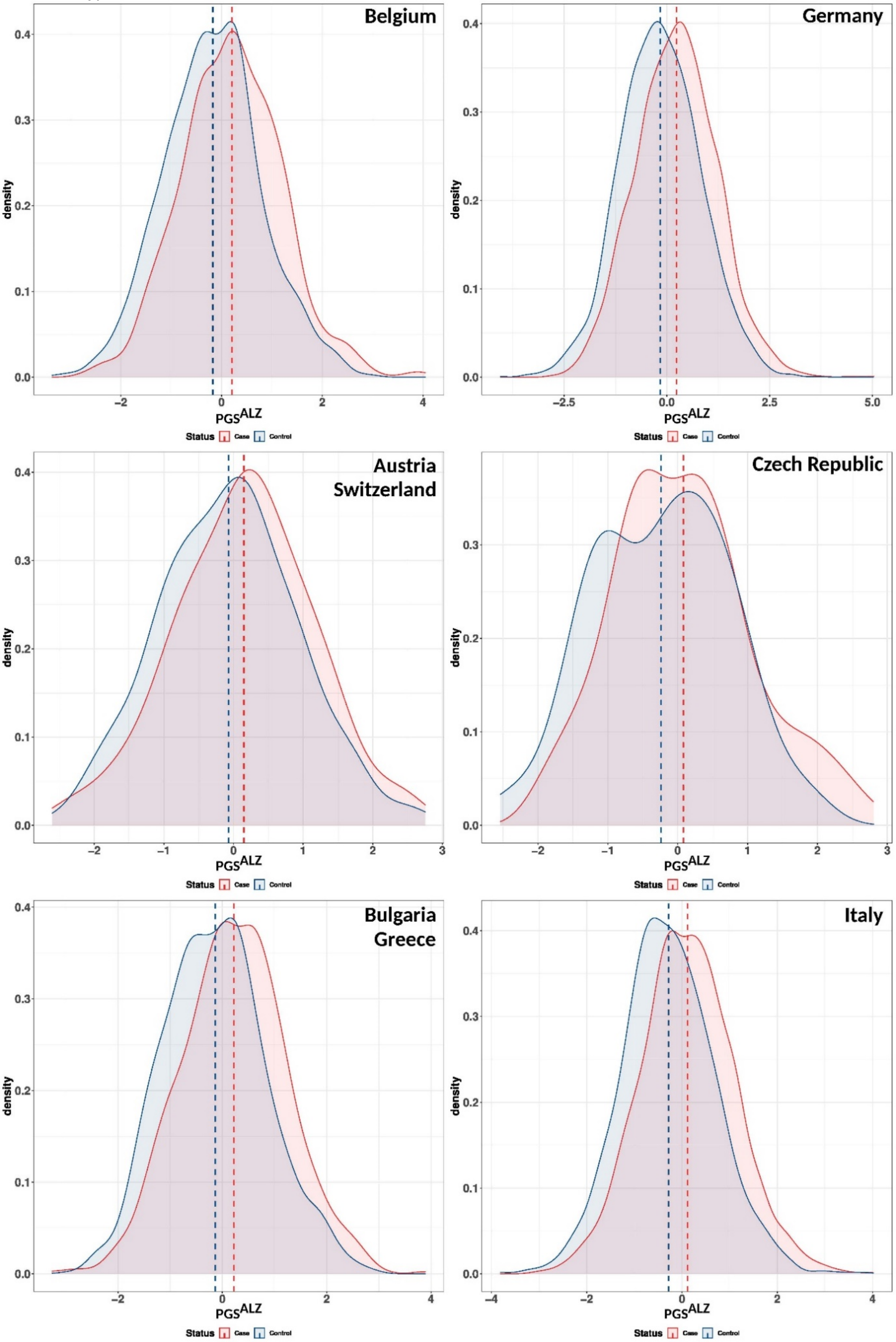

**Supplementary Figure 3:** $PGS^{ALZ}$ distribution (France, Spain, Portugal)

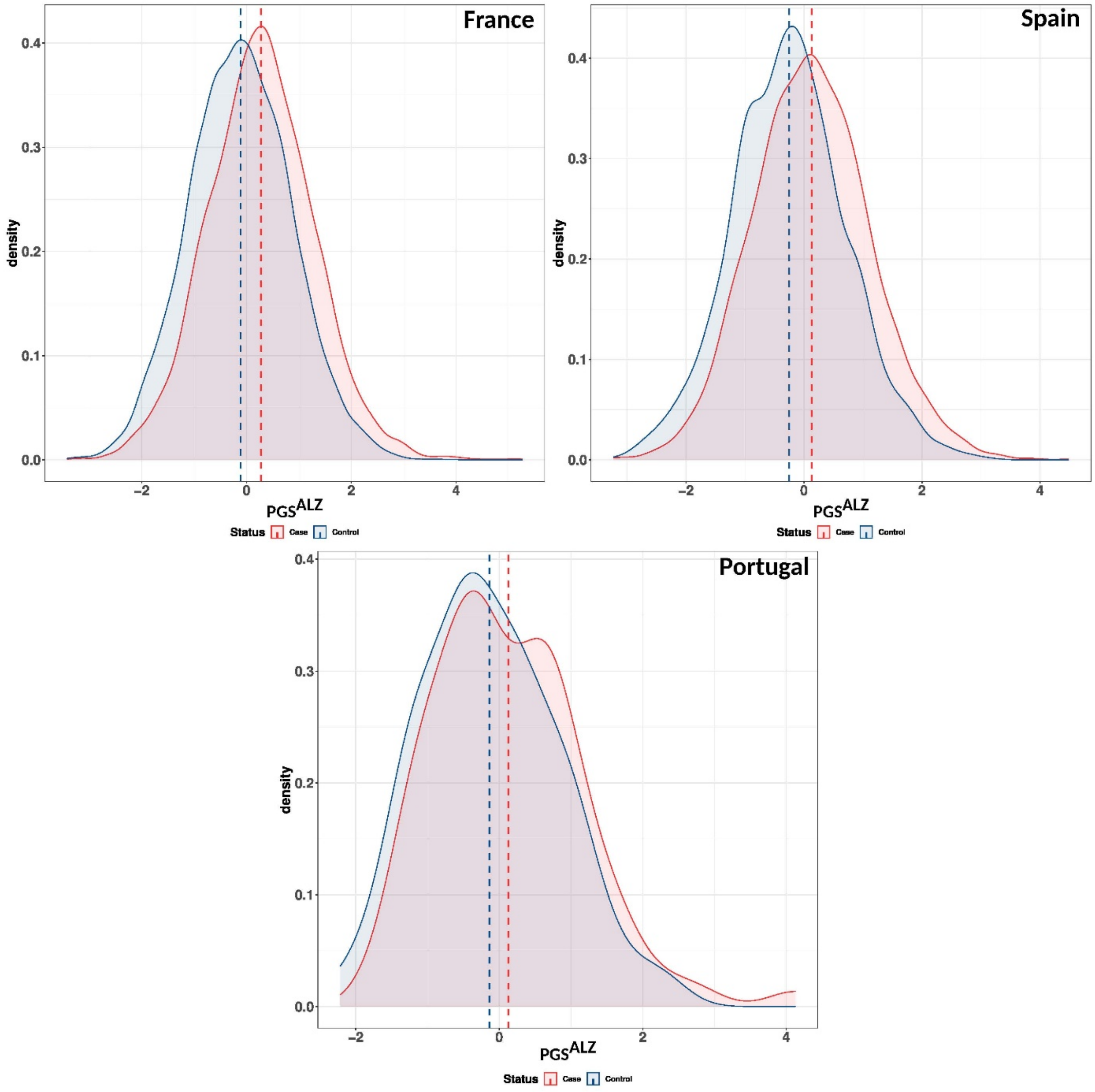

**Supplementary Figure 4:** PGS$^{ALZ}$ association with AD risk across Europe. Ncases, number of cases; Ncontrols, number of controls; OR, Odds ratio. The lines in the Forrest plots indicate the 95% confidence interval for the ORs.

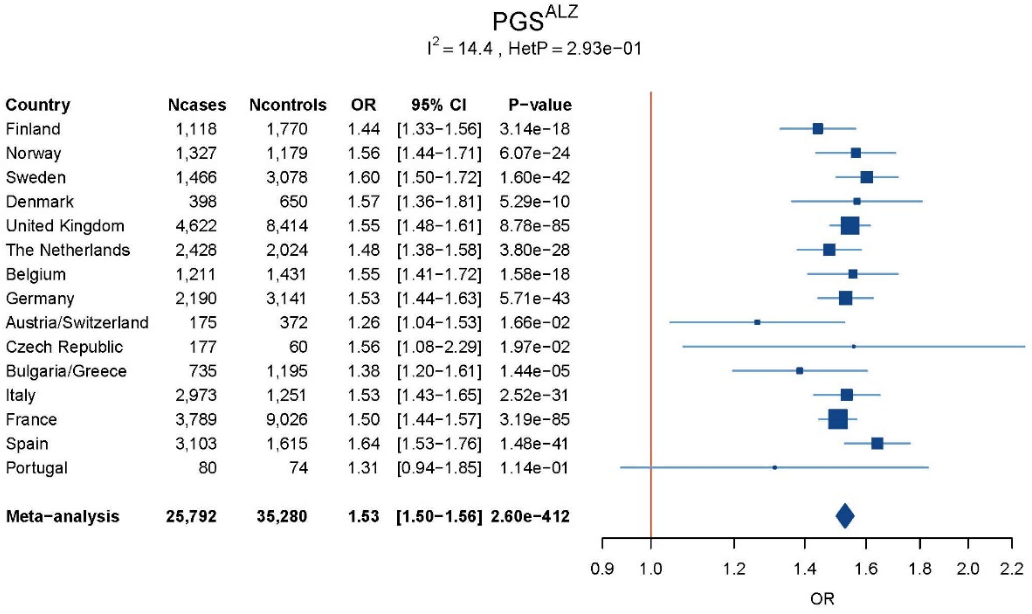

**Supplementary Figure 5:** $PGS^{ALZ}$ association with AD risk across Europe adjusted for difference in distributions across populations. Ncases, number of cases; Ncontrols, number of controls; OR, Odds ratio. The lines in the Forrest plots indicate the 95% confidence interval for the ORs.

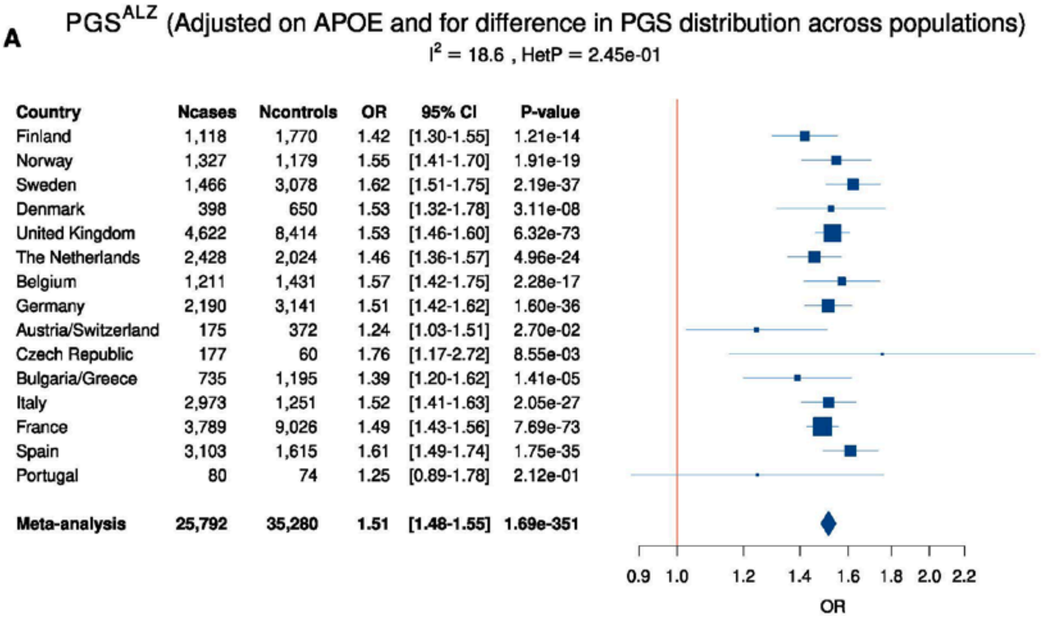


| Country | Ncases | Ncontrols | OR | 95% CI | P-value |
|---|---|---|---|---|---|
| Finland | 1,118 | 1,770 | 1.42 | [1.30-1.55] | 1.21e-14 |
| Norway | 1,327 | 1,179 | 1.55 | [1.41-1.70] | 1.91e-19 |
| Sweden | 1,466 | 3,078 | 1.62 | [1.51-1.75] | 2.19e-37 |
| Denmark | 398 | 650 | 1.53 | [1.32-1.78] | 3.11e-08 |
| United Kingdom | 4,622 | 8,414 | 1.53 | [1.46-1.60] | 6.32e-73 |
| The Netherlands | 2,428 | 2,024 | 1.46 | [1.36-1.57] | 4.96e-24 |
| Belgium | 1,211 | 1,431 | 1.57 | [1.42-1.75] | 2.28e-17 |
| Germany | 2,190 | 3,141 | 1.51 | [1.42-1.62] | 1.60e-36 |
| Austria/Switzerland | 175 | 372 | 1.24 | [1.03-1.51] | 2.70e-02 |
| Czech Republic | 177 | 60 | 1.76 | [1.17-2.72] | 8.55e-03 |
| Bulgaria/Greece | 735 | 1,195 | 1.39 | [1.20-1.62] | 1.41e-05 |
| Italy | 2,973 | 1,251 | 1.52 | [1.41-1.63] | 2.05e-27 |
| France | 3,789 | 9,026 | 1.49 | [1.43-1.56] | 7.69e-73 |
| Spain | 3,103 | 1,615 | 1.61 | [1.49-1.74] | 1.75e-35 |
| Portugal | 80 | 74 | 1.25 | [0.89-1.78] | 2.12e-01 |
| **Meta-analysis** | **25,792** | **35,280** | **1.51** | **[1.48-1.55]** | **1.69e-351** |

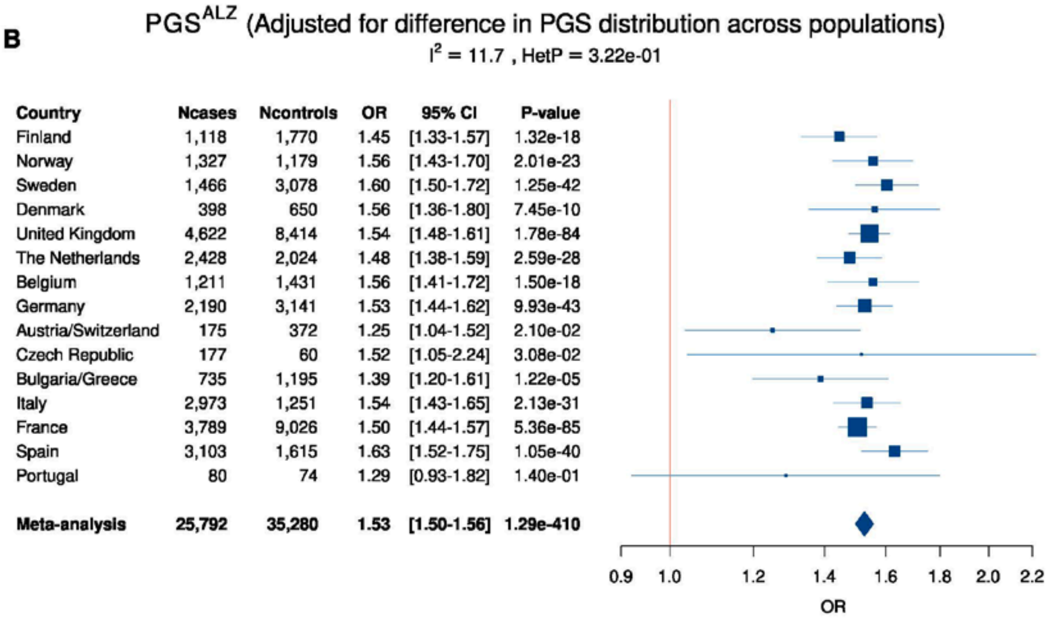


| Country | Ncases | Ncontrols | OR | 95% CI | P-value |
|---|---|---|---|---|---|
| Finland | 1,118 | 1,770 | 1.45 | [1.33-1.57] | 1.32e-18 |
| Norway | 1,327 | 1,179 | 1.56 | [1.43-1.70] | 2.01e-23 |
| Sweden | 1,466 | 3,078 | 1.60 | [1.50-1.72] | 1.25e-42 |
| Denmark | 398 | 650 | 1.56 | [1.36-1.80] | 7.45e-10 |
| United Kingdom | 4,622 | 8,414 | 1.54 | [1.48-1.61] | 1.78e-84 |
| The Netherlands | 2,428 | 2,024 | 1.48 | [1.38-1.59] | 2.59e-28 |
| Belgium | 1,211 | 1,431 | 1.56 | [1.41-1.72] | 1.50e-18 |
| Germany | 2,190 | 3,141 | 1.53 | [1.44-1.62] | 9.93e-43 |
| Austria/Switzerland | 175 | 372 | 1.25 | [1.04-1.52] | 2.10e-02 |
| Czech Republic | 177 | 60 | 1.52 | [1.05-2.24] | 3.08e-02 |
| Bulgaria/Greece | 735 | 1,195 | 1.39 | [1.20-1.61] | 1.22e-05 |
| Italy | 2,973 | 1,251 | 1.54 | [1.43-1.65] | 2.13e-31 |
| France | 3,789 | 9,026 | 1.50 | [1.44-1.57] | 5.36e-85 |
| Spain | 3,103 | 1,615 | 1.63 | [1.52-1.75] | 1.05e-40 |
| Portugal | 80 | 74 | 1.29 | [0.93-1.82] | 1.40e-01 |
| **Meta-analysis** | **25,792** | **35,280** | **1.53** | **[1.50-1.56]** | **1.29e-410** |

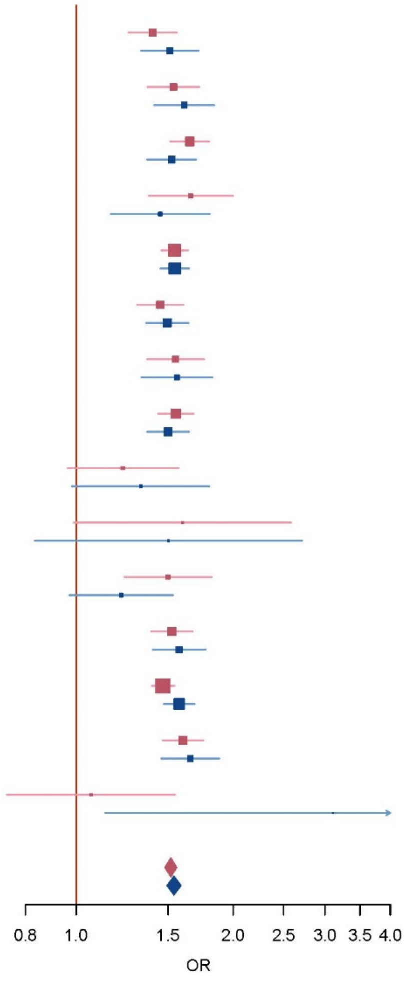

**Supplementary Figure 7:** Association of PGS$^{ALZ}$ with the level of Aβ42, Tau and p-TAU in the cerebrospinal fluid in European-ancestry populations. Ncases, number of cases; Ncontrols, number of controls, OR, Odds ratio. The lines in the Forrest plots indicate the 95% confidence interval for the ORs. If HetP <0.05, random-effect is shown for the meta-analysis results.

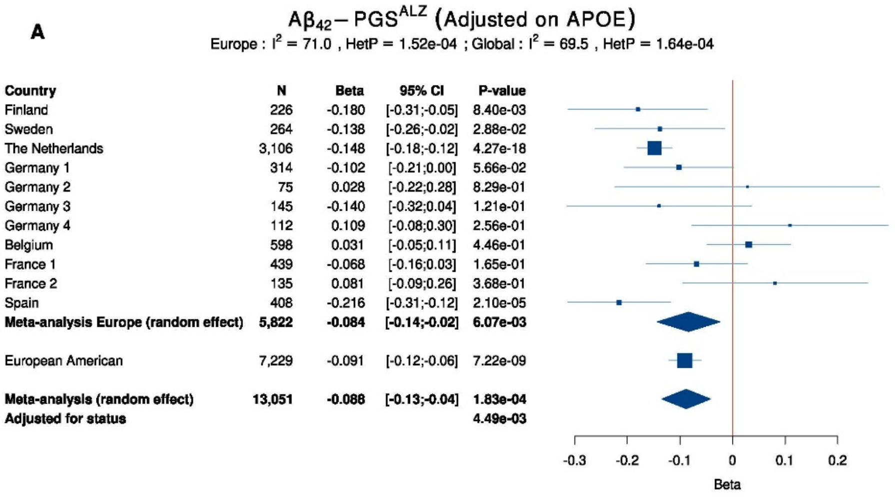


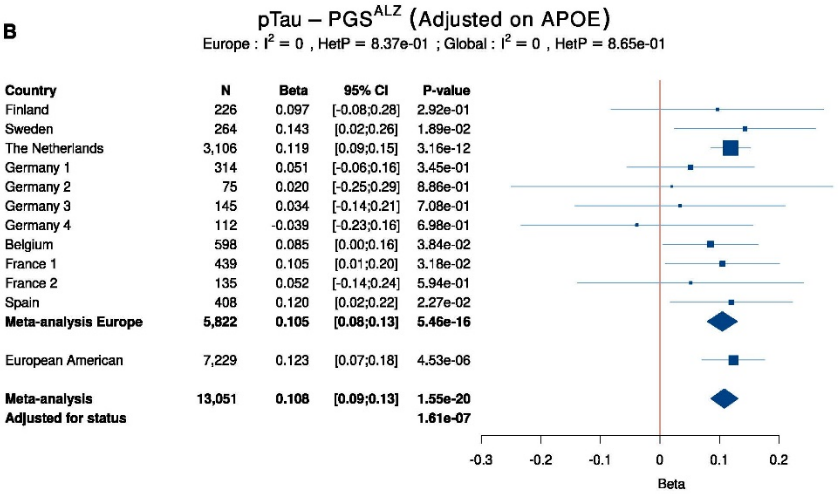


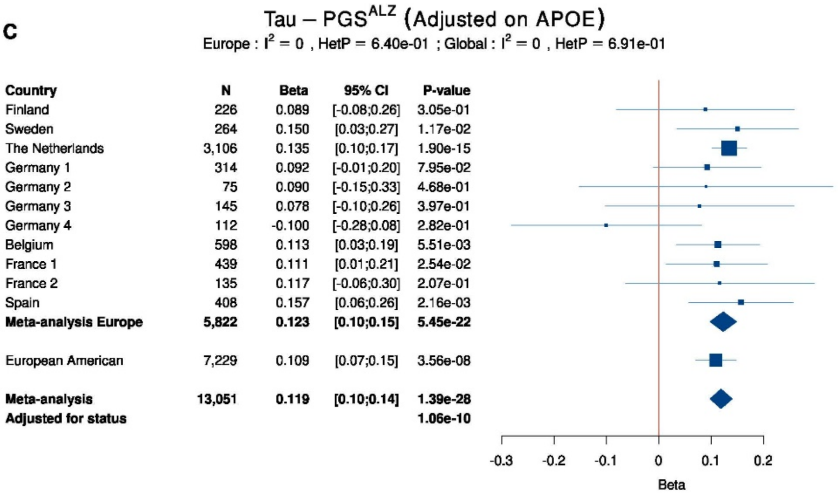

**Supplementary Figure 8:** $PGS^{ALZ}$ distribution (European American MVP, Australia, Maghreb, Sub Saharan Africa, African American MVP, African American ADSP)

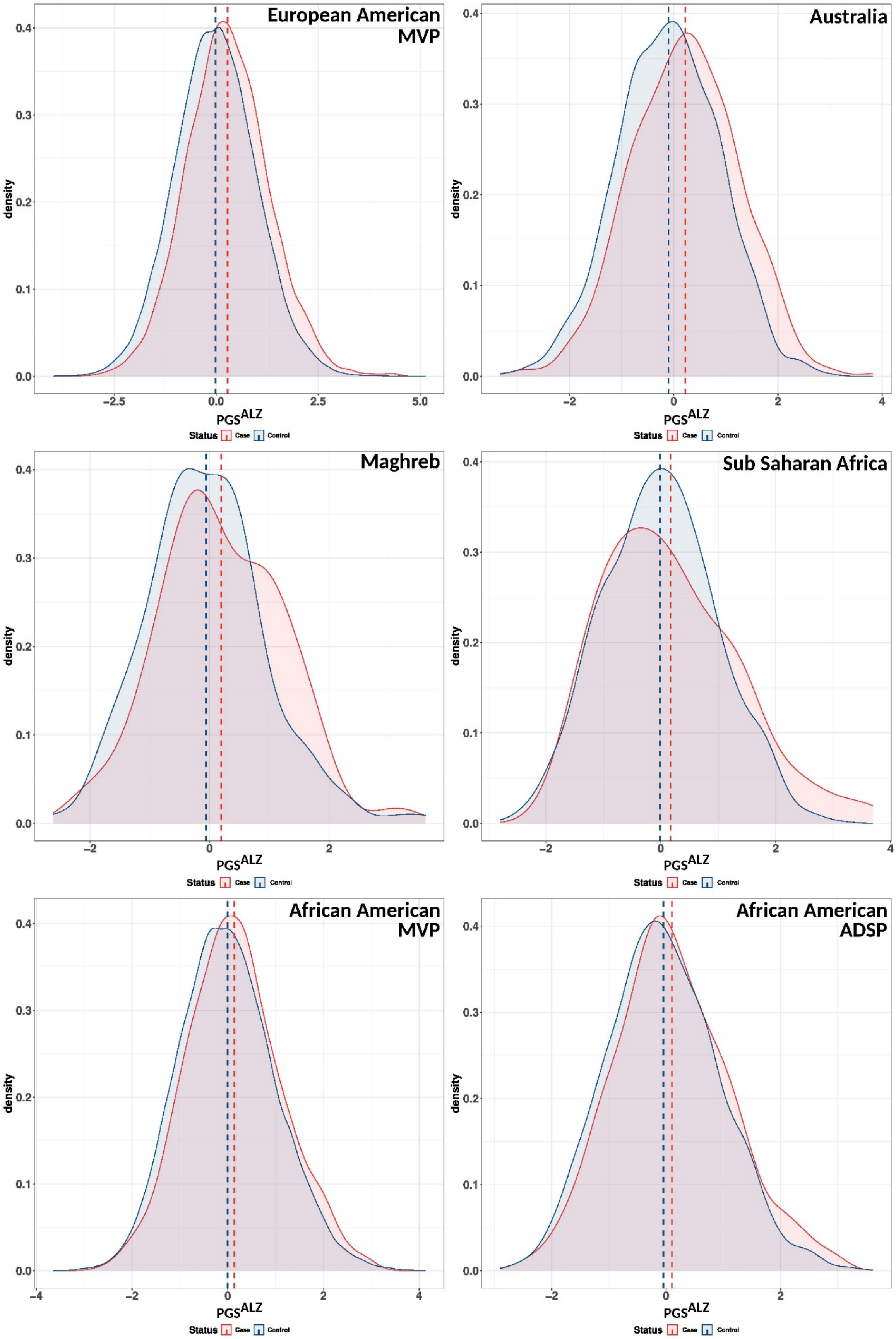

**Supplementary Figure 9:** $PGS^{ALZ}$ distribution (US LA ancestry MVP, US LA ancestry ADSP, Colombia,, Brazil, Argentina, Chile)

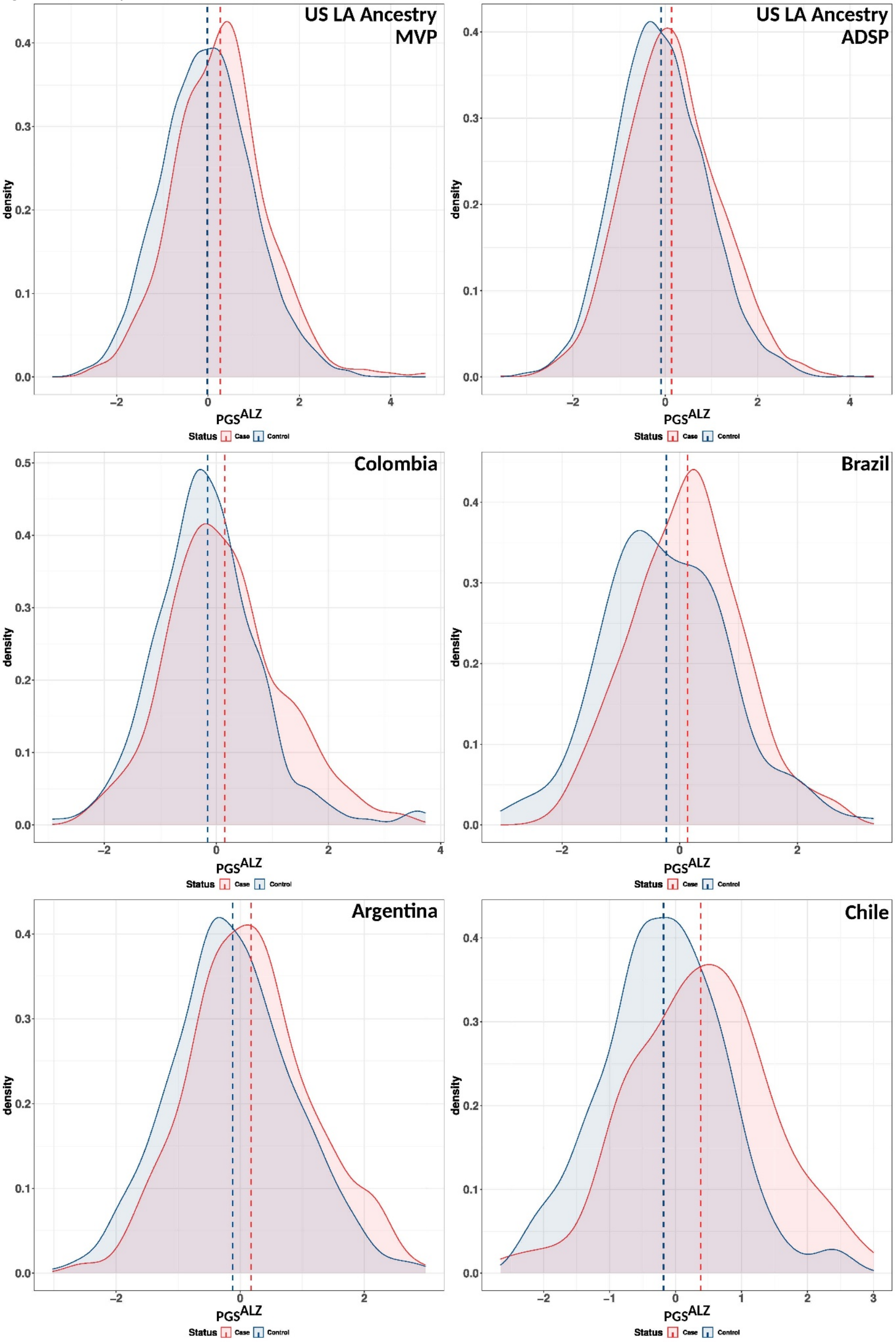

**Supplementary Figure 10:** $PGS^{ALZ}$ distribution (China, Japan, South Korea, India)

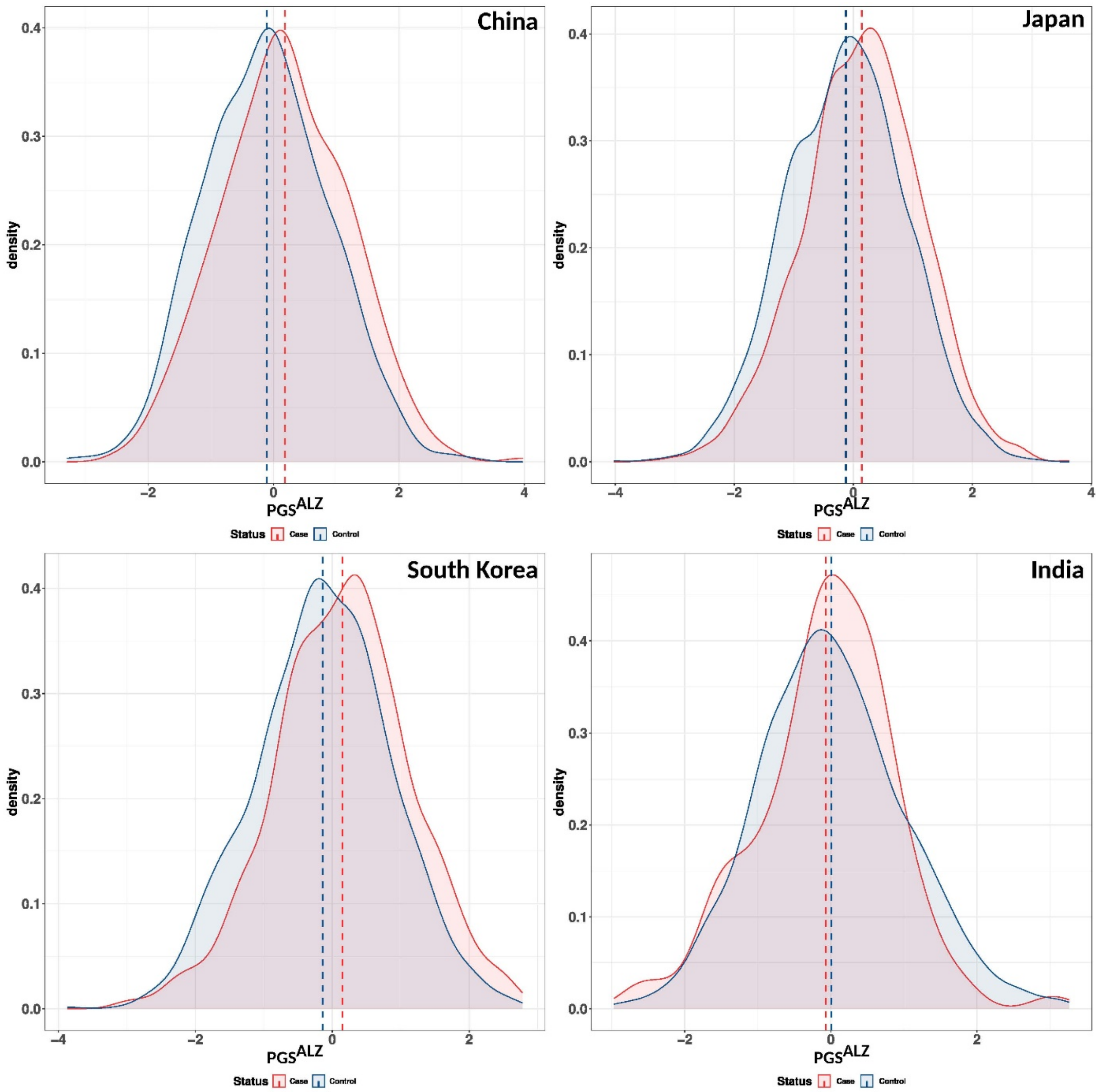

**Supplementary Figure 11:** Association of $PGS^{ALZ}$ with the risk of developing AD in multi-ancestry populations. European ancestry meta-analysis includes MVP and Australia. African-American-ancestry (more than 75% AA ancestry) meta-analysis includes MVP and ADSP. East-Asia meta-analysis includes China, Korea and Japan. Latino-American ancestry (self-reporting) meta-analysis includes MVP, ADSP and Salsa. South-America meta-analysis includes Argentina, Brazil, Chile and Colombia. Ncases, number of cases; Ncontrols, number of controls, OR, Odds ratio. The lines in the Forrest plots indicate the 95% confidence interval for the ORs. If HetP <0.05, random-effect is shown for the meta-analysis results.

$PGS^{ALZ}$

| Country | Ncases | Ncontrols | OR | 95% CI | P−value | HetIsq | HetPVal |
|---|---|---|---|---|---|---|---|
| European American MVP | 4,561 | 84,587 | 1.39 | [1.34−1.44] | 2.40e−81 | | |
| Australia | 544 | 1,190 | 1.39 | [1.25−1.54] | 1.04e−09 | | |
| **Meta−analysis EUR Ancestry** | **5,105** | **85,777** | **1.39** | **[1.34−1.43]** | **1.87e−89** | **0** | **9.99e−01** |
| Maghreb | 58 | 187 | 1.46 | [1.01−2.15] | 4.81e−02 | | |
| Sub−Saharan Africa | 85 | 1,091 | 1.11 | [0.89−1.39] | 3.48e−01 | | |
| African American MVP | 713 | 19,405 | 1.20 | [1.10−1.30] | 5.12e−05 | | |
| African American ADSP | 1,108 | 2,325 | 1.15 | [1.07−1.24] | 1.72e−04 | | |
| **Meta−analysis AA Ancestry** | **1,821** | **21,730** | **1.17** | **[1.11−1.24]** | **4.04e−08** | **0** | **5.31e−01** |
| US LA Ancestry MVP | 375 | 7,166 | 1.30 | [1.15−1.46] | 3.38e−05 | | |
| US LA Ancestry ADSP | 1,629 | 2,456 | 1.26 | [1.18−1.35] | 3.58e−12 | | |
| Colombia | 254 | 241 | 1.27 | [1.04−1.57] | 2.14e−02 | | |
| Brazil | 301 | 178 | 1.57 | [1.28−1.95] | 2.39e−05 | | |
| Argentina | 417 | 601 | 1.32 | [1.14−1.53] | 1.51e−04 | | |
| Chile | 122 | 252 | 1.77 | [1.39−2.28] | 5.60e−06 | | |
| **Meta−analysis US LA Ancestry** | **2,004** | **9,622** | **1.27** | **[1.20−1.35]** | **6.12e−16** | **0** | **7.10e−01** |
| **Meta−analysis South America** | **1,094** | **1,272** | **1.42** | **[1.29−1.56]** | **5.44e−13** | **50.7** | **1.08e−01** |
| **Meta−analysis LA Ancestry** | **3,098** | **10,894** | **1.31** | **[1.25−1.37]** | **1.57e−26** | **50.1** | **7.45e−02** |
| China | 453 | 786 | 1.35 | [1.19−1.54] | 2.33e−06 | | |
| Japan | 2,220 | 2,472 | 1.30 | [1.22−1.38] | 1.76e−17 | | |
| South Korea | 1,119 | 1,172 | 1.35 | [1.24−1.47] | 5.57e−12 | | |
| **Meta−analysis East Asia** | **3,792** | **4,430** | **1.32** | **[1.26−1.38]** | **1.32e−32** | **0** | **6.75e−01** |
| India | 105 | 1,500 | 0.97 | [0.79−1.19] | 7.85e−01 | | |

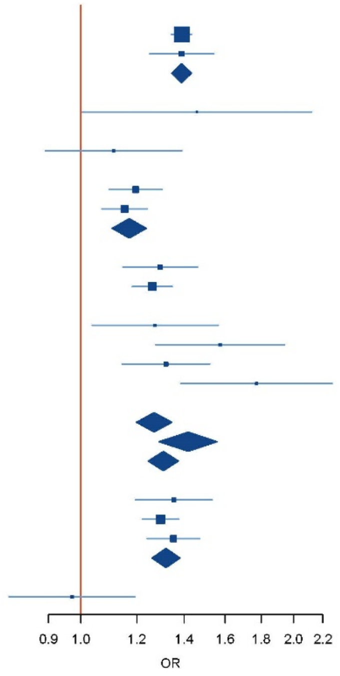

**Supplementary Figure 12.** Association of PGS$^{APOE}$ with age at onset in multi-ancestry populations. European ancestry meta-analysis includes MVP and Australia. African-American-ancestry (more than 75% AA ancestry) meta-analysis includes MVP and ADSP. East-Asia meta-analysis includes China, Korea and Japan. Latino-American ancestry (self-reporting) meta-analysis includes MVP, ADSP and Salsa. South-America meta-analysis includes Argentina, Brazil, Chile and Colombia. Ncases,number of cases. The lines in the Forrest plots indicate the 95% confidence interval for the ORs. If HetP <0.05, random-effect is shown for the meta-analysis results.

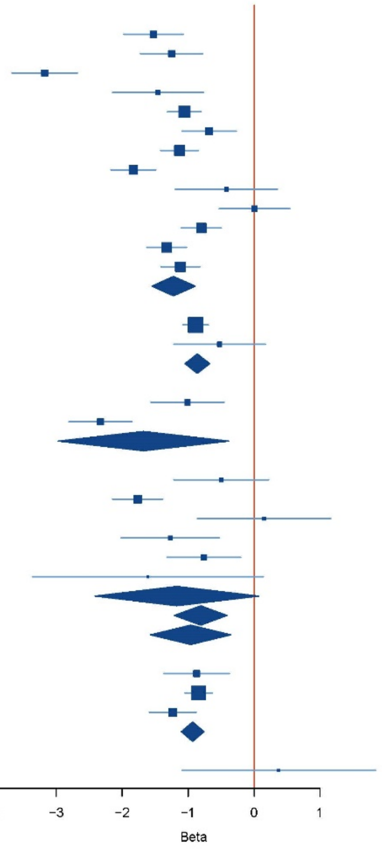

**Supplementary Figure 13:** Workflow indicating the summary statistics and populations used to develop and test PGS$^{ALZ}$, PGS$^{ALZ+}$ and PRS.

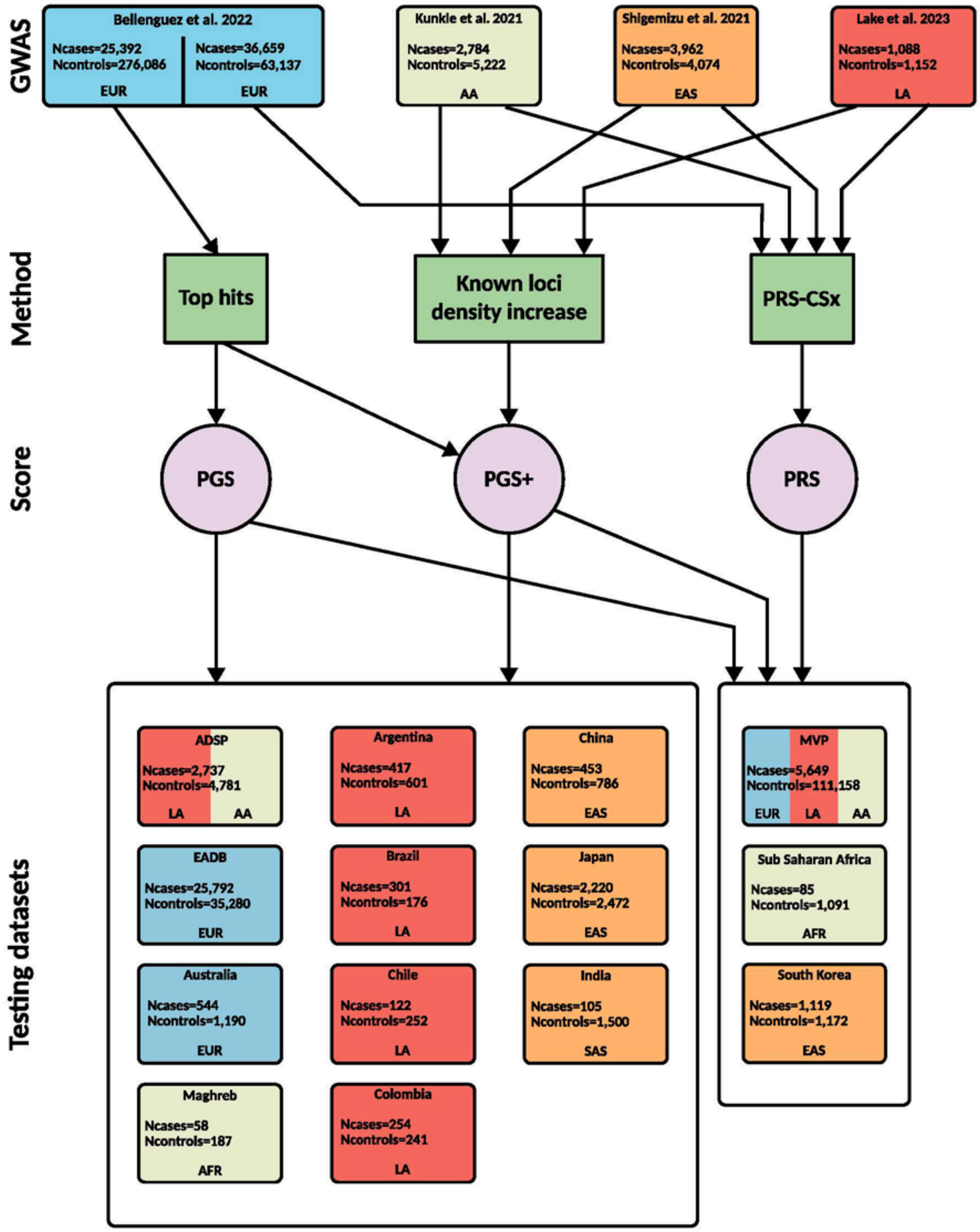

**Figure 14:** Comparison of the association of $PGS^{ALZ}$ or PRS (excluding APOE region) with AD risk and the corresponding predictive values (adjusted Nagelkerke R2 and Liability R2) in EPIDEMCA. All $PGS^{ALZ}$ and PRS were adjusted for difference in distribution between populations; OR, Odds ratio per standard deviation; $PRS^{EUR}$ were generated by using only European ancestry summary statistics; $PRS^{COMB}$ were generated by combining European, African American (AA), Latin-American (LA) and East Asian ancestry summary statistics. Sparseness parameter at $10^{-8}$, $10^{-7}$, $10^{-6}$, $10^{-5}$, $10^{-4}$, $10^{-3}$, $10^{-2}$ or 1.

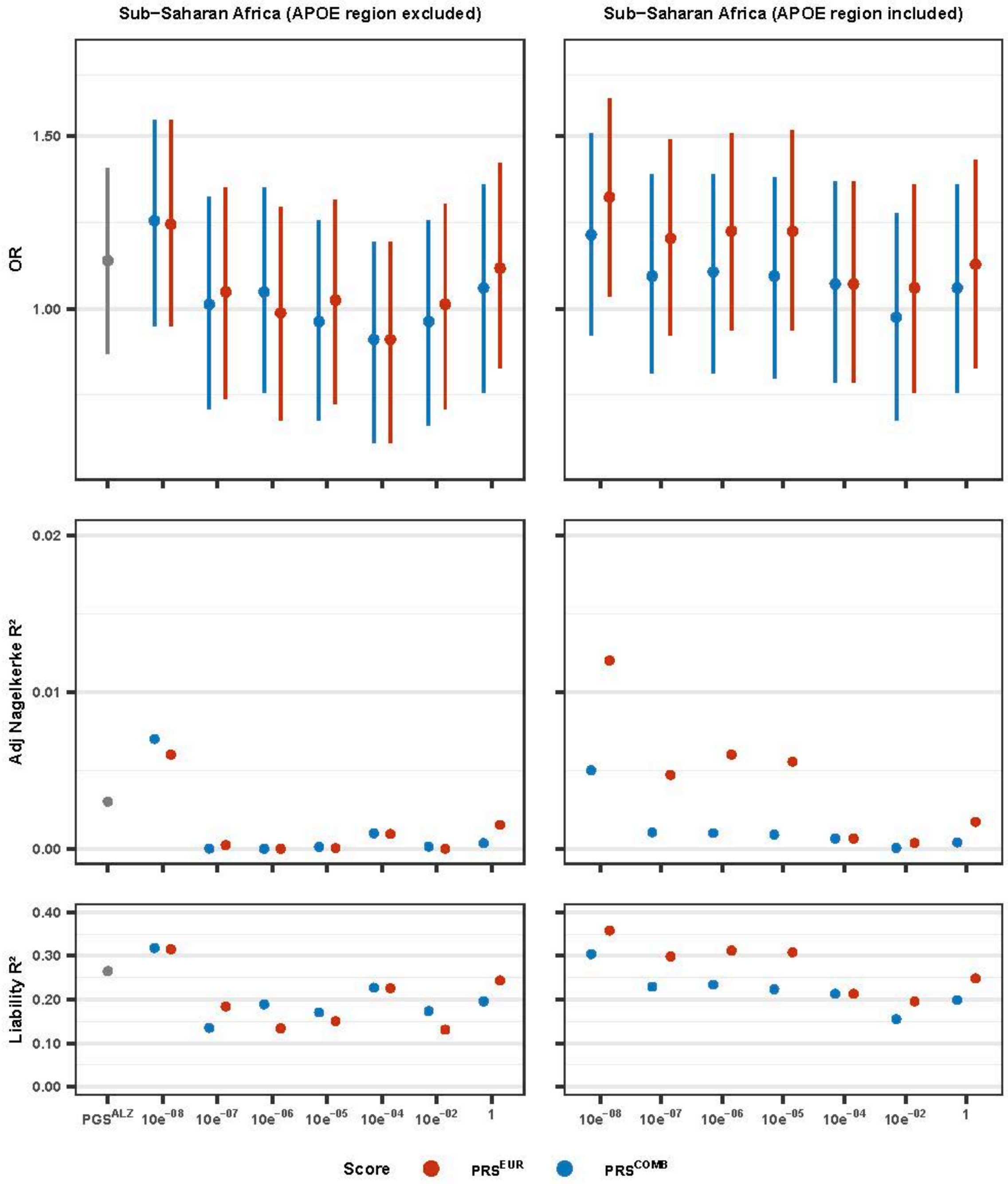

### REFERENCES


1. De Roeck, A. et al. An intronic VNTR affects splicing of ABCA7 and increases risk of Alzheimer's disease. Acta Neuropathol. 135, 827–837 (2018).
2. Sheardova, K. et al. Czech Brain Aging Study (CBAS): Prospective multicentre cohort study on risk and protective factors for dementia in the Czech Republic. BMJ Open 9, (2019).
3. Steinberg, S. et al. Loss-of-function variants in ABCA7 confer risk of Alzheimer's disease. Nat. Genet. 47, 445–447 (2015).
4. Ngandu, T. et al. A 2 year multidomain intervention of diet, exercise, cognitive training, and vascular risk monitoring versus control to prevent cognitive decline in at-risk elderly people (FINGER): A randomised controlled trial. Lancet 385, 2255–2263 (2015).
5. Hanon, O. et al. Plasma amyloid levels within the Alzheimer's process and correlations with central biomarkers. Alzheimer's Dement. 14, 858–868 (2018).
6. Dufouil, C. et al. Cognitive and imaging markers in non-demented subjects attending a memory clinic: Study design and baseline findings of the MEMENTO cohort. Alzheimer's Res. Ther. 9, (2017).
7. Nicolas, G. et al. Screening of dementia genes by whole-exome sequencing in early-onset Alzheimer disease: Input and lessons. Eur. J. Hum. Genet. 24, 710–716 (2016).
8. McKhann, G. et al. Clinical diagnosis of Alzheimer's disease: report of the NINCDS-ADRDA Work Group under the auspices of Department of Health and Human Services Task Force on Alzheimer's Disease. Neurology 34, 939–44 (1984).
9. Kornhuber, J. et al. Early and differential diagnosis of dementia and mild cognitive impairment: Dement. Geriatr. Cogn. Disord. 27, 404–417 (2009).
10. Luck, T. et al. Mild cognitive impairment in general practice: Age-specific prevalence and correlate results from the German study on ageing, cognition and dementia in primary care patients (AgeCoDe). Dement. Geriatr. Cogn. Disord. 24, 307–316 (2007).
11. McKhann, G. M. et al. The diagnosis of dementia due to Alzheimer's disease: Recommendations from the National Institute on Aging-Alzheimer's Association workgroups on diagnostic guidelines for Alzheimer's disease. Alzheimer's Dement. 7, 263–269 (2011).
12. Van Der Flier, W. M. & Scheltens, P. Amsterdam dementia cohort: Performing research to optimize care. Journal of Alzheimer's Disease vol. 62 1091–1111 (2018).
13. Holstege, H. et al. The 100-plus Study of cognitively healthy centenarians: rationale, design and cohort description. Eur. J. Epidemiol. 33, (2018).
14. Aalten, P. et al. The Dutch Parelsnoer Institute - Neurodegenerative diseases; methods, design and baseline results. BMC Neurol. 14, (2014).
15. Dubois, B. et al. Research criteria for the diagnosis of Alzheimer's disease: revising the NINCDS-ADRDA criteria. Lancet Neurology vol. 6 734–746 (2007).
16. Ramakers, I. et al. Biobank Alzheimer Center Limburg cohort: design and cohort characteristics. Prep.
17. (APA), A. P. A. Diagnostic and statistical manual of mental disorders. (1994).
18. Sachdev, P. S. et al. The Sydney Memory and Ageing Study (MAS): Methodology and baseline medical and neuropsychiatric characteristics of an elderly epidemiological non-demented cohort of Australians aged 70-90 years. Int. Psychogeriatrics 22, 1248–1264 (2010).
19. Moreno-Grau, S. et al. Genome-wide association analysis of dementia and its clinical endophenotypes reveal novel loci associated with Alzheimer's disease and three causality networks: The GR@ACE project. Alzheimer's Dement. 15, 1333–1347 (2019).
20. Ruiz, A. et al. Assessing the role of the TREM2 p.R47H variant as a risk factor for Alzheimer's disease and frontotemporal dementia. Neurobiol. Aging 35, 444.e1–4 (2014).
21. Ikram, M. A. et al. The Rotterdam Study: 2018 update on objectives, design and main results. Eur. J. Epidemiol. 32, 807–850 (2017).
22. Niemeijer, M. N. et al. ABCB1 gene variants, digoxin and risk of sudden cardiac death in a general population. Heart 101, 1973–1979 (2015).
23. Loh, P.-R. et al. Reference-based phasing using the Haplotype Reference Consortium panel. Nat. Genet. 48, 1443–1448 (2016).
24. De Bruijn, R. F. A. G. et al. Determinants, MRI correlates, and prognosis of mild cognitive impairment: The rotterdam study. in Journal of Alzheimer's Disease vol. 42 S239–S249 (IOS Press, 2014).
25. 3C Study Group. Vascular factors and risk of dementia: design of the Three-City Study and baseline characteristics of the study population. Neuroepidemiology 22, 316–25 (2003).
